# Linguistic vitality improves health and wellbeing in Indigenous communities: a scoping review

**DOI:** 10.1101/2024.08.20.24310741

**Authors:** Louise Harding, Ryan DeCaire, Karleen Delaurier-Lyle, Ursula Ellis, Julia Schillo, Mark Turin

## Abstract

**Introduction:** While Indigenous communities have long recognized the importance of their languages for their wellbeing, this topic has only recently received attention in scholarship, research and public policy. This scoping review synthesizes and assesses existing literature on the links between Indigenous linguistic vitality and health or wellness in English-speaking settler colonial countries (Australia, Canada, New Zealand, and the United States).

**Methods:** The JBI methodology for scoping reviews was followed by an interdisciplinary research team. Key databases searched included MEDLINE, PsycInfo, and Cumulative Index to Nursing and Allied Health Literature. Searches were restricted to English language literature. The last search was on February 8, 2021. Quantitative and qualitative analyses were conducted to categorize and elucidate the nature of the links reported.

**Results:** Over 10,000 records were reviewed and 262 met the inclusion criteria – 70% academic and 30% gray literature. The largest number of studies focus on Canadian contexts. 78% of the original research studies report only supportive links between Indigenous languages and health, while 98% of the literature reviews report supportive links. Linguistic vitality tends to support health and wellness outcomes, while the diminishment of languages is associated with worse health. The most prevalent links with linguistic vitality are healthcare outcomes, overall health and healing, and mental, cognitive, and psychological health and development. The results of the remaining original research studies were mixed (10%), statistically non-significant (6%), adverse (5%) and neutral (1%).

**Conclusions:** The results of this scoping review suggest that linguistic vitality is a determinant of health for Indigenous peoples in the contexts studied. Recommendations for harnessing the healing effects of language include increasing tangible support to language programs, delivering linguistically tailored health care and promotion, and advancing knowledge through funding relevant community-engaged research and education.

**Article Highlights:**

- Our interdisciplinary authorship team conducted a scoping review involving the screening of nearly 10,000 publications.
- 262 academic and grey literature reports were published between 1949-2019 which had a central focus on exploring the links between Indigenous languages and health.
- 78% of original research studies and 98% of reviews report exclusively supportive links between linguistic vitality and the health and wellness of Indigenous peoples.
- Key areas linked to health include healthcare outcomes, overall health and healing, and mental, cognitive, and psychological health and development.
- Linguistic vitality is a determinant of health for Indigenous peoples and should be leveraged through supporting language programs, delivering linguistically tailored care, and funding community-engaged research and education.

## 1. Introduction

> “In Navajo we say nihizaad hiná ‘our language is alive.’ This idea [and the worldview that comes with it] represents a very different epistemological orientation than one that focuses on language only for its utility or potential for economic prosperity. Our commitments to our languages are commitments to who we are as peoples. It is within us to keep them alive.” (McKenzie, 2020, p. 506)

Each language offers a unique way of understanding and categorizing the world, reflecting insights into history, knowledge, and culture (Sapir, 1929; Shaw, 2001). Given this connection, the decline in the use of Indigenous, minoritized and under-resourced languages— through colonization, displacement, denigration, and assimilative education policies—has had and continues to have dramatic and deleterious effects on speech communities (United Nations Economic and Social Council, 2016). In recent years, the nature of the connection between linguistic vitality and Indigenous community health has gained increasing attention among scholars, policy makers, and language practitioners (First Peoples’ Cultural Council, 2018; Hallett et al., 2007; House of Representatives Standing Committee on Aboriginal and Torres Strait Islander Affairs, 2012; McIvor et al., 2009). Curiously, while a growing body of publications support and assert the existence of a link between linguistic vitality and health, a relatively limited body of research data and evidence is currently leveraged to tell this story. In this scoping review, we synthesize, analyze and assess the current evidence on the connections between Indigenous languages and health to inform supportive policy and decisions about resource allocation.

### 1.1 Indigenous linguistic vitality and revitalization

Language endangerment and language oppression are global issues (Pine & Turin, 2017; Roche, 2022). The United Nations has estimated that 40% of the languages spoken around the world today are at risk of disappearing as colonial languages like English and Spanish dominate other speech forms (Requesens-Galnares, 2023; Turin, 2012). Indigenous languages are particularly threatened, resulting in the recognition of the rights to use, speak and transmit Indigenous languages being codified in the United Nations Declaration of Rights of Indigenous Peoples (UNDRIP) (United Nations General Assembly, 2007), and the subsequent announcement of 2022-2032 as the International Decade of Indigenous Languages (UNESCO, 2020).

In this review, we focus on the languages spoken by the original inhabitants of Canada, the United States of America, Australia, and Aotearoa New Zealand, who share the experience of Anglo settler colonialism. Settler colonialism, as described by historian Patrick Wolfe, is a system which “strives for the dissolution of native societies” and “erects a new colonial society on the expropriated land base” (2006, p. 388). The term settler refers to the founders of these colonial societies, their descendants, and subsequent immigrants. Anglo settler colonialism specifically aims to replace or assimilate Indigenous populations with a new society of English-speaking settlers, and often involves a government-sanctioned commitment to English language monolingualism (Smandych, 2013; Veracini, 2010). An exception to Anglophone dominance within this context is French dominance in Eastern Canada, and particularly in the province of Quebec, where it is the only official language and exerts pressures on Indigenous languages similar to that exerted by English in the rest of Canada (Patrick, 2005). In all four countries included in our scoping review, attempts to forcibly introduce colonial languages such as English have included prohibiting the use of Indigenous languages (Mako, 2012), forcibly removing children from their homes and communities (Canadian Council of Provincial Child and Youth Advocates, 2010; Royal Commission of Aboriginal Peoples, 1996; Wilson, 1997), and denigrating Indigenous language speakers (The Aboriginal Healing Foundation, 2006). The results have been devastating for many Indigenous communities and for the flourishing of their languages (L. Davis, 2017).

In this context, Indigenous-led initiatives to maintain, revitalize, and reclaim languages are acts of Indigenous sovereignty, resistance and resurgence (J. L. Davis, 2018; Perley, 2011; Simpson, 2017). These very often take the form of multidisciplinary, heterogeneous, innovative interventions to offset the global decline in linguistic diversity (Pine & Turin, 2017). By way of tangible example, on the eastern side of Canada, co-author RDC is a Kanien’kéha (Mohawk language) learner, instructor and researcher, whose work is principally focused on developing and employing Kanien’kéha adult immersion programs which serve as a pathway for adults to become highly proficient in Kanien’kéha so that they can create and foster immersion and Kanien’kéha-medium environments in critical community domains and thus work to restore intergenerational transmission. Similarly, co-author MT’s work with the Heiltsuk Language and Culture Mobilization Partnership in British Columbia is aimed at creating “new opportunities for speaking, writing and reading the Heiltsuk language” [Híɫzaqv/Háiɫzaqv] with the objective to “expand and deepen existing community language revitalization and cultural documentation in a digital environment,” (J. Carpenter et al., 2021, p. 2).

In recent decades, governments have made some moves to recognize Indigenous language rights (Skutnabb-Kangas & Phillipson, 2023), including designating official language status for the Indigenous te reo Māori language in Aotearoa New Zealand (Māori Language Act, 1987; Te Ture Mō Te Reo Māori/ Māori Language Act, 2016), introducing Indigenous language acts in the USA (Native American Languages Act, 1990), Canada (Indigenous Languages Act, 2019), and the state of New South Wales in Australia (Aboriginal Languages Act, 2017). While such efforts have been seen by some as encouraging steps, there is not always a direct link between the granting of rights or recognition and tangible, meaningful, long-term support for Indigenous communities (Coulthard, 2014; Inutiq, 2016; Patrick, 2005). Indigenous language revitalization initiatives continue to be challenged by a lack of sustainable, consistent, and dedicated funding and supportive legislative policies (K. Carpenter & Tsykarev, 2020; Haque & Patrick, 2015; Hinton & Meek, 2018; Olawsky, 2020).

### 1.2 Indigenous languages and health

While Indigenous communities know in embodied and tangible ways of the connections between the vitality of their languages, health, and well-being, a growing awareness of these connections among the broader public and particularly within public health, language sciences, and other scientific communities can be used to help advance the case for resource allocation and public policies that are dedicated to supporting linguistic diversity and language revitalization.

For example, a groundbreaking 2007 study by Hallett and colleagues reported a strong correlation between community-level Indigenous language knowledge and reduced youth suicide rates in British Columbia First Nations communities (Hallett et al., 2007). The report captured the attention of Canada’s Prime Minister Justin Trudeau who referenced it in an interview as an example of why it is important to support Indigenous communities to “teach language and culture and take pride in identity” (House, 2016). While the data that was analyzed in that study is now over 25 years old, it continues to have a significant influence worldwide.

Our aim with this scoping review is to have lasting impact by conducting the most comprehensive, systematic, and rigorous synthesis and assessment of evidence to date on the links between Indigenous languages and health written in English, and to deliver recommendations based on our findings. Further, we hope to clarify the nature of the links between Indigenous languages and health to allow more nuance and specificity in relevant discussions and initiatives. To that end, we apply a scoping review methodology to investigate a simple research question:

> What is the extent and scope of the literature on the relationships between Indigenous linguistic vitality and the health/wellness of Indigenous populations in Australia, Canada, New Zealand, and the United States of America?

## 2. Methods

### 2.1 Positionality statement and description of collaboration

This work was led by Mark Turin (MT) and Louise Harding (LH) who sought collaborative input, critical advice, and insight from Ryan DeCaire (RDC), Karleen Delaurier-Lyle (KDL), Ursula Ellis (UE) and Julia Schillo (JS). Ryan DeCaire is Kanien’kehá:ka (Mohawk) and Karleen Delaurier-Lyle is of Anishinabek and Cree mixed-settler ancestry and a member of the Berens River First Nation. Mark Turin, Louise Harding, Ursula Ellis, and Julia Schillo are non-Indigenous settler academics. Members of our multidisciplinary team brought knowledge in fields spanning the information sciences, Indigenous language documentation and revitalization, linguistics, population and public health, ethics, food sovereignty, and Indigenous wellness. In the remainder of the manuscript, we use acronyms to refer to the authors.

### 2.2 Protocol and registration

Our protocol was drafted in accordance with the Joanna Briggs Institute (JBI) methodology for scoping reviews, setting out the background, research question, eligibility criteria for studies, and methods (Peters et al., 2020). We selected a scoping review methodology because it offers a systematic approach to collecting and synthesizing literature that allows for a broad research question. Further, it does not require evidence appraisal, which can at times be problematic when Western research tools are applied to evaluate Indigenous ways of knowing due to key epistemological, ontological, and methodological differences (Chambers et al., 2018; Kovach, 2021; Smith, 2021). Further, the scoping review approach allowed us to survey the full range of variables relating to linguistic vitality, revitalization and language use as well as to health and wellness, rather than focusing narrowly on highly specific independent and dependent variables at this relatively early stage of inquiry. We included gray literature in our review, or literature published outside of traditional academic publishing channels, to include publications that might be relevant to Indigenous communities. The gray literature search strategy was guided by a template produced by Jackie Stapleton, based on the methods used in a systematic review by Godin et al. (2015). Our final protocol was registered prospectively with the Open Science Framework on 26 January 2021 (Appendix A) (Harding, Delaurier-Lyle, et al., 2021). Reporting follows the PRISMA-Scr Checklist (Appendix B).

### 2.3 Eligibility criteria

In order to be included in this review, literature was required to have a central focus on making connections between the health or wellbeing of Indigenous communities and Indigenous linguistic vitality, including language use, knowledge, maintenance, transmission, teaching, revitalization, and reclamation, as well as other structures and initiatives that support or undermine linguistic vitality.

Health and wellbeing were considered broadly in an attempt to encompass a wide range of the worldviews represented in the literature (Boddington & Raisanen, 2009). Individual and community wellness were included within the full spectrum of prevention, determinants of health, health promotion, health outcomes, healing, and spiritual wellbeing. Measures of community wellbeing that were outside of this scope included economic wellbeing, graduation rates, employment rates, and housing adequacy.

Indigenous communities in Canada, the United States of America and its territories, Australia, and the Realm of New Zealand were included. Even though the French language is dominant in Quebec and other parts of eastern Canada, we chose to include these regions for reasons of feasibility and on account of common contextual features that are shared with the rest of Canada, including ties to England through the commonwealth. Further, as French has co-official language status in Canada, it has a high degree of politically and culturally reinforced esteem and thus receives greater support than do Indigenous languages (L. Davis, 2017).

Eligible sources of evidence for our review included academic articles, book chapters indexed in academic databases, theses (undergraduate and graduate), dissertations, conference proceedings and abstracts, course syllabi, and government documents written in English. We also included reports and fact sheets published by various stakeholders including universities, associations, non-governmental organizations (NGOs), intergovernmental organizations (IGOs) and research agencies. We excluded informal communications such as blogs, emails, and social media posts, magazines, newspaper articles, trade publications, non-scholarly books, and books published by vanity or predatory publishers. We only included literature published in English due to feasibility limitations including the language knowledge of the study team, limited resources, and in recognition of the global dominance of English in the landscape of academic publishing. No date limits were applied to inclusion.

### 2.4 Information sources & search

We searched the following academic bibliographic databases on January 26, 2021: MEDLINE (Ovid), PsycInfo (EBSCO) Cumulative Index to Nursing & Allied Health Literature (CINAHL Complete: EBSCO), Bibliography of Native North Americans (EBSCO), Australian Education Index (ProQuest), Educational Resource Information Centre (ERIC: EBSCO), MLA International Bibliography (EBSCO), Web of Science Core Collection, Communication & Mass Media Complete (EBSCO), and Linguistics and Language Behavior Abstracts (LLBA: ProQuest). To locate unpublished studies and gray literature, we searched the following databases between January 26 and February 8, 2021: desLibris, PsycEXTRA (EBSCO), Native Health Database, iPortal, ProQuest Dissertations & Theses Global, Theses Canada, OAIster, Analysis & Policy Observatory, Informit Indigenous Collection, and Advanced Google. To complete our gray literature search strategy, we also conducted targeted website searching and solicited articles from members of the research team and their colleagues. Finally, we screened the reference lists of all included sources to locate additional studies that might be relevant.

The search strategies were drafted by LH with librarians UE and KDL, piloted in Ovid MEDLINE, and discussed with other members of the research team (Appendix C and D). We created a list of keywords and subject headings which pertained to Indigenous populations in each of the four countries as well as to language, linguistics, revitalization, and reclamation. LH executed the search of all databases. Duplicates were automatically identified by the reference management software Zotero and manually removed. The remaining duplicates were automatically identified and removed by the screening and data extraction tool Covidence.

### 2.5 Selection of sources of evidence

To establish congruence, LH and JS conducted two pilot tests of the inclusion criteria and screening process on 35 titles and abstracts in November 2020 and January 2021. Following the official search, LH and JS independently screened all titles and abstracts from the academic databases for assessment against the inclusion criteria using Covidence and then sought consensus. LH subsequently screened the full texts of all articles that had been included. LH independently screened the gray literature sources in two stages analogous to the academic database records screening process. MT assisted in resolving any discrepancies or ambiguities in study selection. We noted reasons for exclusion of sources of evidence at the full text review stage.

### 2.6 Data charting process

We used studies, rather than reports or publications, the units of interest, to avoid double counting since studies can be reported in several sources (e.g. in both a journal article and a dissertation) (Li et al., 2022). Therefore, we identified and collated multiple reports of single studies before completing data charting. LH and MT jointly developed a data-charting form that was iteratively refined throughout the extraction process (Appendix E), after which LH independently charted the data. After piloting the form on a sample of 61 diverse studies from the academic and gray literature, the research team met to review the preliminary findings and to discuss refinements to the data-charting form and synthesis strategy before completing the data charting process.

### 2.7 Data items

Data from each study was abstracted on year of publication, geographic coverage, whether it was academic or gray literature, and the specific type of publication. Studies were classified as original research if the authors presented new data, literature reviews if they primarily synthesized or commented on existing research, or mixed if they presented a balance of the two. Impactful quotes were noted, and brief summaries were prepared of each study’s conclusions on the links between languages and health. To promote impartial reporting of results, only non-evaluative, dispassionate language was used in the summaries and the conclusions were drawn directly from the publications without further interpretation. Additional notes of potential relevance were included in the final column of the data-charting form.

### 2.8 Synthesis of results

Data representing the year of publication, geographic coverage, whether a study was classified as academic or gray literature, specific publication types (journal article, report, etc.), and whether a study presented original research or was a review were prepared and rendered as figures. A table was prepared for the appendix containing each citation, its geographic coverage and summary of results. Due to the iterative nature of a scoping review, we deviated slightly from the registered protocol at this step to report on geographic coverage instead of population characteristics, and to provide a more contextualized summary rather than just a comment on the relationships between Indigenous languages and health reported.

We committed to a qualitative analysis of the aspects of health and wellness captured in the study summaries using NVivo software (Release 1.7.1, QSR International). LH applied inductive content analysis methods to formulate thematic categories and subcategories (Elo & Kyngäs, 2008), using a rich coding strategy that allowed for each study to be assigned to more than one category or subcategory when applicable. An additional code was assigned to studies that analyzed national survey data to track how this distinct and prevalent study type was distributed thematically. Following the first round of coding, LH and MT consulted to refine the themes, continuing this same process in discussion with other members of the research team until the final set and labels were agreed upon.

Finally, we applied vote-counting methods to tabulate the direction of the link between linguistic vitality and health reported in each study (Bushman & Wang, 2009). This method was chosen because the effect measures varied widely across the studies. Each summary was assigned a code: supportive, adverse, statistically non-significant, neutral (i.e., the authors studied an aspect of health or wellness which they did not explicitly present as positive or negative), or mixed. We considered mixed to be its own category based on an assumption that these studies were distinct from those which concluded on only one type of link. To avoid double-counting studies, original research and literature reviews were tabulated separately. The results of studies that both analyzed original research data and performed a literature review were segregated and counted once in each category for this analysis (i.e., the findings from the literature review portion were counted as one review study, and the findings from the original research portion were counted as one original research study). As the vote-counting method can only provide a crude estimate of the true existence of a link between languages and health as it weighs each study equally without accounting for critical factors such as effect size, study quality, or validity (Bushman & Wang, 2009), the goal of this aspect of the analysis was only to obtain a perspective on what types of links between languages and health were reported in the included studies.

## 3. Results

We identified over 10,000 records: 9,864 via academic databases and 298 via the gray literature search strategy and reference lists (Fig 1). After duplicates were removed we screened a total of 8,201 publications. We excluded 7,063 at the title and abstract review stage due to irrelevance (n=7,059) or inaccessibility (n=4). We reviewed 1,122 full texts, and excluded 339 relevant publications because the analyses and discussions of the links between Indigenous languages and health that they contained were too limited in depth, length, and/or specificity to satisfy our inclusion criterion of centrality. We also removed 52 that did not meet the source type inclusion criteria, 14 that were duplicates, and one that was not written in English. After careful review, 262 studies met the inclusion criteria. Seven of the studies are reported in two or more publications, so the total number of publications is 271. All references and summaries of each are available in the Supplementary Material Table S1.

**Fig 1:**
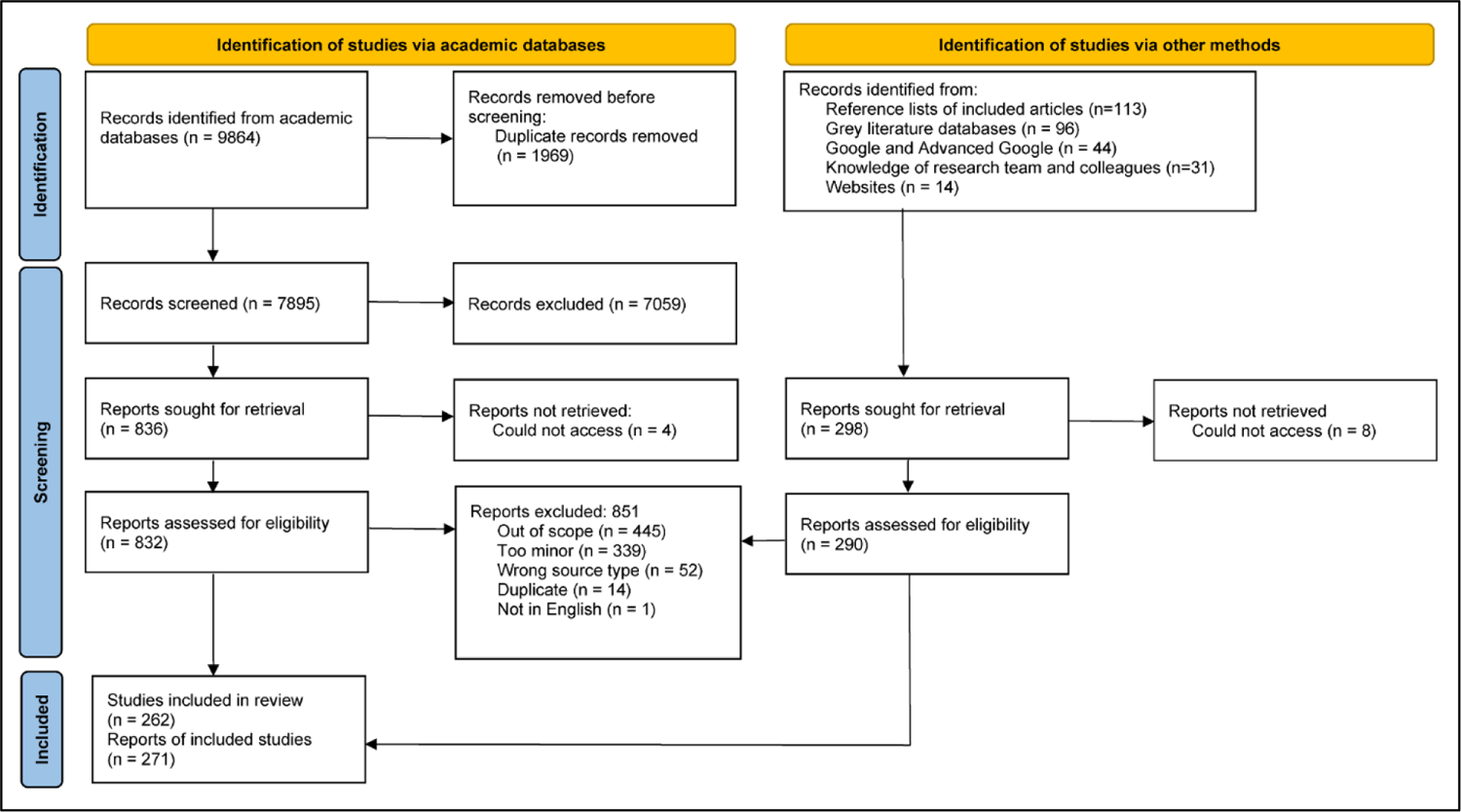
PRISMA 2020 flow diagram showing process for study inclusion. From: Page MJ, McKenzie JE, Bossuyt PM, Boutron I, Hoffmann TC, Mulrow CD, et al. The PRISMA 2020 statement: an updated guideline for reporting systematic reviews. BMJ 2021;372:n71. doi: 10.1136/bmj.n71. For more information, visit: http://www.prisma-statement.org

### 3.1 High-level overview of the included studies

We classified 84% (n=220) of the studies as original research, 13% (n=33) as literature reviews, and 3% (n=9) as mixed research/reviews (Fig 2). 70% of the total publications are academic and 30% are gray literature (Fig 3A). The most prevalent source types are academic journal articles (67%), reports and informational documents (16%), and graduate theses, dissertations, and capstone projects (10%; Fig 3B). All except one of the journal articles are peer-reviewed, and we have chosen to include this one outlier nevertheless because it is one of the most widely cited in the field of Indigenous languages and health and is published in an open access journal that offers a post-publication peer review process with reviewer comments posted which denote approval with reservations (Whalen et al., 2016). All book chapters in the final sample are published by reputable scholarly publishers, where industry standards involve some form of review before publication.

**Fig 2:**
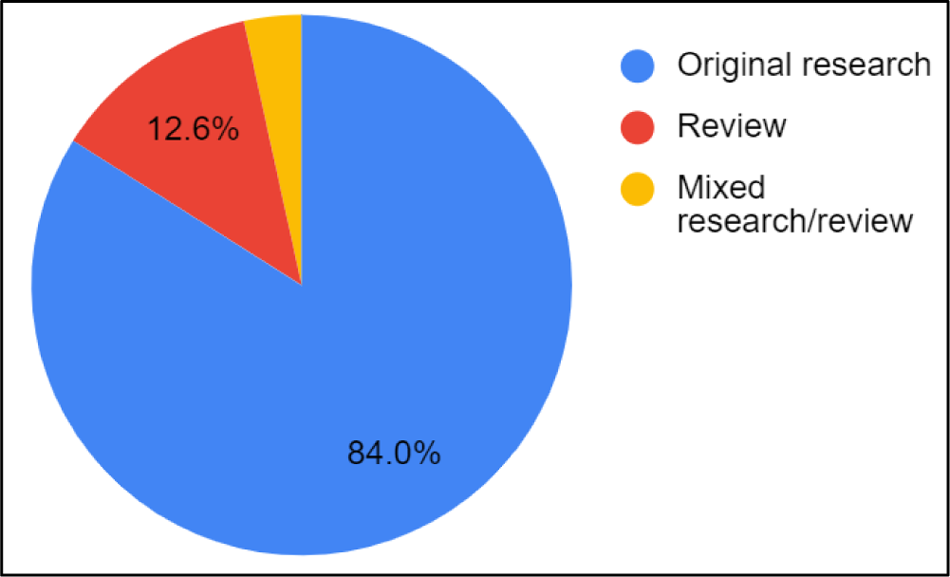
Classification of all studies into original research, review, and mixed research/review (n=262 studies).

**Fig 3:**
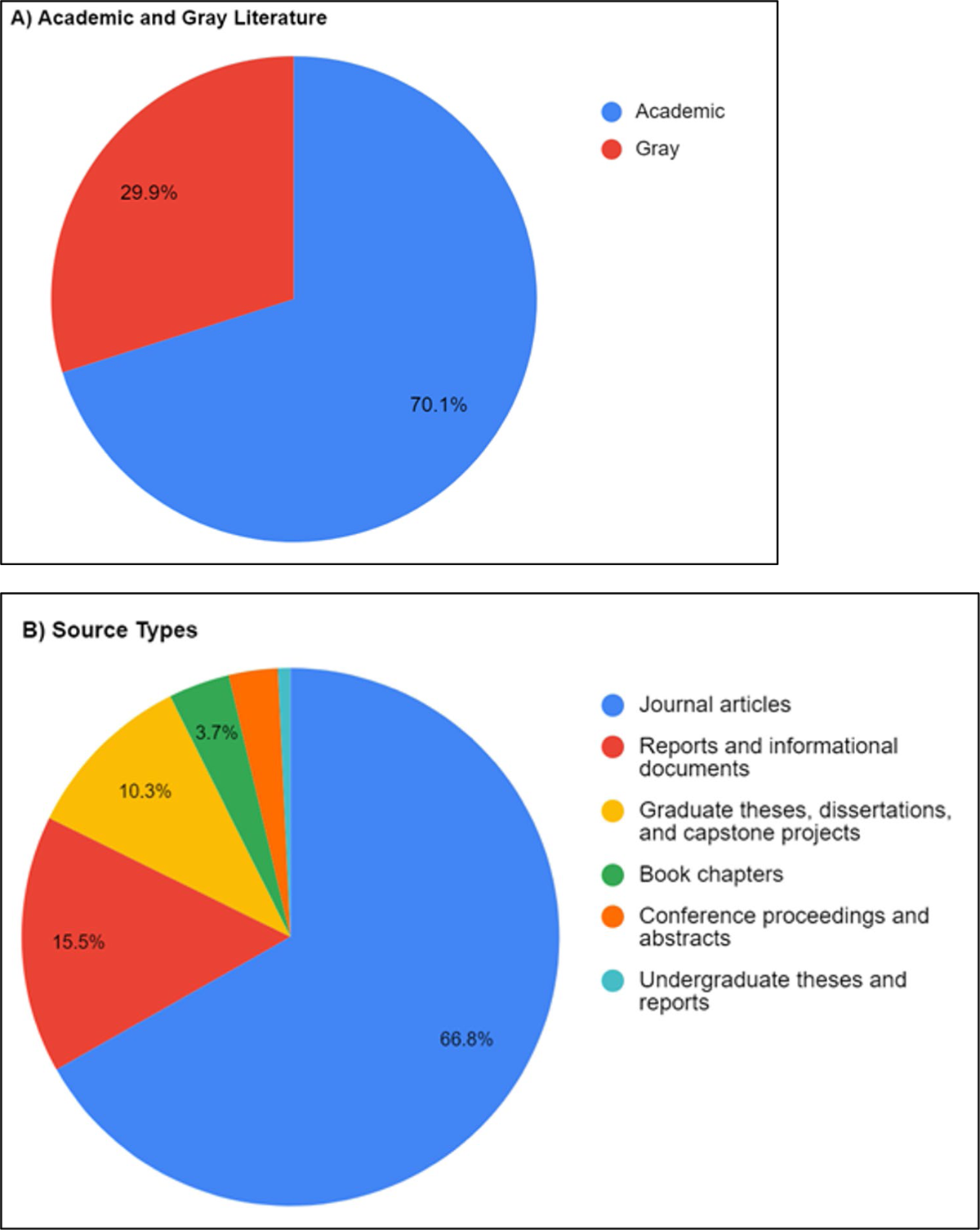
Distribution of academic and gray literature and source types in the dataset. (A) Classification of all publications into academic and gray literature. (B) Source types of all publications. Unlabeled slices are conference proceedings and abstracts 3.0%, and undergraduate theses and reports 0.7%. Sample size is n=271 publications for both charts, representing 262 studies.

Five of the included reviews surveyed aspects of the literature on the relationships between Indigenous linguistic vitality and health with similar scopes to ours reported here (Angelo et al., 2019; McIvor et al., 2009; Taff et al., 2018; van Beek, 2016; Whalen et al., 2016), one of which was published in a peer-reviewed academic journal (McIvor et al., 2009). Of additional note is a realist review from 2022 which was not included due to the date cut-off rule but will be addressed in the discussion due to its relevance (Whalen et al., 2022). That review built on a 2016 report which is included in our sample (Whalen et al., 2016), and involved searching one database and citation chaining. The review conducted by our team distinguishes itself from all prior and existing reviews on the topic of which we are aware through our use of a large spectrum of databases and search terms, application of the rigorous scoping review methodology, and our prospective registration of the study protocol.

The earliest date of publication included in our review is 1942, and at least one study has been published on the topic of languages and health every year since 1993 (Fig 4). There was an increase in publications around the turn of the millennium, and 50% of the total were published after 2013. Only 4 publications released in 2021 are captured in our scoping review as studies published after this cut-off were not included.

**Fig 4:**
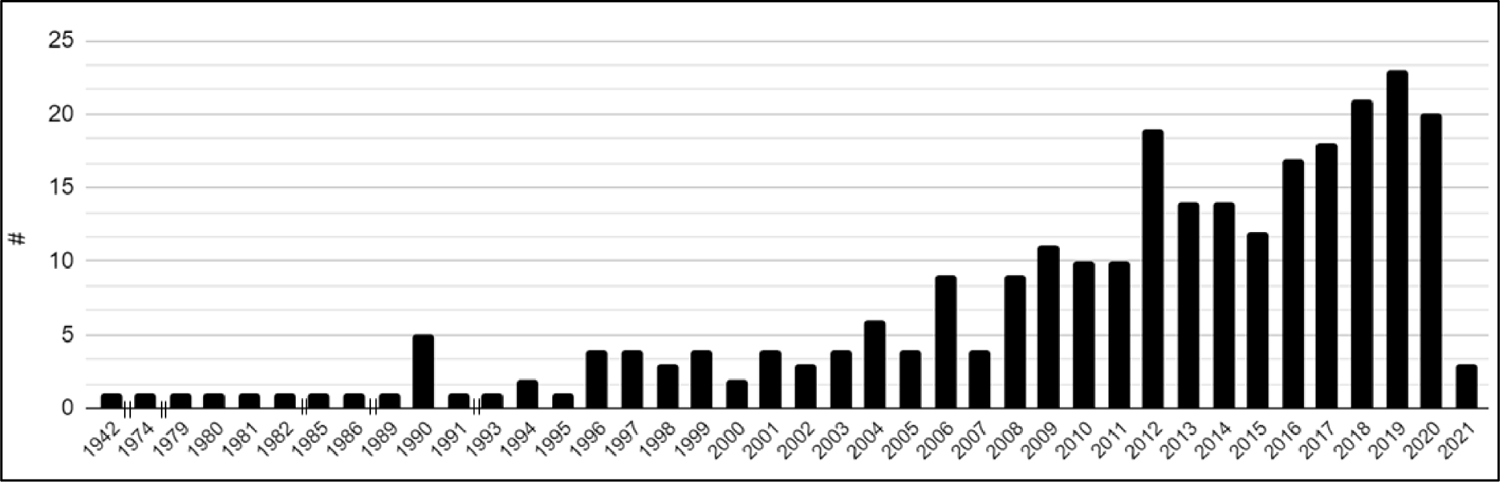
Years of publication for all reports (n=271). Double vertical lines || on the x-axis denote non-consecutive years.

The largest number of studies focus on the Canadian context, and the least on New Zealand (Fig 5A). Proportionate to the size of the Indigenous populations in each country, populations in the USA are underrepresented in the dataset and populations in Australia and Canada are overrepresented (Fig 5B) (Australian Bureau of Statistics, 2022; Statistics Canada, 2022; Stats NZ Tatauranga Aotearoa, 2022; United States Census Bureau, 2021). We encountered challenges when screening certain literature for inclusion about Pacific Islander peoples due to ambiguous usage of the population categories of Asian Americans and Pacific Islanders (AAPI), Asian/Pacific American (APA), and Asian/Pacific Islander (API). As Asian peoples are out of scope of this review and only some Pacific Islander peoples are within the scope (i.e., peoples who are Indigenous to what are now known as the U.S. Pacific Islands region and the Realm of New Zealand), we had to exclude an estimated 35 studies that are otherwise relevant to this scoping study in which the results are neither clearly nor exclusively linked to these populations.

**Fig 5:**
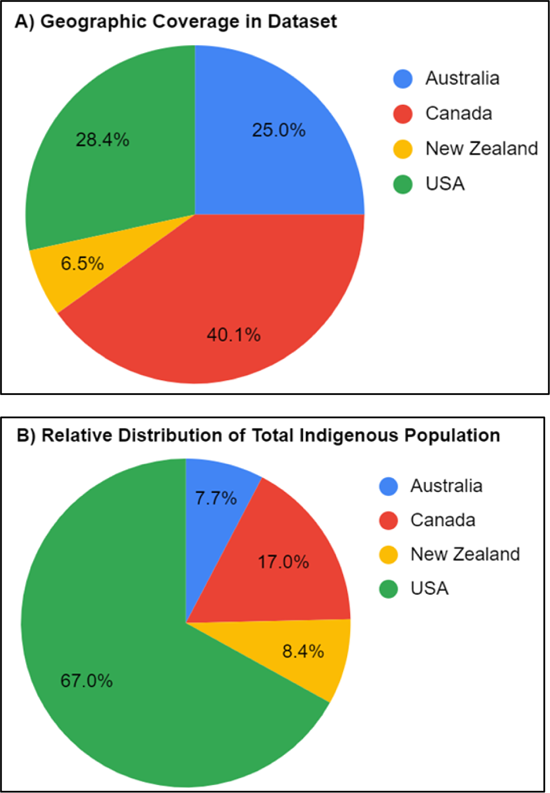
Geographic coverage in dataset with comparison to the relative distribution of the total Indigenous population. (A) Geographic coverage of studies in the dataset, with 292 data points for 262 studies since some studies looked at populations in more than one country. (B) Distribution of the aggregated population of 10,387,728 Indigenous peoples between the four countries studied, for comparison (Australian Bureau of Statistics, 2022; Statistics Canada, 2022; Stats NZ Tatauranga Aotearoa, 2022; United States Census Bureau, 2021).

The study summaries are offered in Supplementary Material Table S1. A variety of methods are applied in the studies, ranging from conventional quantitative and qualitative methods to public hearings and narrative presentations of lived experiences. Over ten percent (n=32) of the original research studies analyze quantitative data from national population surveys, most commonly the Aboriginal Peoples Survey from Canada (n=12) (Statistics Canada, 2012) and three national Australian surveys of Aboriginal and Torres Strait Islander peoples (n=9) (Australian Bureau of Statistics, n.d.).

### 3.2 Language variables in the dataset

The language variables in the studies span the spectrum of language use, proficiency, fluency, maintenance, learning, vitality, revitalization, reclamation, access to linguistically appropriate services, linguistic suppression and oppression, punishment for language use, and language loss and endangerment. There is notable variability in how language proficiency and frequency of use are measured in the dataset. For example, several studies analyzed data from the Canadian Aboriginal Peoples Survey (APS) asks, “do you speak an Aboriginal language, even if only a few words?”, which is analyzed as a binary yes/no variable (Statistics Canada, 2012), while the three national Australian surveys ask for primary language at home (Australian Bureau of Statistics, n.d.). The 1996 Canadian census queried respondents’ ability to speak a language well enough to conduct a conversation (Norris & MacCon, 2004), which Hallett and colleagues used in their landmark 2007 study to compare communities where a majority of the members responded “yes” to this census question with those where a majority answered “no” (Hallett et al., 2007). Other research teams formulated their own language variables, such as Noreen et al. (Noreen et al., 2018) who asked respondents whether they speak the Cree language exclusively at home, and Russell (2018) who indexed a range of language competencies, from “respondents who could talk about ‘almost anything’, ‘many things’ or ‘some things’ in te reo Māori […] [to] those who could only talk about ‘simple/basic things’ or ‘a few words or phrases’.”

Angelo and colleagues (2019) address this diversity in the language variable in their own review and argue that it can complicate the understanding of the links between languages and health. They advocate for greater consideration of key mediators of the relationship between languages and health including the local configuration of languages (language ecologies), contexts of use, proficiency, and type of language, and assert that “what people report as ‘speaking an Indigenous language’ may be thought of differently in different ecologies” (Angelo et al., 2019, p. 19).

### 3.3 Links between languages and health in original research studies

Of the 229 studies which present original research data, 78% (n=179) report supportive links between Indigenous linguistic vitality/revitalization and health/wellness, 10% (n=22) are mixed, 6% (n=14) are statistically non-significant, 5% (n=11) report adverse links, and 1% (n=3) are neutral (Fig 6).

**Fig 6:**
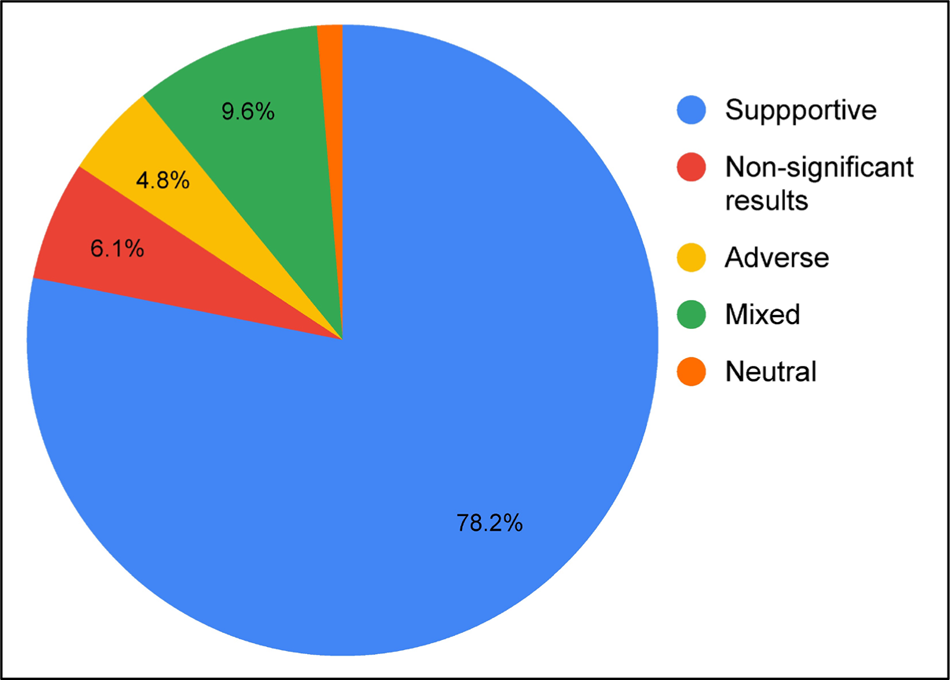
Direction of links between Indigenous languages and health reported in studies presenting original research data (n=229). The unlabeled slice is Neutral, 1.3%.

A wide range of measures of health and wellness are represented in the dataset (Table 1). Many studies examine multiple aspects of health and wellness and their interconnectedness:

> “First Nations seek to achieve whole health—physical, mental, emotional, spiritual, social, and economic well-being—through a coordinated, comprehensive approach that respects, values, and utilizes First Nations cultural knowledge, approaches, languages, and ways of knowing.” (Thunderbird Partnership Foundation & Health Canada, 2015, p. 1)

**Table 1:** Links Between Languages and Health in Original Research Studies.

| Direction of link | Aspects of health |
| --- | --- |
| Supportive (n=179) | Health care, education, and promotion outcomes (n=79)<br>Health care (n=67)<br>Patient education (n=11)<br>Health promotion (n=4)<br>Overall health, wellness, resilience, and healing (n=68)<br>Health and wellness, generally (n=47)<br>Trauma and healing (n=25)<br>Child and youth health and general development (n=2)<br>Mental, cognitive, and psychological health and development (n=51)<br>Identity (n=17)<br>Mental and emotional health and wellbeing generally (n=9)<br>Mood (n=9)<br>Suicide (n=9)<br>Self-esteem (n=8)<br>Cognitive and psychological health and development (n=4)<br>Stress (n=4)<br>Anxiety (n=2)<br>Health protective or risk factors and behaviors (n=24)<br>Social support and community connectedness or isolation (n=13)<br>Diet and movement (n=6)<br>Resilience (n=4)<br>Physical health and sickness (n=11)<br>Non-specific physical health, sickness, illness (n=6)<br>Specific illness or disease (n=4)<br>Spiritual wellbeing (n=9)<br>Substance use behaviors and addictions (n=4) |
| Non-significant findings (n=14) | Mental, cognitive, and psychological health and development (n=6)<br>Mood (n=2)<br>Cognitive and psychological health and development (n=2)<br>Health protective or risk factors and behaviors (n=4)<br>High-risk sexual behavior (n=2)<br>Physical health and sickness (n=2)<br>Overall health, wellness, resilience, and healing (n=1)<br>Substance use behaviors and addictions (n=1) |
| Adverse (n=11) | Health protective or risk factors and behaviors (n=4)<br>Cancer screening participation (n=2)<br>Substance use behaviors and addictions (n=4)<br>Overall health, wellness, resilience, and healing (n=2)<br>Physical health and sickness (n=1) |
| Neutral (n=3) | Accessing a traditional healer (n=2)<br>Substance use behaviors and addictions (n=1) |

Tabulation of the conclusions of each study presenting original research data on the direction of the link between Indigenous languages and health (n=229). Only subcategories containing more than one study are included in the table for reasons of concision, and studies reporting mixed results are not included due to complexities in interpretation. Both are instead summarized in the text.

Access to linguistically appropriate services emerged as an important dimension in relation to the links to health, encapsulating situations where Indigenous language speakers are supported to use “their first (i.e., the language they have learned as a baby) and strongest language […] to access services or information which might otherwise be detrimental if English-only” (Angelo et al., 2019, p. 7). Questions of language access are mostly covered in the context of health services, but also include studies on the benefits of bilingual public signage (Townsend, 2014), food labels (Bird et al., 2008), and prescription labels (Meisel & Kiely, 1981).

### 3.4 Supportive links in original research

#### 3.4.1 Health outcomes from linguistically appropriate health care, education, and promotion initiatives

The largest category overall contains reports that health outcomes are supported by linguistically appropriate health care, education, and promotion initiatives (n=79), accounting for almost half (44%) of all supportive studies. Almost a third of all studies in this category (n=23) focus on Aboriginal and Torres Strait Islander peoples in the Northern Territory of Australia, and particularly those who live in remote areas. Most of the studies focus specifically on health care and assessment services (n=67), while linguistically tailored patient education is a focus of eleven studies and health promotion is a focus of four. While these studies touch on a range of health conditions, brain health was a particularly salient focus, with 17 studies focusing on the need for linguistically appropriate assessment, treatment, and support for people with conditions such as dementia, traumatic brain injury, communication disorders, and in the field of speech-language pathology overall.

One study of a sample of 366 Inuit children who primarily spoke Inuktitut reports that three-quarters of the cohort were diagnostically misclassified by a commonly applied cognitive assessment scale, largely due to reasons of somewhat limited English language verbal comprehension (Wilgosh et al., 1986). In all cases, shortcomings in linguistically appropriate care are linked to lack of understanding by patients and worse outcomes such as incorrect diagnoses, low health literacy, misunderstanding of medical instructions, and feelings of alienation or not being respected. Even where interpreters are available in health care settings, many studies report that they are not consistently used. Such failings have been repeatedly identified over time and multiple calls have been made for urgent resolution:

> “Anaesthetists would not consider providing anaesthesia for anyone who spoke little or no English without the assistance of an interpreter unless it was an extreme emergency. Despite this, over a two-month period at [Royal Darwin Hospital], 29 indigenous people with little or no English underwent anaesthesia without an interpreter. This problem is unlikely to be of recent onset or confined to [Royal Darwin Hospital] alone.” (Cheng et al., 2004, p. 546)

> “If it’s a German speaking person we automatically go get that [interpreter services] straight away or find someone to do it. But if we want a Pitjantjatjara, or an Alyawarr speaking person, you know, we can use the interpreter speaking services, but we don’t.” (Artuso et al., 2013, p. 9)

> “The Committee considers it a national disgrace that an Indigenous person may face […] a serious health issue without effective interpreting support.” (House of Representatives Standing Committee on Aboriginal and Torres Strait Islander Affairs, 2012, p. 183)

The second largest category (n=68) contains studies on supportive links to overall health, wellness, resilience, and healing. 47 of these studies relate to health and wellness generally and non-specifically, including self-reported health status and quality of life. A third of the studies (n=25) examine language suppression, denigration, and loss as sources of trauma, and language teaching, learning, maintenance, and revitalization as opportunities for individuals and communities to heal, survive, and thrive (e.g., (Ball & Lewis, 2014; D. L. Brown, 2016; Cohen, 2001; Jacob et al., 2019)). Two studies report that speaking and learning Indigenous languages supports the overall health and development of children and youth (Ball, 2006; Gerlach, 2018).

#### 3.4.2 Mental, cognitive, and psychological health and development

The next largest category (n=51) speaks to how mental, cognitive, and psychological health and development are supported by linguistic vitality and challenged by language loss and suppression. Most frequently studied are identity (n=17), mental and emotional health and wellbeing generally (n=9), aspects of mood including feelings of happiness or depression (n=9), suicide rates (n=9), and self-esteem (n=8). Four studies report links between learning, speaking, or revitalizing Indigenous languages and better cognitive and psychological health throughout the lifespan (Counceller, 2011; Eckhart, 1983; Jacklin & Warry, 2010; Oldfield, 2016). The six remaining studies focus on stress (Cunningham & Paradies, 2012; England Aytes, 2015; Taff et al., 2018; Wilson, 1997) and anxiety (Brougham & Haar, 2013; Whitbeck et al., 2004). Overall, the community-engaged studies in this category tend to present the links between languages and mental health in a way that is holistic and within the context of multiple supportive factors, as described in a joint report by the Thunderbird Partnership Foundation and Health Canada:

> “Mental wellness is supported by factors such as culture, language, Elders, families, and creation. It is necessary for healthy individual, family, and community life.” (2015, p. 1)

#### 3.4.3 Other supportive categories

Twenty-four studies focus on preventative and risk factors. Over half of these studies (n=13) relate to social support and community connectedness, reporting that language use, revitalization, and preservation strengthen communities while language loss can challenge these connections and worsen health and wellbeing. One study presents this from the inverse perspective, reporting that the social isolation of Indigenous seniors can negatively impact linguistic vitality because of their important role in language transmission and preservation (Employment and Social Development Canada, 2018). Six studies report links with improvements in dietary and movement patterns, including increased physical fitness and healthy weight loss by students in a language revitalization program (Erasmus, 2019), greater participation in traditional physical activities and consumption of traditional foods (Hilgendorf et al., 2019; Janssen et al., 2014; Lévesque et al., 2015; Redwood et al., 2008), and higher self-efficacy in healthy food preparation (Mercille et al., 2012). Four studies report on higher overall resilience to threats to health and wellbeing (Liebenberg et al., 2015; Pearce et al., 2015; Phillips, 2010; Thunderbird Partnership Foundation & Health Canada, 2015). Both of the final two studies in this category analyze data from a 1993 survey with Hopi women and report associations between language use, likelihood of practicing traditional behaviors to keep healthy (Coe et al., 2004), and participation in routine breast cancer screening (Giuliano A et al., 1998).

Eleven studies report supportive effects on physical health related to language use. Six of these studies focus on physical health, sickness, and illness in a general, non-specific sense (Bach, 2016; Basso, 1990; H. J. Brown et al., 2012; Fitzgerald, 2017; Hilgendorf et al., 2019; Running Bear et al., 2018). Four focus on specific illnesses and diseases—three on diabetes prevalence (Oster et al., 2014), progression (Teng et al., 2019), and related hospitalizations (C. King et al., 2018), and one on asthma rates (Cunningham, 2010). The final study reports benefits to childhood oral health (Ball et al., 2013).

The literature unequivocally supports the positive effects of linguistic vitality on spiritual wellbeing in the nine studies that assess it, as encapsulated in the following quotes:

> “What must be remembered is that language is not simply a tool for everyday communication, but through recording of stories, songs, legends, poetry and lore, holds the key to a people’s history and opens the door to cultural and spiritual understanding.” (Wilson, 1997, p. 259)

> “There are many spiritual and cultural precepts Hawaiians once knew and are rediscovering through the reemergence of the Hawaiian language.” (Bach, 2016, p. 82)

> “Our journey into the healing potential of language is an inexorable course through spirituality.” (Iwama et al., 2009, p. 8)

Four studies report links between linguistic vitality and safer, less frequent substance use. Brass (Brass, 2013) reports that Aboriginal respondents who used their language at home were less likely to consume more than four alcoholic drinks per day when compared to people who didn’t use their language at home. Healey and Meadows report that the emotional pain of language loss can lead people to “tur[n] to addictive substances to cope” (2008, p. 30), a finding also echoed in a publication by Health Canada (Thunderbird Partnership Foundation & Health Canada, 2015). An excerpt from a consultation by Taff and colleagues provides an account of a language learner’s path to sobriety:

> “[…] I was at a ceremony to do with language where we were taught that alcohol does not serve. I made a commitment for four years to not drink because I wanted to be a good language learner. I wanted to be able to look at my elder and speak to him and not feel ashamed of myself. I’m thirty-three now and I haven’t drunk since I was twenty.” (2018, p. 6)

### 3.5 Non-significant results in original research

14 studies report statistically non-significant results from the analyses which they conducted on the links between languages and health. All these studies were observational, involving the administration of surveys and quantitative analyses. The six studies in the mental, cognitive, and psychological health and development category focus on mood (Beckstein, 2015; B. Davis, 2012), cognitive development (Cummings, 1998; Goyal et al., 2018), history of diagnosis of an anxiety disorder (Nasreen et al., 2018), overall mental health as measured by a range of component factors and a history of having considered suicide (Guevremont et al., 2016). Four studies look at health protective or risk factors and behaviors, specifically cancer screening rates and knowledge (Gonzales et al., 2012), high-risk sexual behavior (Myers et al., 1999), survival sex work (Sharma et al., 2020), and healthy movement patterns, measured as leisure-time physical activity and walking as a means of active transportation (Ryan et al., 2018). The studies on physical health and sickness focus on the prevalence of morbid obesity (Hodge et al., 2011) and history of hysterectomy (Zhang et al., 2005). One study examines correlations to rates of heavy drinking (Ryan, Cooke, et al., 2016), and one looks at self-rated health (Eliassen et al., 2012).

### 3.6 Adverse links in original research

11 studies report adverse links between measures of language and health, all of which use quantitative epidemiological methods to analyze decontextualized national survey data.

Four of the studies report links to health risk factors and behaviors. Two of these report links between use of an Indigenous language at home and a lower likelihood of lifetime participation in screening tests for colorectal and breast cancers (Perdue et al., 2011; Tapia et al., 2019). Redwood and colleagues (2012) report that participants who spoke an Alaska Native or American Indian language at home were more likely to be exposed to occupational and environmental hazards such as to asbestos and pesticides. And Temple and Russell (2018) report that older Aboriginal and Torres Strait Islander people who speak their Indigenous language were more likely to experience food insecurity, particularly those who live in remote areas.

All four studies within the substance use behaviors and addictions category are analyses of data from the 2006 and 2012 Aboriginal Peoples Surveys in Canada and report associations between Aboriginal language use and smoking rates among youth and adults (Ryan et al., 2015, 2017; Ryan, Leatherdale, et al., 2016; Van Bewer & Woodgate, 2017). Three of the four studies are from the same research team.

Two studies report adverse links to measures of overall health, wellness, resilience, and healing, and both are based on analyses of Australian national survey data on Aboriginal and Torres Strait Islander peoples. The first focuses on five-year-old children in the Northern Territory and reports a higher likelihood of being assessed as developmentally vulnerable if they spoke English as a second language compared to English as a first language, independent of other perinatal and socio-demographic factors (Guthridge et al., 2016). The second study identifies an association between not speaking English as a primary language at home and self-reporting fair or poor health when compared to people who primarily spoke English at home, even after accounting for other factors (Sibthorpe et al., 2001).

The final study is in the category of physical health and sickness and reports that COVID-19 rates were lower in Oklahoma reservations with higher percentages of English language-only households (Rodriguez-Lonebear et al., 2020).

### 3.7 Neutral links in original research

Three studies report links to health-related behaviors which we have categorized as neutral, as the authors of the respective studies did not explicitly specify whether these behaviors support or adversely impact health outcomes, i.e., they took a neutral approach to the subject.

Two of these studies report links between Indigenous language use and the likelihood of accessing and believing in the efficacy of traditional Indigenous medicine healers (Henderson, 2009; Waldram, 1990a), while the third reports a link between speaking an Indigenous language at home and the likelihood of using smokeless tobacco products as compared to cigarettes or cigars (Redwood et al., 2010). The studies in this category applied quantitative analytic methods to retrospective national survey data (Henderson, 2009), to a large-scale health survey (Redwood et al., 2010), and to interviews (Waldram, 1990a).

### 3.8 Mixed findings in original research

22 studies report some combination of supportive, adverse, non-significant and/or neutral links between language and health. Eight of the mixed studies focus on health risk and protective factors, seven on physical health, six on mental or cognitive health, two on substance use, one on spiritual wellbeing, and one on overall health.

Twelve of these studies report a mix of findings across the multiple aspects of language and/or health which they assessed. For example, Dockery (2011) reports a supportive link between speaking an Aboriginal or Torres Strait Islander language and lower rates of alcohol use, but mixed links to various measures of mental health. Jenni and colleagues (Jenni et al., 2017) report that participation in a language learning program supported the students’ overall wellbeing, substance use dependency recovery, and spiritual wellbeing, but that this same participation had a mixed effect on physical health as some learners experienced exhaustion and fatigue.

Nine of the mixed results studies report on links that differ by gender, geographical location, and/or the ages of participants. For example, Cooke and colleagues (2013) report associations between Indigenous language knowledge and obesity among Métis boys aged 6-10 and girls aged 11-14 in Canada, but not among girls aged 6-10 or boys aged 11-14. Two studies from each Australia and Canada report that the positive psychological effects of Indigenous language fluency were only significant for populations in their sample who lived in rural and remote areas (Dockery, 2011; Hossain & Lamb, 2019). The author of the Australian study hypothesized that their finding reflected the stress of “coexistence within both a traditional, minority culture and a mainstream culture” for Indigenous language speakers living in non-remote areas (Dockery, 2011, p. 19), while the authors of the Canadian study suggested that other unmeasured variables which influence wellbeing may explain their finding.

Two studies report different links to health depending on whether the population is bilingual or mainly/exclusively speaks their Indigenous language. We coded these studies in the mixed category since the interpretation of these findings in the context of our research question is not straightforward. Duncan and colleagues (2014) report higher levels of physical activity among the bilingual speakers in their Northern Plains American Indian sample compared those who exclusively spoke their Indigenous language. Meanwhile, Zienczuk and Egeland (2012) report that the proportion of adults who were overweight or obese was higher among the populations who either spoke only English or both an Inuit language and English at home compared to those who primarily spoke an Inuit language. Given that Zienczuk and Egeland report a gradient where odds of an at-risk body mass index increases if both an Inuit language and English are spoken at home and increases further if only English is spoken at home, the frequency of use of English as an additional variable complicates the interpretation of this study in the context of understanding the direction of links between Indigenous language use and physical health.

### 3.9 Links between languages and health in literature reviews

Of the 42 studies which review previous literature, 41 report only supportive links between languages and health (Table 2). The only exception is a literature review that takes a neutral approach—Waldram (1990b) reports a link between speaking an Indigenous language and seeking the services of a traditional Indigenous healer and/or believing in the superiority of traditional medicines for some medical problems. The author does not specify or speculate on how these beliefs and behaviors may be associated with health or wellness outcomes.

**Table 2:** Links Between Languages and Health in Literature Reviews.

| Direction of link | Aspects of health |
| --- | --- |
| Supportive (n=41) | Mental, cognitive, and psychological health and development (n=22)<br>Health care, assessment, access, education, and promotion (n=17)<br>Overall health, wellness, resilience, and healing (n=17)<br>Health protective or risk factors and behaviors (n=9)<br>Physical health and sickness (n=9)<br>Spiritual wellbeing (n=4)<br>Substance use behaviors and addictions (n=3) |
| Neutral (n=1) | Accessing a traditional healer (n=1) |

Tabulation of the conclusions of literature review studies on direction of link between languages and health (n=42).

Mental, cognitive, and psychological health are the primary focus of the literature reviews (n=22), followed by health care, assessment, access, education, and promotion (n=17):

> “A refusal to take Aboriginal languages seriously not only results directly in less than optimal medical outcomes, but also in mistrust and disengagement with the health sector and non-compliance with treatment regimens.” (Amery, 2017, p. 15)

Overall health and wellness is an equally large category (n=17), as is described in an Australian social justice report:

> “Where languages are eroded and lost, so too is the cultural knowledge. This in turn has potential to impact on the health and well-being of Indigenous peoples.” (Aboriginal and Torres Strait Islander Social Justice Commissioner, 2009, p. 58)

The remaining aspects of health supportively linked to language in the literature reviews are health protective and risk factors (n=9), physical health (n=9), spirituality (n=4) and safe substance use (n=3). Exemplifying and summarizing the strong trend towards supportive links is this quote from an Australian report:

> “It is abundantly clear that our cultural health and wellbeing rests on the linkages we have and maintain with mother tongue and mother culture, and it is abundantly clear that when these linkages are threatened our anguish over loss causes an intensification of the holistic lifelong learning process which then cycles round and causes serious ill-health. On this basis alone I say to myself—do we really need further evidence to demonstrate that there is a direct causal link between a healthy mother tongue and mother culture and a healthy community? If our children are dying through lack of culture or too much culture all at once, we surely must see that something is dreadfully wrong.” (Williams, 2011, p. 64)

## 4. Discussion

In this scoping review of 262 studies about the links between linguistic vitality and the health and wellness of Indigenous peoples in Anglo settler-colonial societies, our research team collated and assessed a diverse body of academic and gray literature. It has become apparent that while a limited set of studies tend to be cited on this topic, it is a well-researched field where a wide variety of measures of language, health, and community contexts have been examined using a range of research methods.

A majority of the studies that were selected for inclusion reported supportive links between languages and health throughout the lifespan. The use, proficiency, fluency, maintenance, vitality, learning, revitalization, and reclamation of Indigenous languages tended to be positively associated with health and wellness. Meanwhile, the diminishment, denigration and destruction of Indigenous languages were associated with worse health and wellness.

The pervasiveness and urgency of cases in which there was a documented lack of access to linguistically appropriate health care was a highly prevalent theme. Alongside this were studies demonstrating how providing health care and education in Indigenous languages, rather than English, can improve the likelihood of positive outcomes. As a key health ethics issue arising from this study, language is relevant to health professionals’ commitments to provide culturally appropriate, safe, and effective health care (Curtis et al., 2019; Miles, 2004) and of public health to enact justice and respect for all persons (Public Health Agency of Canada, 2021). The large number of studies focusing on the need for linguistically appropriate care for people with conditions affecting the mind and brain has implications for neuroethics where recent research has highlighted the limitations of reductionist approaches to neurology and psychiatry and the need to meaningfully incorporate Indigenous ways of knowing and being into research and care (Harding, 2022). These findings also add evidence that language oppression is necropolitical, i.e. that “language oppression kills,” (Roche, 2022, p. 34), and highlight the centrality of language revitalization in Indigenous resurgence (Simpson, 2017).

A comparatively small portion of the 262 studies reported adverse links between linguistic vitality and health. None of the included literature reviews identified any adverse links. However, the 2022 realist review on Indigenous languages and health which we introduced in section 3.1 reported that 31 of their 180 included studies suggest negative impacts on health (Whalen et al., 2022).

Regarding the specific studies in our sample that reported adverse links, there are some limitations to using many of these studies to advance an understanding of potential adverse links. The authors of most of these articles noted concerns around confounding variables and potentially irrelevant measures of health more frequently than did the authors of the studies reporting supportive links.

Specifically, six of the eleven studies examined aspects of health that are known to be correlated with socioeconomic status and structural barriers: smoking, exposure to occupational and environmental hazards, and food insecurity (Hiscock et al., 2012; Moses et al., 1993; Shafiee et al., 2022). The authors of the two studies that report links between Indigenous language use and lower cancer screening participation speculate on possible confounders, suggesting that the results may reflect an avoidance of the medical system based on experiences of systemic racism (Perdue et al., 2011) or that “language itself is not the risk factor, but rather a marker for access to services” (Schumacher et al., 2008, p. 735). Sibthorpe and colleagues (2001) postulate that the global self-assessed health question on the population-level survey they analyzed was not culturally valid because the finding of lower overall health among Indigenous language speakers was not consistent with mortality or morbidity data.

Another consideration surrounding the utility of some data in our sample arises when considering the subset of studies which used quantitative, epidemiological methods to analyze decontextualized national survey data. As we have noted, all the studies that reported only adverse links between language and health assumed this same methodological approach.

Through an editorial written in response to one of the studies in our sample that reported a correlation between smoking and language use (Ryan et al., 2015), King (2015) makes the point that the research design of the study lacked community contextualization or validation. King recommends that even though secondary analyses of de-identified data do not require ethics board review, they should follow the same ethical criteria as primary research in Canada of “engag[ing] experts with the requisite contextual knowledge – Métis community knowledge holders, in this case – to guide the data interpretation” (2015, p. e457).

A related epistemological consideration arose from the wide range of measures of health and wellbeing reported in the publications screened for inclusion, and the challenges of defining eligibility criteria that were both inclusive and could meet the public health policy advocacy objectives of this review. The English term “health” can be restrictive and government measures such as attendance rates to cancer screening programs or prevalence of tobacco use can lack relevance in some contexts (Boddington & Raisanen, 2009). Two-Eyed Seeing is a useful concept for approaching these differences from Mi’kmaq Elder Albert Marshall, who authored one of the studies included in this review (Iwama et al., 2009):

> “[…] the gift of multiple perspectives […] learning to see from one eye with the strengths of Indigenous knowledges and ways of knowing, and from the other eye with the strengths of Western knowledges and ways of knowing, and […] using both these eyes together, for the benefit of all,” (Bartlett et al., 2012, p. 355)

The uneven distribution of publications across the four countries studied may indicate the need for more resourcing of and focus on Indigenous language and health research and policy work in the USA, and that Australia and Canada are current leaders in this field. However, the challenge we noted around how Pacific Islanders are grouped together with other Indigenous and non-Indigenous populations in research and census data may also have contributed to the underrepresentation of communities in the USA.

To provide a brief comment on the publications since our cut-off date in 2021, we ran our search again in MEDLINE (Ovid) on June 21, 2024. We limited the results to those with language*, linguistic*, revitali*, or recla* in the title, which yielded 135 results. LH reviewed each title and abstract and noted two relevant articles, including the 2022 realist review by Whalen and colleagues which we have already commented on here (Whalen et al., 2022). The other article reports on data from a community-based participatory research project involving the administration of surveys to 191 Anishnaabe adults (Gonzalez, 2021). Quantitative analyses suggested that spiritual connectedness through prayer is a mediator of the relationship between Indigenous language use in the home and positive mental health.

The strengths of our current study include comprehensiveness, analytical rigor, and multidisciplinarity. Making use of a large number of databases spanning different academic disciplines and including gray literature allowed us to have a broad scope and reduce selection bias. The use of a scoping review methodology that follows many of the established best practice guidelines, including the pre-registration of a research protocol, affords scientific rigor, replicability, and transparency to the work. An extensive supplement was prepared with summaries of each study to contextualize findings and support further research and exploration of the study data (Supplementary Material Table S1). And finally, the multidisciplinary composition of our research group has helped strengthen our process, engage with Indigenous understandings, and contextualize our findings.

### 4.1 Limitations

There are several limitations of our application of the scoping review method. Firstly, given that a scoping review highlights commonalities and generalizations, the ability to provide in-depth analyses of content that speak to the specificity of Indigenous communities, languages, and health conditions is necessarily constrained and somewhat limited. This has the additional consequence that many of our findings are quite broad, and we are not in a position to attend to how they will play out on the ground. Secondly, while we went to considerable lengths to include all relevant publications, the underreporting of negative or inconclusive findings in academic studies remains a known source of publication bias affecting literature reviews (Mlinarić et al., 2017). Thirdly, only one researcher charted the data due to our resource limitations.

The studies we captured were limited by the definitions of languages and health that we formulated. Our decision to focus on Anglo-settler colonial contexts, and therefore only papers written in English, was also based on our team’s capacity and resources. Further exploration into French, Spanish, and other settler colonial contexts will increase the sample size of literature and diversity of the understandings presented on the links between Indigenous languages and health.

The search terms we used for Indigenous communities were limited to established umbrella terms (Indigenous, Aboriginal, First Nations, etc.) because at the time of conducting the search there was a shortage of available resources that offered greater specificity (Harding, Marra, et al., 2021). This limitation has since been addressed for future literature reviews through the expansion of a set of search templates first created by the University of Alberta Library in 2021 which now provide highly specific search terms pertaining to communities in all four countries (University of Alberta Library, 2023).

New and relevant studies have been published since we conducted our search in early 2021, including but not limited to the two which appeared in our 2024 abbreviated search of MEDLINE. Therefore, we are limited in our ability to comment on new data since 2021.

There are also limitations to the analytic techniques we applied. While the content analysis we conducted was relatively straightforward with minimal interpretation required, there is always potential for unconscious bias to impact findings; for example our beliefs in the value of linguistic vitality and our positions within generally liberal academic institutions could bias findings towards supportive links between languages and health (Chambers et al., 2018). Finally, the vote-counting method offers only a crude estimate of the existence of a link between languages and health and only captures statistically significant results (Bushman & Wang, 2009).

## 5. Conclusions and Recommendations

The above limitations and constraints aside, the results of this scoping review suggest that language is a determinant of Indigenous peoples’ health in Anglo-settler states. Actions should be prioritized which ensure that the healing effects of language can be harnessed, and that the significant and measurable human costs of linguistic oppression are understood and redressed.

We conclude our contribution with four recommendations relating to future research, community-based language revitalization work, health care, and post-secondary curricula (Table 3).

**Table 3:** Summary of recommendations.

| Recommendation | Responsible Parties |
| --- | --- |
| Create funding calls for community-engaged, Indigenous led, contextually specific research aiming to maximize and support the health benefits of languages. | Federal and regional research institutes for health and languages, and organizations with special interest in Indigenous languages and health. |
| Provide sustainable, long-term funding for programs that support linguistic vitality within Indigenous communities. | Federal and regional governments, and other organizations with special interest in Indigenous languages and health. |
| Ensure that health care and promotion are linguistically appropriate and accessible to all, irrespective of linguistic background. | Health care services administrators and health care bodies. |
| Develop courses and appropriate pedagogy on linguistic vitality and health. | Post-secondary institutions. |

Summary of recommendations arising from the scoping review results.

The next step for research on the health benefits of Indigenous languages would benefit from a focus on specificity, delving deeper into the various nuances, confounders, mediators, and other critical factors which arise in different contexts to influence this relationship. The references and summaries provided in the Supplementary Material Table S1 can be leveraged to support this. A systematic review with targeted, community-developed research questions and culturally appropriate evidence critical appraisal tools (Harfield et al., 2020) may be another productive direction. However, this research runs the risk of over-intellectualizing a human experience through a Western scientific lens, particularly given that this review revealed that most studies that engage deeply with communities have already identified supportive links and the presence of multiple holistic supportive factors. Guidance from King (2015), Williams (2011), Puebla (2014) and others remind researchers in this field to critically appraise the context and meaningfulness of any study findings which deviate from communities’ own understandings. Indigenous leadership and sustained, reciprocal community engagement following ethical research principles and using meaningful methods are therefore critical to ensuring the relevance, actionability, specificity, and ethics of such research (Hayward et al., 2021; Kovach, 2021; Smith, 2021; Thurber et al., 2020).

Targeted funding, which could be administered by federal institutes for health research and/or Indigenous languages, or state/province level resources, would advance the research. An example of a collaborative and systematic research project which received government funding is the ongoing study by Sivak and colleagues with the Barngarla community in Australia into the links between community-based language reclamation and mental health and wellbeing (2019).

By identifying, collating, and analyzing the wealth of existing evidence in this area, we hope that this scoping review can be harnessed in service of a more robust, consistent, and tangible response to Indigenous communities’ calls for sustainable funding for initiatives which seek to maintain, reclaim, and/or revitalize their languages and support language learners.

Language work is vital to the wellness of communities, and forcing community language workers to repeatedly apply for piecemeal, unpredictable, and short-term funding with no promise of long-term, multi-year support is neither appropriate nor ethical. Given the silos in which central government research funding streams tend to operate and the inherently interdisciplinary nature of the interaction between health and language, it may be cost-effective and prudent to allocate some strategic funds from health budgets towards supporting linguistic vitality to augment other existing funding sources.

The health field and health workers must act to better incorporate diverse languages into health and healing initiatives, and to ensure that all health care is linguistically appropriate and that language barriers are reduced. Special priority should be put towards ensuring that Indigenous patients’ linguistic needs are met, and that no patient experiences poor health care on account of a lack of professional interpreting support. The focus on language in health should continue to be linked closely with newer and positive developments in culturally safe care, culture as medicine, and land-based healing.

Our study findings are also actionable in a broader societal context as part of wider movements working towards Indigenous language and culture revitalization that are focused on renewed relationships with Indigenous peoples. Understanding that Indigenous languages are integral to the holistic wellbeing of a community and not just a practical tool of communication which can be replaced without consequence with English, French or Spanish is part of respecting and valuing the knowledge traditions of Indigenous peoples and their cultures. This understanding is shared by many non-Indigenous minority language communities as well, and as such can be a place for allyship and collaborative action. The references collated in this review provide sufficient content for a full post-secondary course to disseminate and advance understandings of the links between Indigenous linguistic vitality and health, which could be developed and delivered by a department of Indigenous languages, Indigenous studies, public health, or incorporated into existing courses in a number of disciplines.

In conclusion, it is time for the links between Indigenous linguistic vitality, health, and wellness to be taken seriously and for this understanding to be enacted through tangible support for Indigenous communities. As we progress through the United Nations Decade of Indigenous Languages, we are better positioned than ever to be part of this positive transformation.

## Data Availability

All data produced in the present study are available upon reasonable request to the authors

## Acknowledgments

We acknowledge Judy Illes, Sarah Dupont, and other colleagues and students who supported and enabled various stages of this project. The University of British Columbia Point Grey campus where many of the authors work is located on the traditional, ancestral, and unceded homelands of the hən̓ q̓ əmin̓ əm̓ -speaking xʷməθkʷə^y̓^ əm (Musqueam) people.

## Appendix Titles

**Appendix A:** Indigenous Linguistic Vitality and Health: A Scoping Review Protocol

**Appendix B:** Preferred Reporting Items for Systematic reviews and Meta-Analyses extension for Scoping Reviews (PRISMA-ScR) Checklist

**Appendix C:** Academic Database Search Strategy

**Appendix D:** Gray Literature Search Strategy

**Appendix E:** Data Extraction Chart

## Supplementary Material

**Table S1: Summaries of Included Studies on Indigenous Languages and Health.** Citations, geographic coverage, and a summary of each study is provided in tabular format.

