## Appendix A for "Linguistic vitality improves health and wellbeing in Indigenous communities: a scoping review"

### Abstract

**Objective:** The objective of this scoping review is to document and assess the extent and scope of literature on the relationships between Indigenous linguistic vitality/revitalization and health in Indigenous populations in anglophone, settler-colonial societies.

**Introduction:** Indigenous communities have long known about the importance of their languages for wellbeing, but this topic has only recently received attention in research and policy. As an emerging, heterogeneous and interdisciplinary field of work that has not yet been comprehensively reviewed, this scoping review will fill important gaps and guide public health and policy recommendations.

**Inclusion criteria:** Academic and grey literature written in English will be included that describes a connection between Indigenous linguistic vitality/revitalization and health/wellness in one of the four anglophone, settler-colonial states: Canada, the United States of America, New Zealand, or Australia.

**Methods:** The proposed scoping review will be conducted in accordance with the JBI methodology for scoping reviews (Peters et al., 2020). Databases to be searched include MEDLINE (Ovid), Bibliography of Native North Americans (EBSCO), Australian Education Index (ProQuest), and Linguistics and Language Behavior Abstracts (LLBA: ProQuest). Sources of unpublished studies/grey literature to be searched include desLibris, PsycEXTRA, Native Health Database, iPortal, and Google Scholar. Results will be presented in charts, tables and narrative formats.

### Introduction

British Columbia, Canada, where the research team is based, is home to 34 distinct Indigenous languages. These languages have been under great stress from the violence of ongoing settler colonialism, like many Indigenous languages across the four anglo-settler states of Canada, the United States of America, New Zealand and Australia. Indigenous language revitalization and reclamation initiatives are acts of resistance and resilience with important impacts on community wellness. These initiatives are multidisciplinary, heterogeneous, and innovative, and both learn from and inform initiatives by other communities around the world who face threats to linguistic diversity (Pine & Turin, 2017).

On an international scale, the United Nations has taken several initiatives to support the vitality of Indigenous languages. The 2007 United Nations Declaration of Rights of Indigenous Peoples (UNDRIP) included a section about the right to Indigenous languages, and was adopted despite votes against it by each of the four anglo-settler states (United Nations General Assembly, 2007). In 2016, the UN General Assembly adopted a resolution proclaiming 2019 as the Year of Indigenous Languages, and then built on this by declaring 2022-2032 as the Decade of Indigenous Languages (United Nations, 2018; UNESCO, 2020). While all promising steps in the right direction, the true impacts of these large-scale initiatives on the vitality of Indigenous languages are still unfolding. Many Indigenous language reclamation initiatives continue to be challenged by a lack of funding and supportive policies, and their importance for both community and individual health and wellbeing is often underappreciated.

In 2007, a groundbreaking research study by Hallet, Chandler & Lalonde showed a negative correlation between the vitality of Indigenous languages and youth suicide rates in BC First Nations communities (Hallet, Chandler & Lalonde, 2007). The study has since been heavily referenced for advocacy purposes and in subsequent research, including by language revitalization organizations and by Canada’s Prime Minister Justin Trudeau in his pledges to implement the Indigenous Languages Act (The Canadian Press, 2016).

Based on the important impact that Hallet and colleagues’ study has had on policy, awareness, and direct actions, our research team conceptualized the current project with the intention that systematically identifying and describing additional literature that reports on the various relationships between linguistic vitality/revitalization and health could have significant direct benefit. As Indigenous communities already know about the relationships between linguistic vitality and health, the fact that scientists are just learning this is in many ways irrelevant, but could be used to help advance the case for where resources need to be allocated by Non-Governmental Organizations (NGOs), Intergovernmental Organizations (IGOs), and public health, policy, and government bodies. As an emerging, heterogeneous and interdisciplinary field of work that has not yet been comprehensively reviewed, it is well suited to the systematic, broad approach of a scoping review.

We are aware of two previous reviews on this topic. The first may be classified as a narrative review, and was conducted by Whalen, Moss and Baldwin and never officially published (Whalen, Moss & Baldwin, 2016). The authors reviewed a selection of Indigenous language programs alongside research that has found correlations between health improvement and language revitalization to advocate for increased support for language revitalization initiatives. They reported relationships between Indigenous languages and reductions in youth suicide, diabetes, rates of “poor health”, and a wide range of behavioural risk factors. They also found relationships between Indigenous languages and being classified in a “good” wellness group, practicing traditional health promoting behaviours, and higher high school graduation rates. The manuscript was submitted for publication to the open access research platform *F1000Research* in 2016, and was approved with reservations by two reviewers. Both the manuscript and the reviewers’ comments are openly available on the research platform. The main comment from the reviewers was that they wanted the manuscript to include specific information about *how* Indigenous languages may improve health. Despite that the authors did not openly respond to the reviewers’ comments or revise the manuscript, the manuscript has been cited a number of times in its un-published format and is recognized as an important contribution to the field.

The second review we are aware of was an undergraduate thesis by van Beek at the University of British Columbia, completed through a practicum with the First Peoples’ Cultural Council (van Beek, 2016). van Beek conducted a narrative review of the body of work regarding health and language, searching for academic and grey literature using terms about Indigenous populations, health, wellness, and language learning and speaking. The review reported on 55 pieces of literature through an annotated bibliography and discussion of five emergent themes: indicators of health and wellbeing, academic success, Indigenous wellness concepts, identity, and resilience. van Beek reported associations between Indigenous languages and lower rates of diabetes, suicide and HIV. The review was not academically published, but is available on the First Peoples’ Cultural Council website as grey literature and has been cited by at least two academic publications.

A preliminary search for additional scoping and systematic reviews on the topic was conducted on January 21, 2021 in JBI Evidence Synthesis, Cochrane Database of Systematic Reviews, Google Scholar and Epistemonikos, and did not yield any additional reviews. The impact of these two previous reviews on the field of Indigenous languages and health by evidence of their citations despite not being officially published indicate both a need and a potential impact for the current scoping review.

The objective of this scoping review is to assess and synthesize available literature on the relationships between Indigenous linguistic vitality/revitalization and health in Indigenous populations in anglophone, settler-colonial societies.

### Review question

What is the extent and scope of literature on the relationships between health and Indigenous language vitality/revitalization in Indigenous populations in anglophone settler-colonial societies?

### Keywords

Indigenous languages; health; Indigenous peoples; linguistic vitality; language revitalization; scoping review.

### Eligibility criteria

##### Participants

##### This review will consider literature about communities that are Indigenous to Canada, the United States of America, Australia, and New Zealand as the populations of interest. Indigenous peoples in these contexts are defined as the original inhabitants at the time of contact by European settlers.

##### Concept

Literature will be included that has a major focus on describing connections or relationships between some aspect(s) of health or wellbeing in an Indigenous population and an Indigenous language(s).

Indigenous communities’ conceptions of health often encompass more than just the absence of disease and include spiritual elements, as previously discussed by Boddington & Räisänen from an Aboriginal Australian perspective (Boddington & Räisänen, 2009). The authors of this protocol similarly acknowledge the possible restrictive and colonial connotations of the English term “health”, and define health for this review as including the full spectrum of prevention, determinants of health, health promotion, healing, spirituality, and other meanings of wellness and wellbeing. The definitional and conceptual challenges surrounding health will be further addressed in the scoping review publication in the context of the search results.

Indigenous languages will be considered in the context of their vitality, including the number and proportion of speakers, level of documentation, and intergenerational transmission, as well as revitalization and reclamation initiatives (Pine & Turin, 2017). Literature primarily about culture will only be included if it explicitly and majorly focuses on Indigenous languages as part of culture.

##### Context

This review will consider literature about Indigenous populations in Canada, the United States of America, Australia, and New Zealand. These four countries are all settler colonial contexts where English is the dominant language, which are two main factors that have resulted in similar contexts for Indigenous languages.

##### Types of Sources

Eligible sources of evidence will be in the written form and include academic articles, book chapters indexed in academic databases, theses (undergraduate and graduate) and dissertations, conference proceedings, course syllabi, government documents, and reports and fact sheets published by stakeholders including universities, associations, non-governmental organizations (NGOs), intergovernmental organizations (IGOs) and research agencies.

Ineligible sources of evidence include informal communications such as blogs, emails, and social media posts, non-scholarly books, and books published by vanity or predatory publishers.

### Methods

The proposed scoping review will be conducted in accordance with the JBI methodology for scoping reviews (Peters et al., 2020). The grey literature search strategy will be guided by a template produced by Jackie Stapleton, based on the methods used in a systematic review by Godin et al (2015). University reference librarians with subject specializations in health (U.E.) and Indigenous studies (K.D-L.) will collaborate on the search strategy.

##### Search strategy

The search strategy will aim to locate both published and unpublished (grey or difficult to locate) literature. Initial limited searches of MEDLINE (Ovid), PsycInfo, and Google Scholar were undertaken to identify articles on the topic. The text words contained in the titles and abstracts of relevant articles, and the index terms used to describe the articles were used to develop a full search strategy for MEDLINE (see Appendix 1). The search strategy, including all identified keywords and index terms, will be adapted for each included database and information source.

The databases to be searched include MEDLINE (Ovid), PsycInfo (EBSCO), Cumulative Index to Nursing & Allied Health Literature (CINAHL Complete: EBSCO), Bibliography of Native North Americans (EBSCO), Australian Education Index (ProQuest), Educational Resource Information Centre (ERIC: EBSCO), MLA International Bibliography (EBSCO), Web of Science Core Collection, Communication & Mass Media Complete (EBSCO), Linguistics and Language Behavior Abstracts (LLBA: ProQuest). Sources of unpublished studies/grey literature to be searched include desLibris, PsycEXTRA (EBSCO), Native Health Database, iPortal, ProQuest Dissertations & Theses Global, Theses Canada, OALster, Analysis & Policy Observatory, Informit Indigenous Collection, and Advanced Google.

The reference list of all included sources of evidence will be screened for additional sources. The reviewers may contact authors of literature for further information, where relevant. The entire search strategy and results will be recorded and presented in a transparent and auditable format.

Only literature published in English will be included due to feasibility limitations including the language knowledge of the study team and a limited timeline. There will be no date limits applied to inclusion.

##### Study/Source of Evidence selection

A pilot test was conducted in November 2020 by the two reviewers (L.H. and J.S.) with the MEDLINE search strategy and inclusion criteria, and 100% agreement was reached through discussion on 35 titles and abstracts. The test was repeated in January 2021 to incorporate minor amendments to the strategy and inclusion criteria; agreement was reached through discussion on 36 titles and abstracts.

Following the search, all identified citations will be collated and uploaded into Zotero, deduplicated, and uploaded to Covidence. Titles and abstracts of articles from academic databases will be screened by the two independent reviewers for assessment against the inclusion criteria. Literature from the grey literature search will be screened for the first stage of inclusion by one reviewer. The literature included from these two steps will be brought forward for full text review by the two independent reviewers for full text inclusion *(revised March 19, 2021 due to resource constraints: The literature included from these two steps will be brought forward for full text review by one reviewer (L.H.), who will consult with the other reviewers to resolve any ambiguities*). Reasons for exclusion of sources of evidence at full text will be recorded and reported in the scoping review. Any disagreements that arise between the reviewers in the selection process will be resolved through consensus discussion or by the decision of a third reviewer. The results of the search and the inclusion process will be reported in full in the final scoping review and presented in a Preferred Reporting Items for Systematic Reviews and Meta-Analyses extension for Scoping Review (PRISMA-ScR) flow diagram (Tricco et al., 2018).

##### Data Extraction

Data will be extracted from literature included in the scoping review by one reviewer using a data extraction tool in Covidence developed and piloted by two members of the review team. The reviewer will select at least 10% of the data extraction to be validated by a second reviewer, choosing the most complex and challenging sources. Disagreements will be resolved through discussion or involvement of a third reviewer.

The data extracted will include specific details about the participants, concept, context, study methods and key findings relevant to the review question/s. Potentially relevant additional references will also be extracted, and subjected to the full text screening process. A draft extraction form is provided in Appendix II. The extraction tool will be modified and revised as necessary during the process of extracting data from each included evidence source, and modifications will be detailed in the scoping review publication. If appropriate and where required, authors of literature will be contacted to request missing or additional data.

##### Data Analysis and Presentation

The data will be presented in: 1) a summary table of participant, source and relevant findings for each paper (Table 1); 2) tables, histograms and boxplots displaying the frequencies and counts recorded from each column in the data extraction tool (Figures 1 and 2); and 3) a narrative summary relating the results to the review objective and questions. Any additional analyses that are added iteratively will be identified as such in the scoping review manuscript.

### Acknowledgements

The authors thank Sarah Dupont for research support.

### Funding

The authors have no specific funding to report.

### Conflicts of interest

There is no conflict of interest in this project.

Tricco, A.C., Lillie, E., Zarin, W. et al. PRISMA Extension for Scoping Reviews

(PRISMA-ScR): Checklist and Explanation. *Ann Intern Med*. 2018;169:467-473.

United Nations. (2016, Apr 6). UNESCO call for int. year of Indigenous languages 2019. Retrieved 21 Jan 2021 from:<https://www.un.org/development/desa/indigenouspeoples/news/2018/04/establishment-of-the-steering-committee-for-the-organization-of-the-2019-international-year-of-indigenous-languages/>

United Nations General Assembly. (2007, Oct 2). United Nations Declaration on the Rights of Indigenous Peoples : resolution / adopted by the General Assembly, A/RES/61/295. Retrieved 21 Jan 2021 from: https://www.refworld.org/docid/471355a82.html

UNESCO. (2020, Feb 28). Upcoming Decade of Indigenous Languages (2022 – 2032) to focus on Indigenous language users’ human rights. Retrieved 21 Jan 2021 from:<https://en.unesco.org/news/upcoming-decade-indigenous-languages-2022-2032-focus-indigenous-language-users-human-rights>

van Beek, S. (2016). Intersections: Indigenous Language, Health and Wellness : First Peoples’ Cultural Council. Review of Literature; Deliverable for FNIS UBC Practicum 2016. First Peoples’ Cultural Council; University of British Columbia.

Whalen, D.H., Moss, M., & Baldwin, D. (2016) Whalen DH, Moss M and Baldwin D. Healing through Language: Positive Physical Health Effects of Indigenous Language Use [Version 1; Peer Review: 2 Approved with Reservations]. *F1000Research* *5*(852),

### Tables

##### Table 1: Sample summary table of reviewed literature

Table 1: Summary of reviewed literature

| Citation | Type of Literature | Population characteristics | Relationships between Indigenous languages and health reported |
| --- | --- | --- | --- |

### Figures

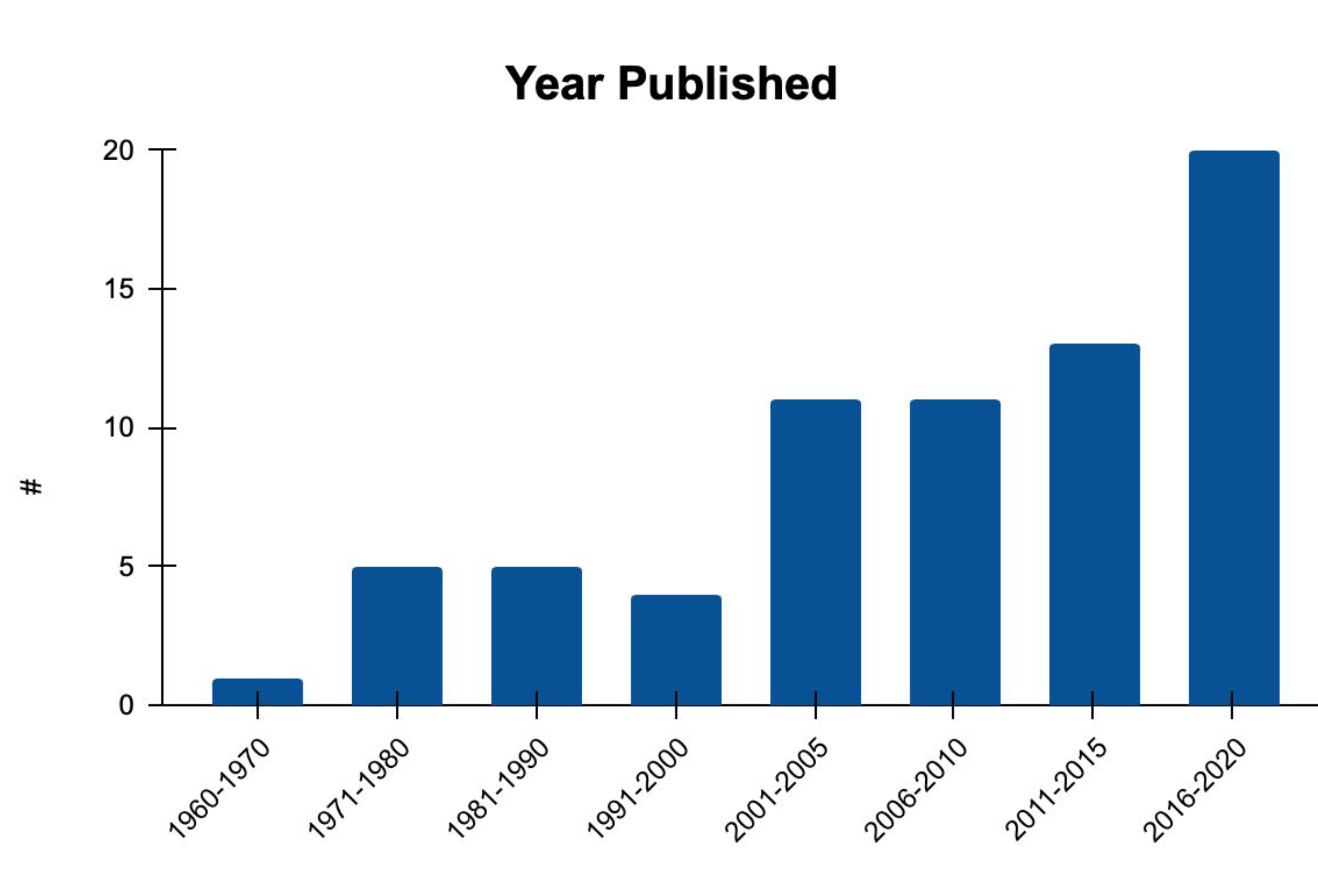

Figure 1: Sample histogram of years of publication of reviewed literature.

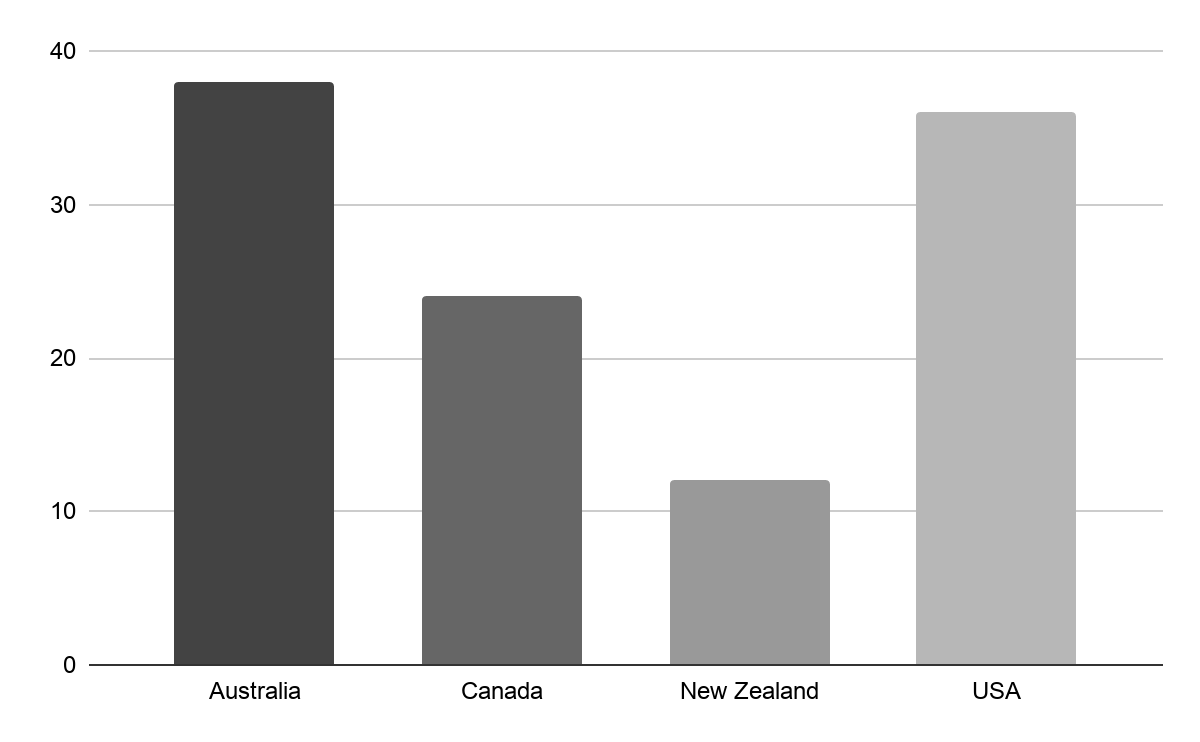

Figure 2: Sample histogram of countries of Indigenous communities in reviewed literature.

### Appendices

##### Appendix I: Sample search strategy

**Search strategy for MEDLINE (Ovid)**

| Subject Headings | ((exp American Native Continental Ancestry Group/) or (Oceanic Ancestry Group/) or (exp Indigenous Peoples/)) and exp language/ |
| --- | --- |
| All fields keywords | (aborig* or indigen* or first nation or first nations or first peoples or native people* or (alaska* adj3 native*) or inuit* or inu?k or Eskimo* or metis or (native* adj3 america*) or (hawai* adj2 native*) or pacific islander* or (pacific island* adj3 native*) or america* indian* or amerindian* or Torres Strait Island* or Maori*) and (language* or linguist* or revitali* or recla?m*) |

Limits applied: English language

##### Appendix II: Draft data extraction criteria

| **Item name** | **Details** |
| --- | --- |
| Title | Title of source |
| Author(s) | Author(s) of source |
| Type of literature | e.g. Academic article, academic book chapter, conference proceeding |
| Source of evidence for link between Indigenous languages and health | Does the evidence come from their primary research (if yes, describe methods), citing other literature (if yes, insert citations), personal experiences or narratives, not specified, etc. |
| Country(ies) of Indigenous population(s) | The country(ies) that populations discussed in the source in relation to languages and health are Indigenous to. |
| Name of Indigenous community(ies) | The names of the Indigenous community(ies) discussed in the work in relation to language and health. |
| Sample size, if applicable | For primary research |
| Health concept | What aspect of health or wellness is linked to language ? |
| Language concept | What aspect(s) of linguistic vitality, reclamation or revitalization is discussed? |
| Key findings | Key findings relevant to review questions |
| Direction of association between Indigenous language vitality and health reported | Choices: Positive (Indigenous linguistic vitality has a positive effect on health); Negative (negative effect on health); Mixed (reports mixed effects); Other |
| Potentially relevant references to screen for inclusion | References from the works cited list to screen for relevance. |
| Other notes | Any additional notes on the piece that are relevant to the reviewer. |
