## Appendix C for "Linguistic vitality improves health and wellbeing in Indigenous communities: a scoping review"

Appendix C: Academic Database Search Strategy

The academic search was conducted on January 26, 2021.

| **Database** | **Search terms** | **Limits applied** | **Results** |
| --- | --- | --- | --- |
| PsycInfo (EBSCO) | (( DE "Indigenous Populations" OR DE "Alaska Natives" OR DE "American Indians" OR DE "Inuit" OR DE "Pacific Islanders" OR DE "Hawaii Natives" ) OR TX ( aborig* OR indigen* OR “first nation” OR “first nations” OR “first peoples” OR “native people*” OR alaska* N3 native* OR inuit* OR inu#k OR eskimo* OR metis OR native* N3 america* OR hawai* N2 native* OR “pacific islander*” OR pacific island* N2 native* OR “american indian*” OR amerindian* OR “torres strait island*” OR maori* )) AND (( DE "Language" OR DE "Dialect" OR DE "Foreign Languages" OR DE "Interpreters" OR DE "Monolingualism" OR DE "Multilingualism" OR DE "Bilingualism" OR DE "Native Language" OR DE "Sign Language" OR DE "Written Language" OR DE "Linguistics" OR DE "Cognitive Linguistics" OR DE "Ethnolinguistics" OR DE "Metalinguistics" OR DE "Neurolinguistics" OR DE "Orthography" OR DE "Psycholinguistics" OR DE "Sociolinguistics" OR DE "Verbal Communication" OR DE "Language Proficiency" OR DE "Manual Communication" OR DE "Oral Communication" ) OR TX ( language* OR linguist* OR revitali* OR recla#m*)) | Limited to language: English. | 3,903 |
| MEDLINE (Ovid) | 1 exp American Native Continental Ancestry Group/  2 exp Oceanic Ancestry Group/  3 exp Indigenous Peoples/  4 exp Language/  5 (aborig* or indigen* or first nation or first nations or first peoples or native people* or (alaska* adj3 native*) or inuit* or inu?k or Eskimo* or metis or (native* adj3 america*) or (hawai* adj2 native*) or pacific islander* or (pacific island* adj3 native*) or america* indian* or amerindian* or Torres Strait Island* or Maori*).mp. [mp=title, abstract, original title, name of substance word, subject heading word, floating sub-heading word, keyword heading word, organism supplementary concept word, protocol supplementary concept word, rare disease supplementary concept word, unique identifier, synonyms]  6 (language* or linguist* or revitali* or recla?m*).mp. [mp=title, abstract, original title, name of substance word, subject heading word, floating sub-heading word, keyword heading word, organism supplementary concept word, protocol supplementary concept word, rare disease supplementary concept word, unique identifier, synonyms]  7 1 or 2 or 3 or 5  8 4 or 6  9 7 and 8  10 limit 9 to english |  | 2,835 |
| CINAHL Complete (EBSCO) | (aborig* OR indigen* OR “first nations” OR “first nation” OR “first peoples” OR “native people*” OR alaska* N3 native* OR inuit* OR inu#k OR eskimo* OR metis OR native* N3 america* OR hawai* N2 native* OR “pacific islander*” OR pacific island* N2 native* OR “american indian*” OR amerindian* OR “torres strait island*” OR maori* OR (MH "Indigenous Peoples+")) AND (language* OR linguist* OR revitali* OR recla#m* OR (MH "Language") OR (MH "Accents and Dialects") OR (MH "Language Arts+") OR (MH "Linguistics+") OR (MH "Multilingualism") OR (MH "Written Language+")) | Limited to language: English. | 1,659 |
| ERIC (Educational Resource Information Centre) (EBSCO) | (aborig* OR indigen* OR “first nation” OR “first nations” OR “first peoples” OR “native people*” OR alaska* N3 native* OR inuit* OR inu#k OR eskimo* OR metis OR native* N3 america* OR hawai* N2 native* OR “pacific islander*” OR pacific island* N2 native* OR “american indian*” OR amerindian* OR “torres strait island*” OR maori*) AND (language* OR linguist* OR revitali* OR recla#m*) AND (heal* OR wellness OR wellbeing OR well-being) | Limited to language: English. | 600 |
| Bibliography of Native North Americans (EBSCO) | (heal* OR wellness OR wellbeing OR well-being) AND (language* OR linguist* OR revitali* OR recla#m*) | Limited to language: English; source type: all except for newspapers. | 308 |
| Australian Education Index (ProQuest) | (aborig* OR indigen* OR “first nation” OR “first nations” OR “first peoples” OR “native people*” OR alaska* N/3 native* OR inuit* OR inu*k OR eskimo* OR metis OR native* N/3 america* OR hawai* N/2 native* OR “pacific islander*” OR pacific island* N/2 native* OR “american indian*” OR amerindian* OR “torres strait island*” OR maori*) AND (language* OR linguist* OR revitali* OR recla*m*) AND (heal* OR wellness OR wellbeing OR well-being) | Search: anywhere. | 196 |
| Linguistics and Language Behavior Abstracts (LLBA) (ProQuest) | (aborig* OR indigen* OR “first nation” OR “first nations” OR “first peoples” OR “native people*” OR alaska* N/3 native* OR inuit* OR inu*k OR eskimo* OR metis OR native* N/3 america* OR hawai* N/2 native* OR “pacific islander*” OR pacific island* N/2 native* OR “american indian*” OR amerindian* OR “torres strait island*” OR maori*) AND (heal* OR wellness OR wellbeing OR well-being) | Limited to language: English. Search field: anywhere except full text. | 191 |
| Communication & Mass Media Complete (EBSCO) | (aborig* OR indigen* OR “first nation” OR “first nations” OR “first peoples” OR “native people*” OR alaska* N3 native* OR inuit* OR inu#k OR eskimo* OR metis OR native* N3 america* OR hawai* N2 native* OR “pacific islander*” OR pacific island* N2 native* OR “american indian*” OR amerindian* OR “torres strait island*” OR maori*) AND (language* OR linguist* OR revitali* OR recla#m*) AND (heal* OR wellness OR wellbeing OR well-being) | Limited to language: English. | 98 |
| MLA International Bibliography (EBSCO) | (aborig* OR indigen* OR “first nation” OR “first nations” OR “first peoples” OR “native people*” OR alaska* N3 native* OR inuit* OR inu#k OR eskimo* OR metis OR native* N3 america* OR hawai* N2 native* OR “pacific islander*” OR pacific island* N2 native* OR “american indian*” OR amerindian* OR “torres strait island*” OR maori*) AND (language* OR linguist* OR revitali* OR recla#m*) AND (heal* OR wellness OR wellbeing OR well-being) | Limited to language: English. | 54 |
| Web of Science Core Collection: Science Citation Index Expanded (1900-present), Social Sciences Citation Index (1956-present), Arts and Humanities Citation Index (1975-present), Conference Proceedings Citation Index-Science (1990-present), Conference Proceedings Citation Index-Social Sciences & Humanities (1990-present),  Emerging Sources Citation Index (2015-present). | # 1 TI=((aborig* OR indigen* OR “first nation” OR “first nations” OR “first peoples” OR “native people*” OR alaska* NEAR/3 native* OR inuit* OR inu*k OR eskimo* OR metis OR native* NEAR/3 america* OR hawai* NEAR/2 native* OR “pacific islander*” OR pacific island* NEAR/2 native* OR “american indian*” OR amerindian* OR “torres strait island*” OR maori*) AND (language* OR linguist* OR revitali* OR recla*m*) AND (heal* OR wellness OR wellbeing OR well-being))  # 2: AB=((aborig* OR indigen* OR “first nation” OR “first nations” OR “first peoples” OR “native people*” OR alaska* NEAR/3 native* OR inuit* OR inu*k OR eskimo* OR metis OR native* NEAR/3 america* OR hawai* NEAR/2 native* OR “pacific islander*” OR pacific island* NEAR/2 native* OR “american indian*” OR amerindian* OR “torres strait island*” OR maori*) AND (language* OR linguist* OR revitali* OR recla*m*) AND (heal* OR wellness OR wellbeing OR well-being) )  #3: #2 OR #1 |  | 20 |
