## Appendix D for "Linguistic vitality improves health and wellbeing in Indigenous communities: a scoping review"

Appendix D: Gray Literature Search Strategy

### Part 1: Identifying and searching relevant websites

##### A) Identify relevant authorities:

- We compiled a list of websites representing authorities, organizations or stakeholders who publish and/or store documents relevant to our research question through the following steps:
  - Searched Google using the term “Indigenous languages and health” with the region set as each Canada, USA, Australia and New Zealand. Reviewed the first 5 pages of results.
  - Searched Google using the term “Maori languages and health” with the region set as New Zealand. Reviewed the first 5 pages of results.
  - Identified relevant websites from the Canadian Agency for Drugs and Technologies in Health (CADTH) Grey Matters checklist.
  - Elicited suggestions from research team members.

##### B) Documenting Advanced Google search of each site from part A

- Used Advanced Google to search each site from part A. Reviewed the first 5 pages of results.

#### Documentation:

| **Date** | **Organization name & website URL** | **Search terms** | **# of items uploaded to citation management software** |
| --- | --- | --- | --- |
| Jan 29, 2021 | <https://www.nccih.ca/en/> | 1) Indigenous languages health 2) Indigenous languages wellness 3) languages health | 1) 0 2) 0 3) 0 |
| Jan 29, 2021 | <https://cichprofile.ca/> | 1) Indigenous languages health 2) Indigenous languages wellness 3) Aboriginal languages wellness 4) Aboriginal languages health | 1) 1 2) 0 3) 0 4) 0 |
| Jan 29, 2021 | <https://www.who.int/> | 1) Indigenous languages health | 1) 0 |
| Jan 29, 2021 | <https://sencanada.ca/en> | 1) Indigenous languages health | 1) 0 |
| Feb 1, 2021 | [https://www.statcan.gc.ca/eng/](https://www.statcan.gc.ca/eng/start) | 1) Indigenous languages health 2) First nations languages health 3) Michif health | 1) 0 2) 0 3) 0 |
| Feb 1, 2021 | <https://www.un.org/en/> | 1) Indigenous languages health | 1) 0 |
| Feb 1, 2021 | <https://www.afn.ca/> | 1) Indigenous languages health 2) languages health | 1) 2 2) 2 |
| Feb 1, 2021 | <https://fpcc.ca/> | 1) languages health | 1) 2 |
| Feb 1, 2021 | <https://www.cihi.ca/en> | 1) Indigenous languages health 2) languages and health | 1) 0 2) 0 |
| Feb 1, 2021 | <https://en.iyil2019.org/> | 1) Indigenous health | 1) 1 |
| Feb 1, 2021 | <https://www.firstlanguages.org.au/> | 1) health 2) wellness | 1) 2 2) 0 |
| Feb 1, 2021 | <https://www.indigenous.gov.au/> | 1) languages and health 2) language and wellness | 1) 2 2) 0 |
| Feb 1, 2021 | <https://www.reconciliation.org.au/> | 1) languages and health | 1) 1 |
| Feb 1, 2021 | <https://www.tpk.govt.nz/en> | 1) health and language 2) te reo and health | 1) 0 2) 0 |
| Feb 1, 2021 | <https://www.health.govt.nz/> | 1) Maori language and health 2) “te reo” and health | 1) 0 2) 0 |
| Feb 1, 2021 | <https://www.canada.ca/en/indigenous-services-canada> | 1) languages health 2) languages wellness | 1) 0 2) 1 |

### Part 2: Gray Literature Database Search

#### Search Plan

1. Search a variety of word combinations.
2. View results from most to least relevant.

#### Documentation:

| **Date** | **Database** | **Search terms** | **Limits applied, search strategy, and number reviewed** | **# of items uploaded to citation management software** |
| --- | --- | --- | --- | --- |
| Feb 2, 2021 | des Libris | 1) Indigenous AND health AND language 2) Aboriginal AND health AND language | Limited to: all documents, English language. Reviewed the first 50 results for each search. | 1) 8 2) 7 |
| Feb 2, 2021 | PsycEXTRA (EBSCO) | 1) (( DE "Indigenous Populations" OR DE "Alaska Natives" OR DE "American Indians" OR DE "Inuit" OR DE "Pacific Islanders" OR DE "Hawaii Natives" ) OR TX ( aborig* OR indigen* OR “first peoples” OR “first nation” OR "first nations" OR “native people*” OR alaska* N3 native* OR inuit* OR inu#k OR eskimo* OR metis OR native* N3 america* OR hawai* N2 native* OR “pacific islander*” OR pacific island* N2 native* OR “american indian*” OR amerindian* OR “torres strait island*” OR maori* )) AND (( DE "Language" OR DE "Dialect" OR DE "Foreign Languages" OR DE "Interpreters" OR DE "Monolingualism" OR DE "Multilingualism" OR DE "Bilingualism" OR DE "Native Language" OR DE "Sign Language" OR DE "Written Language" OR DE "Linguistics" OR DE "Cognitive Linguistics" OR DE "Ethnolinguistics" OR DE "Metalinguistics" OR DE "Neurolinguistics" OR DE "Orthography" OR DE "Psycholinguistics" OR DE "Sociolinguistics" OR DE "Verbal Communication" OR DE "Language Proficiency" OR DE "Manual Communication" OR DE "Oral Communication" ) OR TX ( language* OR linguist* OR revitali* OR recla#m*)) | Reviewed the first 100 results. | 1) 8 |
| Feb 2, 2021 | Downtown Eastside Research Access Portal (DTES RAP) | 1) language | Limited to: Indigenous Peoples. Reviewed the first 100 results. | 1) 0 |
| Feb 4, 2021 | Native Health Database | 1) language* OR linguist* OR revitali* OR reclaim* OR reclamation | Entered the terms as keywords using the advanced search option. | 1) 16 |
| Feb 2, 2021 | iPortal | 1) (heal* OR wellness OR wellbeing OR well-being) AND (language* OR linguist* OR revitali* OR reclaim* OR reclamation) | Reviewed the first 100 results. | 1) 1 |
| Feb 4, 2021 | ProQuest Dissertations & Theses Global | 1) (aborig* OR indigen* OR “first peoples” OR “first nation” OR “first nations” OR “native people*” OR alaska* N/3 native* OR inuit* OR inu*k OR eskimo* OR metis OR native* N/3 america* OR hawai* N/2 native* OR “pacific islander*” OR pacific island* N/2 native* OR “american indian*” OR amerindian* OR “torres strait island*” OR maori*) AND (language* OR linguist* OR revitali* OR recla*m*) AND (heal* OR wellness OR wellbeing OR well-being) | Limited to: keyword search anywhere except full text, English language. Reviewed the first 100 results. | 1) 10 |
| Feb 4, 2021 | Theses Canada | 1) Indigenous health language* | Limited to: English language. Reviewed the first 100 results. | 1) 0 |
| Feb 4, 2021 | Networked Digital Library of Theses and Dissertations (NDLTD) Global ETD Search | 1) Indigenous AND health AND language* 2) Aboriginal AND health AND language* | Limited to: English language. Reviewed the first 50 results for each search. | 1) 4 2) 0 |
| Feb 4, 2021 | OAIster | 1) Indigenous AND health AND language 2) Indigenous AND health AND languages 3) Aboriginal AND health AND language 4) Aboriginal AND health AND languages | Entered each term as a separate keyword using the advanced search option. Limited to: English language, theses and dissertations. Reviewed the first 50 results for each search. | 1) 3 2) 4 3) 4 4) 0 |
| Feb 2, 2021 | Center for Research Libraries (CRL) Foreign Dissertation Database | 1) (aboriginal OR indigenous OR first nations OR first peoples OR native people OR alaska natives OR inuit OR inuk OR inuuk OR eskimo OR metis OR native american OR hawaii native OR pacific islander OR pacific island native OR american indian OR amerindian OR torres strait islander OR maori) AND (language OR linguistic OR revitalize OR revitalization OR reclaim OR reclamation) AND (health OR heal OR wellness OR wellbeing OR well-being) 2) Indigenous AND language AND health | Limited to: dissertations, English language. Reviewed the first 50 results for each search. | 1) 0  2) 0 |
| Feb 4, 2021 | Trove (National Library of Australia) | 1) Indigenous languages health | Reviewed the first 50 results for each search. Limited to: thesis, English language. Reviewed the first 100 results. | 1) 3 |
| Feb 4, 2021 | Analysis & Policy Observatory | 1) Indigenous languages health | Reviewed the first 100 results. | 1) 22 |
| Feb 8, 2021 | Informit Indigenous Collection | 1) language* health 2) language* wellness | Used the advanced search option. Limited to: products - Indigenous collection. Reviewed the first 100 results for each due to a high number of duplicate records. | 1) 4 2) 2 |

### Strategy 3: Advanced Google and Google Scholar searching

#### Search Plan

1. Search a variety of word combinations in Advanced Google and Google Scholar.
2. View results from most to least relevant.
3. Limit to English results only.
4. Review the first 50 results for each search.

#### Documentation:

| **Date** | **Search engine** | **Search strategy(s) including how items were selected** | **# of items uploaded to citation management software** |
| --- | --- | --- | --- |
| Feb 4, 2021 | Advanced Google | Indigenous languages health  Limit by language: English, and region: Canada | 4 |
| Feb 4, 2021 | Advanced Google | Indigenous languages wellness  Limit by language: English, and region: Canada | 5 |
| Feb 4, 2021 | Advanced Google | Indigenous languages health  Limit by language: English, and region: USA | 6 |
| Feb 4, 2021 | Advanced Google | Indigenous languages wellness  Limit by language: English, and region: USA | 4 |
| Feb 4, 2021 | Advanced Google | Aboriginal languages health  Limit by language: English, and region: Australia | 4 |
| Feb 4, 2021 | Advanced Google | Aboriginal languages wellness  Limit by language: English, and region: Australia | 4 |
| Feb 4, 2021 | Advanced Google | “te reo” health  Limit by language: English, and region: New Zealand | 1 |
| Feb 4, 2021 | Advanced Google | “te reo” wellness  Limit by language: English, and region: New Zealand | 3 |
| Feb 4, 2021 | Google Scholar | Indigenous languages health | 7 |
| Feb 4, 2021 | Google Scholar | Indigenous languages wellness | 6 |
