## Appendix E for "Linguistic vitality improves health and wellbeing in Indigenous communities: a scoping review"

Appendix E: Data Extraction Chart

| **Data Item** | **Guiding Information** |
| --- | --- |
| Year(s) of publication | Year of publication of each report of the study. |
| Literature review | Indicate “Yes” if the publication is primarily a review of existing literature.  Indicate “No” for publications that primarily present original data.  Indicate “Mixed” if the publication combines a significant review of the literature review with original data collection. |
| Academic or gray literature | Academic: article published in an academic journal or chapter in a scholarly book.  Grey: thesis, dissertation, conference proceeding, course syllabus, government document, report or fact sheet published by stakeholders such as universities, associations, non-governmental organizations (NGOs), intergovernmental organizations (IGOs) and research agencies.  Mixed: multiple publications for a single study that include both academic and gray literature sources. |
| Source type(s) | Indicate the specific publication type, i.e. article, thesis, report, etc. |
| Geographic coverage | Countries of the Indigenous communities who are centrally discussed. |
| Impactful quotes | Quotes highlighting the links between Indigenous linguistic vitality/revitalization and health/wellness. |
| Summary | 1-3 sentences summarizing the study’s key findings relevant to the links between Indigenous linguistic vitality/revitalization and health/wellness. |
| Other notes | Anything else of relevance to the study objective to note. |
