## Supplementary Material Table S1 for "Linguistic vitality improves health and wellbeing in Indigenous communities: a scoping review"

Summaries of Included Studies on Indigenous Languages and Health

- We have not changed the language used in the studies or editorialized. so we would like to disclose that we don’t necessarily support the terminology used in the studies to describe Indigenous peoples or health conditions. Language usually reflects the scholarly climate at the time of the studies.

| **Citation(s)** | **Geographic coverage** | **Summary of results** |
| --- | --- | --- |
| Aboriginal and Torres Strait Islander, Social Justice Commissioner, & Australian Human Rights Commission. (2009). Social Justice Report [Report]. https://humanrights.gov.au/our-work/chapter-3-introduction-social-justice-report-2009 | Australia | Chapter 3 of this report reviews the existing evidence and notes that cultural knowledge is lost along with Aboriginal and Torres Strait Islander languages, which can impact health and well-being. Social, emotional, cognitive, and health advantages of having connections to language are noted, including benefits in cognitive development of bilingualism starting in infancy. Culture and identity are protective factors that assist in the development of resilience. |
| Alvarez, A. R. G. (2019). ‘We Were Queens.’ Historical Loss among Native Hawaiians: Exploring Historical Trauma-informed Suicide Prevention [Ph.D.]. University of Denver. | USA | In this secondary analysis of qualitative interview data, it is reported that the Hawaiian participants experience shame, frustration, and some guilt with regard to speaking the Hawaiian language. The author connects this to the high rate of suicide in the Hawaiian community through a critical suicidology framework, in addition to other historical losses that Hawaiian peoples have experienced due to European colonization. |
| Amery, H. (1999). They Don’t Give Us the Full Story: Attitudes to Hospitalisation Amongst Yolŋu People of North-East Arnhem Land - A Comparative Study. ARDS Aboriginal Cultural and Creative Services Northern Territory. https://www.ards.com.au/resources-2/p/they-dont-give-us-the-full-story-attitudes-to-hospitalisation-amongst-yolu-people-of-north-east-arnhem-land-a-comparative-study | Australia | In this study, Yolŋu individuals were interviewed about their experiences of hospitalization. Their primary concerns were communication difficulties, including a lack of satisfaction with explanations concerning diagnosis and treatment, and the absence of interpreters in the hospital. |
| Amery, R. (2017). Recognising the communication gap in Indigenous health care. *Medical Journal of Australia*, 207(1), 13–15. https://doi.org/10.5694/mja17.00042 | Australia | The author of this commentary draws on existing literature to argue that communication gaps in healthcare for Aboriginal peoples due to cultural and linguistic differences contribute to worse health outcomes. They suggest that to address these gaps, communication methods should be taught in mainstream medical education and that there should be more use of interpreters. |
| Angelo, D., O’Shannessy, C., Simpson, J., Kral, I., Smith, H., & Browne, E. (2019). Well-being & Indigenous Language Ecologies (WILE): A strengths-based approach. Australian National University: ARC Centre of Excellence for the Dynamics of Language. https://openresearch-repository.anu.edu.au/bitstream/1885/186414/2/NILR%20LitReview_FINAL_29July2020_c.pdf | Australia  Canada  New Zealand | Based on their literature review, the authors identify eight links between Indigenous languages and well-being (“indicators”): land-based, spiritual, cultural identity, emotional health, physical health, educational, economic, and restorative. They draw on Australian case studies which highlight nine links to well-being: identity, purpose, culture, connection, community and family, country, public recognition and reconciliation, and overall health. Mediators of the links include the local configuration of languages (language ecology), contexts of use, individuals’ proficiency, and type of language (e.g. traditional languages compared to new languages such as Kriol). |
| Armstrong, E., Coffin, J., Hersh, D., Katzenellenbogen, J. M., Thompson, S. C., Ciccone, N., Flicker, L., Woods, D., Hayward, C., Dowell, C., & McAllister, M. (2019). ‘You felt like a prisoner in your own self, trapped’: The experiences of Aboriginal people with acquired communication disorders. *Disability and Rehabilitation, 43*:13, 1903-1916, . https://doi.org/10.1080/09638288.2019.1686073 | Australia | In this study involving interviews with Aboriginal people living with communication disorders and their family members/carers, language was linked to health in two key ways: 1) one of the patients studied was happier, more responsive and more animated when people spoke an Aboriginal language around them. It is hypothesized by the authors that this is because language is central to cultural identity; 2) it is reported that it is difficult to diagnose an Aboriginal language speaker with an acquired communication disorder if they have limited proficiency in English when a translator is not present, as communication with the patient is impaired. |
| Artuso, S., Cargo, M., Brown, A., & Daniel, M. (2013). Factors influencing health care utilisation among Aboriginal cardiac patients in central Australia: A qualitative study. *BMC Health Services Research, 13*(83). https://doi.org/10.1186/1472-6963-13-83 | Australia | In this study involving interviews and focus groups with cardiac patients, community members, health care providers, and community researchers in Alice Springs, language is identified as a barrier to health care for Aboriginal cardiac patients which results in sub-optimal quality of care, placing them at risk for subsequent cardiovascular events and other negative health outcomes. The main topic addressed in the article is the lack of medical information in Aboriginal languages and the limited access to interpreters. |
| Auger, N., Fon Sing, M., Park, A. L., Lo, E., Trempe, N., & Luo, Z.-C. (2012). Preterm birth in the Inuit and First Nations populations of Quebec, Canada, 1981-2008. *International Journal of Circumpolar Health*, 71(1), 17520. https://doi.org/10.3402/IJCH.v71i0.17520 | Canada | Based on an analysis of retrospective data on maternal mother tongue, language spoken at home, and preterm births in Quebec, this study reports that there are: 1) higher risks of preterm births for French/English speakers than Inuit language speakers in Inuit census areas of Quebec, and 2) higher risks of preterm births for Inuit language speakers living in areas of Quebec that were not Inuit or First Nations specific census areas, relative to French/English Speakers. One explanation proposed is that the Inuit language speakers living in their traditional communities may have stronger ties to their traditional cultures and languages, which may support their wellbeing and protect against preterm births. |
| Bach, D. (2016). What matters to the heart: Hawaiian belief and value systems relating to disease and spirituality [Doctoral dissertation, Saybrook University]. https://www.proquest.com/dissertations-theses/what-matters-heart-hawaiian-belief-value-systems/docview/1707929019/se-2 | USA | In this focus group study, language is one of the four beliefs and values identified by Hawaiian people relating to disease and spirituality. This included language learning and language use, as well as the psychological impact of language loss. The author ties all of the beliefs and values to the importance of spiritual balance. |
| Bach, M. J. (2014). Community space for decolonization and resistance: Kodiak Alutiiq Language Club participant perspectives [Master’s thesis, University of Alaska Fairbanks]. In ProQuest Dissertations and Theses. http://hdl.handle.net/11122/4537 | USA | Drawing on interviews, discussions, field notes, and observations, this study reports that participants in a weekly Kodiak Alutiiq Language Club experience healing through Elders sharing their language, stories, histories, culture and traditions. Language learning provides members with an opportunity to understand and heal from the difficult history of language suppression, and allows them to build relationships and undergo personal growth. |
| Balestrery, J. E. (2014). A multi-sited ethnographic study in Alaska: Examining the culture-communication nexus salient to Alaska native elders and conventional health and social services [Doctoral dissertation, University of Michigan]. https://www.proquest.com/docview/1534550874?pq-origsite=summon&accountid=14656 | USA | This ethnographic study identifies that a lack of culturally appropriate health care service provision in local languages impedes the quality of care for Alaska Native patients. |
| Ball, J. (2006). Talking points: Exploring needs and concepts for Aboriginal early childhood language facilitation and supports [Concept discussion paper]. Aboriginal Head Start in Urban and Northern Communities, Public Health Agency of Canada. <https://canadacommons.ca/artifacts/1183603/talking-points/1736732/#details=1> | Canada | This report drawing on consultations with scholars, specialists, and program managers concludes that language has an important role to play in securing Aboriginal children’s cultural identity and their connections with family, community, and spiritual life, and that language also supports child development. Achieving these health benefits requires culturally-appropriate supports, including resources for teaching heritage languages. |
| Ball, J., & Lewis, M. (2014). First Nations elders’ and parents’ views on supporting their children’s language development. Canadian Journal of Speech-Language Pathology and Audiology, 38(2), 224–237. <https://cjslpa.ca/files/2014_CJSLPA_Vol_38/No_02/CJSLPA_Summer_2014_Vol_38_No_2_Paper_7_Ball_Lewis.pdf> | Canada | This interview study reports that early First Nations language learning is conducive to the development of self-esteem and supports children to survive and thrive. First Nations Elders, grandparents and parents of young children expressed a strong preference for children to learn both their mother tongue and English in their early years to help them consolidate their cultural identity, which would be a foundation for their self-esteem and success in life. |
| Ball, J., Moselle, K., & Moselle, S. (2013). Contributions of culture and language in Aboriginal Head Start in urban and northern communities to children’s health outcomes: A review of theory and research. Ottawa, ON: Division of Children, Seniors & Healthy Development, Health Promotion and Chronic Disease Prevention Branch, Public Health Agency of Canada https://ecdip.org/wp-content/uploads/2022/01/Language-Culture-Child-Health-Theory-Research-Ball-Moselle-Moselle.pdf | Canada | In this report, previous literature is reviewed that highlights the mental health impacts of language loss and how language use can improve self-esteem and prevent suicide. Analyses are also conducted of the 2002-2003 and 2007-2010 Regional Health Surveys of First Nations Communities and the 2010-2011 Annual Report on the State of Inuit Culture and Society, revealing additional direct and indirect supportive impacts of languages on overall wellness, childhood oral health, suicide, depression, and mental health. The authors argue for the importance of language and culture in early childhood interventions such as Aboriginal Head Start programs. |
| Barnes, R., Josefowitz, N., & Cole, E. (2006). Residential schools: Impact on aboriginal students’ academic and cognitive development. Canadian Journal of School Psychology, 21(1–2), 18–32. https://doi.org/10.1177/0829573506298751 | Canada | This literature review reports that the prohibition of Indigenous languages in residential schools was an important factor in the negative impact of these schools on children's general psychological and cognitive development. This was related to both the challenges of maintaining fluency in a first language in this context and the lack of opportunities to become proficient in English while attending residential school. |
| Barwin, L., Crighton, E., Shawande, M., & Veronis, L. (2014). Teachings around Self-care and Medicine Gathering: Rebuilding Capacity Begins with Youth. Pimatisiwin: A Journal of Aboriginal & Indigenous Community Health, 11(3), 323–344. | Canada | The authors consulted with First Nations adults and youth in Manitoulin Island, Ontario and reported that language is a determinant of wellbeing, primarily because language is part of the maintenance of culture, traditions and identity. |
| Basso, K. H. (1990). ‘Speaking with names’: Language and landscape among the Western Apache. In Western Apache language and culture: Essays in linguistic anthropology (pp. 138–182). U Arizona Press. | USA | In this ethnographic study, the author describes how place names in the Western Apache languages can be healing through the practice of speaking with names (calling to memory and imagination a place with its traditional Apache place name). This can make people feel better mentally, re-establish psychological balance, replenish equanimity, rejuvenate hope, and heal sickness. |
| BC Association of Aboriginal Friendship Centres. (2020). Urban Indigenous Wellness Report: A BC Friendship Centre perspective. BC Association of Aboriginal Friendship Centres. <https://bcaafc.com/wp-content/uploads/2020/11/BCAAFC-Urban-Indigenous-Wellness-Report.pdf> | Canada | This report draws on a review of research, policies, programs and consultations with urban Indigenous peoples in British Columbia to identify how a disconnection from Indigenous ways of life, including language, has contributed to the decline of the health and wellness for this population. The authors describe language as integral to positive health outcomes and cite literature showing that language can contribute to greater levels of psychological well-being as well as children’s health when childcare is anchored in language. |
| Beckstein, A. (2015). The relationship between decision-making style and self-construal and the subjective happiness of Native Americans [Doctoral dissertation, Arizona State University]. https://www.proquest.com/docview/1744738249?pq-origsite=summon&https://www.proquest.com/pqdtglobal/advanced?accountid=14656 | USA | In this study, a cohort of Native American people living in Arizona were surveyed with psychometric scales to assess for a link between Native language ability and subjective happiness. No relationship was identified. |
| Bell, L., & Marlow, P. E. (2009). Visibility, Healing and Resistance: Voices from the 2005 Dena’ina Language Institute. Journal of American Indian Education, 48(1), 1–18. | USA | This qualitative study with participants in a language revitalization program (Dena’ina Language Institute) reports that they viewed language learning as a mechanism for healing. |
| Belton, S., Kruske, S., Jackson Pulver, L., Sherwood, J., Tune, K., Carapetis, J., Vaughan, G., Peek, M., McLintock, C., & Sullivan, E. (2018). Rheumatic heart disease in pregnancy: How can health services adapt to the needs of Indigenous women? A qualitative study. The Australian & New Zealand Journal of Obstetrics & Gynaecology, 58(4), 425–431. https://doi.org/10.1111/ajo.12744 | Australia | The results of this study involving observation and interviews reports that there is a lack of language-appropriate health education for Aboriginal women with rheumatic heart disease in the Northern Territory, and that this limited the participants’ understanding of the severity of their illness and its implications for childbearing. Health directives in written and spoken English with assumed biomedical knowledge were confusing for the patients and of limited use when delivered without interpreters or culturally appropriate supports. |
| Bergman, R. L. (1974). Paraprofessionals in Indian mental health programs. Psychiatric Annals, 4(11), 76–84. | USA | The author of this commentary argues that language interpreters are essential paraprofessionals in health care for Indigenous peoples, with a focus on Navajo peoples. Without interpreters, major misinterpretations and issues can occur which can be even more serious than the issues caused by cultural differences. |
| Biddle, N., & Swee, H. (2012). The Relationship between Wellbeing and Indigenous Land, Language and Culture in Australia. Australian Geographer, 43(3), 215–232. https://doi.org/10.1080/00049182.2012.706201 | Australia | In this analysis of data from the 2008 National Aboriginal and Torres Strait Islander Social Survey (NATSISS), a positive association between being currently engaged in learning an Indigenous language and happiness is identified. There is also a positive association between speaking or understanding an Indigenous language and sadness; however, this association disappears once a range of other characteristics (in particular, self-reported health) are controlled for. |
| Bird, E. K.-R. (2011). Health, education, language, dialect, and culture in First Nations, Inuit, and Métis communities in Canada: An overview. Canadian Journal of Speech-Language Pathology and Audiology, 35(2), 110–124. | Canada | The author of this article argues for the importance of speech-language pathologists being familiar with the Indigenous languages of the communities and individuals with whom they work to ensure proper assessment and treatment. |
| Bird, S. M., Wiles, J. L., Okalik, L., Kilabuk, J., & Egeland, G. M. (2008). Living with diabetes on Baffin Island: Inuit storytellers share their experiences. Canadian Journal of Public Health = Revue Canadienne de Sante Publique, 99(1), 17–21. | USA | This multi-case study identifies that limited access to health information in Inuktitut, including labels and signs in grocery stores, has a negative impact on people living with and preventing Type 2 diabetes mellitus on Baffin Island. The authors suggest that focusing on language barriers may help to improve the accessibility of knowledge about diabetes and nutrition and enhance relationships between non-Inuit health service providers and Inuit people. |
| Bird, S. R., Held, S., McCormick, A., Hallett, J., Martin, C., & Trottier, C. (2016). The Impact of Historical and Current Loss on Chronic Illness: Perceptions of Crow (Apsaalooke) People. International Journal of Indigenous Health, 11(1), 198–210. https://doi.org/10.18357/ijih111201614993 | Canada | This study involving qualitative interviews with Apsáalooke (Crow) people living with chronic illness reports that frustration over loss of language and desire to know more of the language are having a negative impact on the health of this population. |
| Bougie, É., Wright, S. C., & Taylor, D. M. (2003). Early Heritage-language Education and the Abrupt Shift to a Dominant-language Classroom: Impact on the Personal and Collective Esteem of Inuit Children in Arctic Québec. International Journal of Bilingual Education and Bilingualism, 6(5), 349–373. https://doi.org/10.1080/13670050308667791 | Canada | This quantitative study in Nunavik, a small, extremely isolated Arctic community, compares the self-esteem of four cohorts of Inuit children in English or French Grade 3 programs who had either completed an Inuttitut K-2 program or an English or French K-2 program. Based on measures at the beginning and the end of the Grade 3 school year, the self-esteem of the Inuit children who had transitioned from the Inuttitut program dropped significantly. There was no change in self-esteem of the Inuit children who had transitioned from the English or French K-2 program. The authors suggest that the abrupt transition to a new language (linguistic disadvantage), to not having any Inuit teachers, to not having a heritage language used in school (cultural discontinuity), and to a class including children who already know English or French (social comparison) negatively affects children's perceived competence and their self-esteem. |
| Bowker, D. M. (2017). Knowledge and beliefs regarding HPV and cervical cancer among Lakota women living on the Pine Ridge Reservation and cultural practices most predictive of cervical cancer preventive measures. [Doctoral dissertation, New Mexico State University]. https://www.proquest.com/docview/1954748224?pq-origsite=summon&accountid=14656 | USA | In this survey study, Lakota women who reported that the Lakota language is spoken in their home were 2.5 times more likely to report that they are recommended Pap smears for cervical cancer screening than those who do not speak Lakota in their home. No differences were identified between the two groups on three other cervical cancer preventative measures, nor between any of the preventative measures and three other variables representing being culturally influenced by language. |
| Brass, E. R. (2013). Restoring balance: Determinants of health and depressive symptoms in Aboriginal people [Doctoral thesis, University of Regina]. https://www.proquest.com/docview/1069254953?pq-origsite=summon&accountid=14656 | Canada | In this exploratory analysis of quantitative data from the 2001 Aboriginal Peoples Survey, Métis respondents who could speak, use, or understand their Aboriginal language were less likely to report depressive symptoms than those who could not speak, use, or understand the language. Not being able to speak, use, or understand the language were significant predictors of depressive symptoms. For all respondents (Inuit, Métis, and North American Indian), people who reported not using their Aboriginal language at home were more likely to consume four or more alcoholic drinks per day. |
| Brega, A. G., Henderson, W. G., Harper, M. M., Thomas, J. F., Manson, S. M., Batliner, T. S., Braun, P. A., Quissell, D. O., Wilson, A., Tiwari, T., & Albino, J. (2019). Association of ethnic identity with oral health knowledge, attitudes, behavior, and outcomes on the Navajo Nation. Journal of Health Care for the Poor and Underserved, 30(1), 143–160. https://doi.org/10.1353/hpu.2019.0013 | USA | In this cluster randomized trial involving quantitative surveys and oral health exams with a Navajo population, parents who were better able to speak their heritage language had greater confidence in their ability to manage their children's oral health, engaged in better oral health behaviors, and reported better oral health status. Language was not associated with oral health knowledge or attitudes, or any measures of pediatric oral health. |
| Brougham, D., & Haar, J. M. (2013). Collectivism, cultural identity and employee mental health: A study of New Zealand Māori. Social Indicators Research, 114(3), 1143–1160. https://doi.org/10.1007/s11205-012-0194-6 | New Zealand | In this quantitative survey-based study, a link is reported between cultural speaking ability (the ability to describe one's ancestral links in te reo Māori), strong alignment with collectivist workplace values, and lower anxiety and depression among Māori adults. |
| Brown, D. L. (2016). Daughters of the drum: Decolonizing health and wellness with Native American women. AlterNative: An International Journal of Indigenous Peoples, 12(2). https://doi.org/10.20507/AlterNative.2016.12.2.1 | USA | In this qualitative study involving photovoice and talking circles, Native American women in the Pacific Northwest described that language reclamation is part of resistance, survival, and the decolonization of health and wellness, and is a protective factor in buffering the impacts of historical trauma. |
| Brown, H. J. ., McPherson, G., Peterson, R., Newman, V., & Cranmer, B. (2012). Our land, our language: Connecting dispossession and health equity in an indigenous context. CJNR: Canadian Journal of Nursing Research, 44(2), 44–63. | Canada | In this article presenting a nursing perspective on previous research and original qualitative data from working with the ‘Namgis First Nation in British Columbia, preserving language and sustaining a connection to the land are identified as indicators of health and well-being. Language is described in data from both sources as shaping identity and fostering social inclusion, cohesion and connections. The dispossession of land and language are reported to threaten overall health and well-being and worsen existing illness conditions. |
| Bryce, S. (2002). Lessons from East Arnhem land. Improving adherence to chronic disease treatments. Australian Family Physician, 31(7), 617–621. | Australia | In this commentary based on the author's clinical experience over four and a half years in East Arnhem Land, language and world view are identified as considerable barriers to good communication between Yolŋu people and healthcare providers that must be overcome, and which contribute to low treatment adherence in chronic disease treatments. The author identifies two strategies that have proved somewhat successful: incentivizing providers to learn the traditional language, and teaching providers strategies such as using pictures and models with patients and learning key language concepts. |
| Cairney, S., Abbott, T., Quinn, S., Yamaguchi, J., Wilson, B., & Wakerman, J. (2017). Interplay wellbeing framework: A collaborative methodology ‘bringing together stories and numbers’ to quantify Aboriginal cultural values in remote Australia. International Journal for Equity in Health, 16(1), 68. https://doi.org/10.1186/s12939-017-0563-5 | Australia | This study aimed to validate a measure of wellbeing for Aboriginal peoples called The Interplay Wellbeing Framework and Survey. The analyses confirmed that language, among other factors, plays a key role in the interplay between health, education and employment as part of a holistic system of wellbeing for Aboriginal peoples in Australia. Literacy in an Aboriginal language was identified to have one of the strongest direct impacts on wellbeing of all the factors considered. |
| Cameron, N. (2017). He waipuna koropupū: Taranaki Māori wellbeing and suicide prevention. Journal of Indigenous Wellbeing Te Mauri Pimatisiwin, 2(2), Article 8. | New Zealand | In this community-engaged study including conversations and interviews with Māori peoples, it is reported that loss of language due to colonization has impacted wellness and contributed to higher suicide rates. It is concluded that suicide prevention must involve language revitalization and learning. |
| Carrasco, R. L., & Riegelhaupt, F. (2006). Language, culture, science and the sacred: Issues and concerns in curriculum development for indigenous Americans. In T. Omoniyi & J. A. Fishman (Eds.), Explorations in the sociology of language and religion. (pp. 260–277). John Benjamins Publishing Company. https://doi.org/10.1075/dapsac.20.20car | Canada  USA | In this analytical discussion paper drawing on previous research, it is reported that the eradication of Indigenous languages and cultures and the lingering negative effects of this loss have created a need for healing. Such healing is described as frequently involving the revitalization of languages and cultures. The authors describe how language is conducive to healing because it is sacred and is a factor in both physical and mental well-being, and that using language in ceremonies has powerful healing effects. |
| Cass, A., Lowell, A., Christie, M., Snelling, P. L., Flack, M., Marrnganyin, B., & Brown, I. (2002). Sharing the true stories: Improving communication between Aboriginal patients and healthcare workers. The Medical Journal of Australia, 176(10), 466–470. | Australia | In this study analyzing clinical interactions between Yolngu patients and staff as well as in-depth interviews, language barriers are identified as contributing to serious miscommunications which are often unrecognized by patients. These include fundamental issues in diagnosis, treatment and prevention. Miscommunications were reported to be compounded by the minimal use of interpreters despite their availability, and by frequently calling on family members to translate instead. Issues around cultural and linguistic differences were often linked. |
| Castellano, M. B. (2006). Final Report of the Aboriginal Healing foundation: Volume I: A Healing Journey: Reclaiming Wellness. Aboriginal Healing foundation. https://www.ahf.ca/downloads/final-report-vol-1.pdf  Kishk Anaquot Health Research (2006). Final Report of the Aboriginal Healing foundation: Volume II: Measuring Progress: Program Evaluation. https://www.ahf.ca/downloads/final-report-vol-2.pdf  Archibald, Linda (2006). Final Report of the Aboriginal Healing foundation: Volume III: Promising Healing Practices in Aboriginal Communities. https://www.ahf.ca/downloads/final-report-vol-3.pdf | Canada | This report summarizes the results of seven years of work by the Aboriginal Healing foundation that addressed the intergenerational legacy of Canada’s Indian residential school system. Language is interwoven throughout as both a cause of historic trauma and as a pillar for healing through speaking and revitalization. |
| Cheng, L.-R. L. (1989). Service Delivery to Asian/Pacific LEP Children: A Cross-Cultural Framework. Topics in Language Disorders, 9(3), 1–14. | USA | The author of this commentary argues that it is unethical for Speech-Language Practitioners to not provide linguistically and culturally competent care to Pacific Islander children (with focus on Chamorro people and people from the Northern Mariana Islands), such as when assessing clients for special needs and providing therapy. |
| Cheng, W. Y. C., Blum, P., & Spain, B. (2004). Barriers to effective perioperative communication in Indigenous Australians: An audit of progress since 1996. Anaesthesia and Intensive Care, 32(4), 542–547. | Australia | This study reports on an audit at a Northern Territory hospital which identified that communication difficulty between anesthetists and Indigenous Australians was 31 times higher than for non-Aboriginal patients, and that this was most commonly related to difficulty in speaking English. The anesthetic team utilized the Aboriginal interpreter service in only a minority of cases – over a two-month period, 29 Indigenous peoples with little or no proficiency in English underwent anesthesia without the support of an interpreter. These communication difficulties are a barrier for patients to properly understand the anesthetists' explanations. |
| Coe, K., Attakai, A., Papenfuss, M., Giuliano, A., Martin, L., & Nuvayestewa, L. (2004). Traditionalism and its relationship to disease risk and protective behaviors of women living on the Hopi reservation. Health Care for Women International, 25(5), 391–410. https://doi.org/10.1080/07399330490438314 | USA | For this survey study with Hopi women, the authors constructed a ‘traditionalism’ score with three dimensions: Hopi language usage, cultural participation, and percentage of time off-reservation. High levels of ‘traditionalism’ were significantly associated with disease protective behaviors, and inversely associated with risk factors. When language was analyzed in isolation (controlling for age, education, and marital status), it was significantly associated with practicing Hopi behaviors to keep healthy. |
| Cohen, B. (2001). The spider’s web: Creativity and survival in dynamic balance. Canadian Journal of Native Education, 25(2), 140–148. | Canada | The author of this commentary describes how maintaining Indigenous languages contributes to communities thriving. The author argues that language is the fundamental way to retain cultural values and instill pride and a sense of community, all of which are necessary for a community to heal itself. |
| Condon, J. R. (2004). Cancer in Indigenous Australians in the Northern Territory of Australia [Doctoral thesis, Charles Darwin University]. https://researchers.cdu.edu.au/en/studentTheses/cancer-in-indigenous-australians-in-the-northern-territory-of-aus  Condon, J. R., Cunningham, J., Barnes, T., Armstrong, B. K., & Selva-Nayagam, S. (2006). Cancer diagnosis and treatment in the Northern Territory: Assessing health service performance for Indigenous Australians. Internal Medicine Journal, 36(8), 498–505. | Australia | In this retrospective cohort study involving quantitative analyses of Indigenous patient data in the Northern Territory, the risk of cancer death was much higher for Indigenous language speakers than for English language speakers. Remote residence was only associated with higher risk of cancer death when Indigenous first language was not adjusted for. Indigenous first language speakers were less likely to be offered, or to choose and complete, curative treatment, to travel interstate when referred, or to use private medical services. There was no difference in completion of treatment after curative treatment had been recommended and chosen, nor for cancer type or localization of disease. |
| Cooke, M. J., Wilk, P., Paul, K. W., & Gonneville, S. L. H. (2013). Predictors of obesity among Métis children~ Socio-economic, behavioural and cultural factors. Canadian Journal of Public Health~Revue Canadienne de Sante Publique, 104(4), e298-303. | Canada | In this analysis of data from the 2006 Aboriginal Peoples Survey, Aboriginal language knowledge was associated with obesity in Métis boys aged 6-10 and Métis girls aged 11-14, but not in girls aged 6-10 or boys aged 11-14. The authors postulate that this relationship may be due to other factors not captured in the models such as family or community characteristics or geography. |
| Coombes, J., Hunter, K., Mackean, T., Holland, A. J. A., Sullivan, E., & Ivers, R. (2018). Factors that impact access to ongoing health care for First Nation children with a chronic condition. BMC Health Services Research, 18(1), 448. https://doi.org/10.1186/s12913-018-3263-y | Australia Canada New Zealand | This literature review identifies language as one of seven key barriers to access and ongoing care for First Nations children with chronic conditions. One of the studies included in the review reported that a lack of interpreters was a reason that some families did not attend their scheduled medical appointments. |
| Corbiere, A. O. (2000, May 1). Reconciling Epistemological Orientations: Toward a Wholistic Nishnaabe (Ojibwe/Odawa/Potawatomi) Education. Annual Meeting of the Canadian Indigenous and Native Studies Association (CINSA), Edmonton, Alberta. https://eric.ed.gov/?id=ED467707 | Canada | The author reviews existing literature to argue that providing mother tongue language instruction and creating an epistemological shift in First Nations schools can achieve the educational goals of increased Native student self-esteem, pride in one's heritage, and positive identity formation. |
| Counceller, A. G. L. (2011). Niugneliyukut (we are making new words): A community philosophy of language revitalization [Doctoral thesis, University of Alaska Fairbanks]. http://hdl.handle.net/11122/9049 | USA | This study looks at the Kodiak New Words Council, a contemporary heritage revitalization effort that entails development of new terms in Alutiiq, a severely endangered language. The author reports benefits to participants including intellectual benefits (learning, remembering, and mental stimulation) for both the learners and the Elders, and healing in many ways. |
| Crown, M., Duncan, K., Hurrell, M., Ootoova, R., Tremblay, R., & Yazdanmehr, S. (1993). Making HIV prevention work in the north. Canadian Journal of Public Health = Revue Canadienne de Sante Publique, 84(Supplement 1: AIDS and Health Promotion), S55-8. | Canada | Through examining two case studies, the authors argue that linguistically and culturally appropriate materials and concepts are crucial for education about HIV/AIDS in the Northwest Territories. Crucially, this involves community health representatives. |
| Crozier, S. (1999). An Investigation into the Correlation between Ethnolinguistic Vitality and Well-Being [Master's thesis, Concordia University]. https://www.collectionscanada.ca/obj/s4/f2/dsk1/tape7/PQDD_0002/MQ43541.pdf | Canada | In this study where Indigenous university students were administered questionnaires, all measurements of ethnolinguistic vitality were identified to be positively correlated with measurements of well-being, and many of these correlations were statistically significant. |
| Cummings, M. A. (1998). Acculturation, family variables, and cognition of a subgroup of American Indian children ages 3-9 [Doctoral dissertation, Utah State University]. https://www.proquest.com/docview/304381167?pq-origsite=gscholar&fromopenview=true | USA | The researchers administered two cognitive measures to preschool and young school-aged children of American Indian ancestry to examine the influence that 23 family variables and acculturation (including language) had on measures of spatial abilities. They failed to find evidence of any links between language and visual-spatial skills. |
| Cunningham, J. (2010). Socioeconomic status and self-reported asthma in Indigenous and non-Indigenous Australian adults aged 18-64 years: Analysis of national survey data. International Journal for Equity in Health, 9(18). https://doi.org/10.1186/1475-9276-9-18 | Australia | In this analysis of data on adults from the 2004/2005 National Aboriginal and Torres Strait Islander Health Survey (NATSIHS) and the National Health Survey (NHS), Indigenous people whose main language was not English had significantly lower odds of self-reporting asthma compared to Indigenous people whose main language spoken at home was English. |
| Cunningham, J., & Paradies, Y. C. (2012). Socio-demographic factors and psychological distress in Indigenous and non-Indigenous Australian adults aged 18-64 years: Analysis of national survey data. BMC Public Health, 12(95). https://doi.org/10.1186/1471-2458-12-95 | Australia | In this analysis of data on adults from the 2004/2005 National Aboriginal and Torres Strait Islander Health Survey (NATSIHS) and the National Health Survey (NHS), Aboriginal and Torres Strait Islander people who spoke a main language other than English at home had significantly lower odds of being classified as having 'very high psychological distress' when compared to those who spoke English at home. The finding was consistent among males and females and those living in remote and non-remote areas. |
| Cunningham, J., Sibthorpe, B., & Anderson, I. (1997). Occasional paper: Self-assessed health status, Indigenous Australians (Occasional Paper 4707.0). Australian Bureau of Statistics. https://www.abs.gov.au/ausstats/abs@.nsf/7884593a92027766ca%202568b5007b8617/ba123184a705c366ca2568ba00236570!OpenDocument | Australia | This study analyzed variables associated with self-assessed health status in the 1994 National Aboriginal and Torres Strait Islander Survey (NATSIS). Respondents who spoke English as their main language were significantly more likely to report poor or fair health than those whose main language was not English. This trend was consistent when they looked at how many long-term health conditions the respondents had and what health actions they had taken in the past two weeks, after adjustment for other factors. |
| D’Aprano, A., Silburn, S., Johnston, V., Robinson, G., Oberklaid, F., & Squires, J. (2016). Adaptation of the Ages and Stages Questionnaire for Remote Aboriginal Australia. Qualitative Health Research, 26(5), 613–625. https://doi.org/10.1177/1049732314562891 | Australia | In this research study, the Ages and Stages Questionnaire was translated into the native languages of a population of remote dwelling Australian Aboriginal children and adapted to be culturally appropriate. When it was tested with parents, Aboriginal Health Workers, and early childhood development experts, the adapted questionnaire was identified to have high face validity, to be culturally acceptable, relevant, and more accessible, and to promote literacy in Aboriginal languages. It is concluded that “optimal child development will likely not occur unless culturally appropriate early identification and intervention processes are in place” (p622). |
| Dana-Sacco, G. (2012). Health as a proxy for living the good life: A critical approach to the problem of translation and praxis in language endangered Indigenous communities. Fourth World Journal, 11(2), 7–24. https://doi.org/10.3316/informit.021244143249224 | Canada  USA | In this study involving interviews with Passamaquoddy-Maliseet language speakers, it was identified that speaking the language promotes healing and wellness, and that the language contains teachings about health. The author also identifies that translating information about health into English can remove important elements, which obscures the original meaning. |
| Davies, J., Bukulatjpi, S., Sharma, S., Davis, J., & Johnston, V. (2014). ‘Only your blood can tell the story’—A qualitative research study using semi-structured interviews to explore the hepatitis B related knowledge, perceptions and experiences of remote dwelling Indigenous Australians and their health care providers in northern Australia. BMC Public Health, 14, Article 1233. https://doi.org/10.1186/1471-2458-14-1233 | Australia | In this qualitative study using semi-structured interviews with Yolŋu people living with Hepatitis B, community members, and key informants in remote areas of the Northern Territory, culturally appropriate discussions in a patient’s first language using visual aids are identified as vital to improving communication and developing treatment partnerships. |
| Davis, B. (2012). Cultural Connectedness as Personal Wellness in First Nations Youth (403) [Master’s thesis, University of Western Ontario]. Electronic Thesis and Dissertation Repository. https://ir.lib.uwo.ca/etd/403 | Australia | Based on quantitative data from youth who completed the First Nations Regional Longitudinal Health Survey, this study reports no significant relationship between believing that First Nations language is a community strength and depression. While the authors conclude that language use did not appear to be a protective factor for depression, they assert that the study does not present sufficient evidence to conclude that language has a negative impact on wellness. |
| Davis, B., Mcgrath, N., Knight, S., Davis, S., Norval, M., Freelander, G., & Hudson, L. (2004). Aminina Nud Mulumuluna ('You Gotta Look After Yourself’): Evaluation of the use of traditional art in health promotion for Aboriginal people in the Kimberley region of Western Australia. Australian Psychologist, 39(2), 107–113. https://doi.org/10.1080/00050060410001701816 | Canada | This study evaluated the use of traditional language and art in health promotion materials for Aboriginal people in the Kimberley region and reports that these are viable methods for addressing the holistic needs of Aboriginal health promotion. |
| Department of Infrastructure, Transport, Regional, Australian Institute of Aboriginal and Torres Strait Islander Studies (AIATSIS), & Australian National University. (2020). National Indigenous Languages Report. Office of the Arts, Department of Infrastructure, Transport, Regional Development and Communications. https://www.arts.gov.au/documents/national-indigenous-languages-report-document | Australia | Chapter 2 of this report on Indigenous languages presents an analysis of the 2014-2015 National Aboriginal and Torres Strait Islander Social Survey (NATSIS) which reports links between speaking Aboriginal and Torres Strait Islander languages as the main language at home and individuals’ well-being and health. There was a particularly positive link with mental health, including ratings of life satisfaction and emotional well-being. |
| Devereux, G. (1942). The mental hygiene of the American Indian. Mental Hygiene, 26(1), 71–84. | USA | The author of this commentary reports on how language barriers between American Indian patients and psychiatrists in psychiatric facilities hinder communication. They describe the lack of equivalency of many concepts and that the linguistic isolation that hospitalized patients can face promotes deterioration of their conditions and is not conducive to recovery. |
| Dingwall, K. M., Gray, A. O., McCarthy, A. R., Delima, J. F., & Bowden, S. C. (2017). Exploring the reliability and acceptability of cognitive tests for Indigenous Australians: A pilot study. BMC Psychology, 5. | Australia | This study examined the reliability and acceptability of cognitive tests for a sample of Aboriginal people admitted to a hospital in the Northern Territory of Australia. The authors report that reliable cognitive assessment is challenged by the use of the English language on the test, and that there was general consensus amongst participants that instructions in their first language and use of interpreters and Aboriginal Liaison Officers would assist understanding and improve performance. They recommend using tests with a decreased reliance on language. |
| Dinku, Y., Markham, F., Angelo, D., Simpson, J., O’Shannessy, C., & Dreise, T. (2020). Language use is connected to indicators of wellbeing: Evidence from the National Aboriginal and Torres Strait Islander Social Survey 2014-15. (CAEPR Working Paper). Australian National University. Centre for Aboriginal Economic Policy Research, Canberra. https://caepr.cass.anu.edu.au/sites/default/files/docs/2020/8/CAEPR_WP_no_137_2020_Dinku_et_al.pdf | Australia | This analysis of quantitative data from the 2014–15 National Aboriginal and Torres Strait Islander Social Survey (NATSISS) reports that speaking an Indigenous language as a main language at home is significantly associated with social connectedness and positive emotional well-being, after controlling for English language proficiency and a range of individual and family-level characteristics. Indigenous language speakers were more likely than English-only speakers to report having difficulties accessing health services. The author suggests that Indigenous language use is less consistently correlated with health outcomes because they are most strongly determined by external structural forces (e.g. access to healthcare services) which are largely beyond Indigenous control. All effects reported differed across language ecology areas (English first language vs. Indigenous first language areas) and level of individual Indigenous language use (ranging from none to mainly using an Indigenous language). |
| Dockery, A. M. (2011). Traditional culture and the wellbeing of Indigenous Australians: An analysis of the 2008 NATSISS [CLMR Discussion paper]. Centre for Labour Market Research, Curtin University, Perth. | Australia | This analysis of data from the 2008 National Aboriginal and Torres Strait Islander Social Survey (NATSISS) reports that speaking Indigenous languages is associated with markedly superior self-reported health and a lower likelihood of risky alcohol use. The authors also report that use of Indigenous languages enhances happiness and mental health while simultaneously incurring psychological stress due to stronger feelings of discrimination. When analyzed by geography, the positive effects of fluency in Indigenous languages upon happiness and mental health are identified to accrue primarily in remote areas. |
| Duncan, G. E., McDougall, C. L., Dansie, E., Garroutte, E., Buchwald, D., & Henderson, J. A. (2014). Association of American Indian cultural identity with physical activity. Ethnicity & Disease, 24(1), 1–7. | USA | In this study involving the administration of surveys and laboratory tests to a large sample of Northern Plains American Indian people, participants who spoke both English and their Indigenous language at home as primary languages had significantly higher levels of total physical activity and sufficient leisure-time physical activity compared to those who spoke either language exclusively. |
| Durie, A. (1998). Emancipatory Maori Education: Speaking from the Heart. Language, Culture and Curriculum, 11(3), 297–308. | New Zealand | This paper explores how Māori have historically been faced with an assimilationist educational approach as a means of cultural invasion, which has had long term consequences on their well-being. Language revitalisation is seen as integral to strengthening cultural identity and enhancing well-being. |
| Durie, M. H. (1985). Maori health institutions. Community Mental Health in New Zealand, 2(1), 63–69. | New Zealand | In this commentary on Māori peoples' mental health, it is reported that Māori leaders regard the three principal institutions of good mental health as land (whenua), family (whanau) and language (reo). Specific aspects of language related to health include Māori concepts that cannot be translated to English and the “limitations of English as an adequate medium for expression,” the need for health professionals to communicate in te reo Māori, te reo Māori being an oral language and this putting students at a disadvantage, and that language is the center of culture so its absence is equated with incomplete personal development. |
| Durkin SR. (2008). Eye health programs within remote Aboriginal communities in Australia: A review of the literature. Australian Health Review, 32(4), 664–676. https://doi.org/10.1071/ah080664 | Australia | In a literature review of the most sustainable and culturally appropriate ways to implement eye health care programs in remote Aboriginal communities in Australia, culture and language barriers emerged as one of the ten areas to consider. |
| Eckhart, S. E. (1983). The relationship between language proficiency and cognitive ability for Navajo students enrolled in two types of instructional programs [Doctoral dissertation, Northern Arizona University]. | USA | This dissertation documents how a research team administered cognitive development tests to 9th and 10th grade Navajo students, and identifies that the students enrolled in a maintenance bilingual Navajo/English school had better cognitive abilities than those in a monolingual English school. This is interpreted as showing a relationship between language and cognitive development, and that the maintenance bilingual program serves to develop a healthier self-concept, a more positive attitude towards school, and pride in the native language, which subsequently promote language and cognitive development. |
| Edge, L., & McCallum, T. (2006). Métis identity: Sharing traditional knowledge and healing practices at Métis Elders’ gatherings. 4(2). https://journalindigenouswellbeing.co.nz/media/2019/01/5_Edge.pdf | Canada | This article reports on a collaborative community-based traditional knowledge project with Métis Elders. A return to the Michif language is identified as a factor contributing to Métis identity, health, healing and wellness. |
| Eliassen, B.-M., Braaten, T., Melhus, M., Hansen, K. L., & Broderstad, A. R. (2012). Acculturation and self-rated health among Arctic indigenous peoples: A population-based cross-sectional study. BMC Public Health, 12(948). https://doi.org/10.1186/1471-2458-12-948 | USA | In this questionnaire study involving quantitative analyses, no relationship is reported between spoken Indigenous language ability and self-rated health among Iñupiat people in Alaska. |
| Employment and Social Development Canada. (2018). Social isolation of seniors—Supplement to the social isolation and social innovation toolkit: A Focus on Indigenous Seniors in Canada [Toolkit]. Employment and Social Development Canada. https://www.canada.ca/en/employment-social-development/corporate/seniors/forum/social-isolation-indigenous.html | Canada | In this report integrating previous research, consultations including a workshop with Indigenous seniors, and an environmental scan of existing programs and services, language is identified as a protective factor against health crises and social isolation among Indigenous seniors, and as one of several important social determinants of health. A contributing factor to social isolation among Indigenous seniors is a past characterized by the suppression of their Indigenous languages and cultures and the enduring language differences that some experience. The report also describes how the social isolation of seniors from their families and communities can in turn contribute to the loss of a language and culture because it disrupts their important roles in preservation. |
| England Aytes, K. (2015). Memories hold hands: Perceptions of historical trauma and associated behavioral and emotional responses among four generations of American Indian (Cherokee) descendants [Doctoral dissertation]. Fielding Graduate University. | USA | Interviews with four generations of Cherokee individuals explored cultural losses as a source of trauma and stress. Loss of language was one of the most common losses mentioned by participants: nearly 46% of the respondents reported thinking of the loss of language for their families at least once a month, with 12.68% thinking of the loss daily. |
| Erasmus, M. T. (2019). Goyatıı̀ K’aàt’ıı̀ Ats’edee, K’aàt’ıı̀ Adets’edee: Ho! / Healing our languages, healing ourselves: Now is the time [Master’s thesis, University of Victoria]. http://hdl.handle.net/1828/10855 | Canada | The researcher studied six Indigenous adults reclaiming their ancestral languages in a Master's of Indigenous Language Revitalization program. They reported experiencing benefits related to health and overall well-being including physical fitness and healthy weight loss, emotional healing and healthy lifestyles, self-esteem and confidence, and a greater sense of identity. |
| Faleafa M. (2009). Community rehabilitation outcomes across cultures following traumatic brain injury. Pacific Health Dialog, 15(1), 28–34. | New Zealand | This study reports that neuropsychological tests for Traumatic Brain Injury lack language validity for Pacific Islanders. The result is that misleading conclusions can occur due to language differences, and therefore generate inappropriate guidance for rehabilitation. |
| Fiedeldey-Van Dijk, C., Rowan, M., Dell, C., Mushquash, C., Hopkins, C., Fornssler, B., Hall, L., Mykota, D., Farag, M., & Shea, B. (2017). Honoring Indigenous culture-as-intervention: Development and validity of the Native Wellness AssessmentTM. Journal of Ethnicity in Substance Abuse, 16(2), 181–218. https://doi.org/10.1080/15332640.2015.1119774 | Canada | In this study, people in treatment for addictions who predominantly spoke an Indigenous language consistently reported higher overall wellness, especially spiritually, on the Native Wellness Assessment developed by the researchers, compared to the other Indigenous people studied. This population group also practiced 7%–11% more cultural intervention practices than their counterparts. The authors conclude that this emphasizes the benefits of language-based initiatives as a contributor to culture-as-intervention. |
| Finau, S. A. (2000). Communicating health risks in the Pacific: Scientific construct and cultural reality. Asia-Pacific Journal of Public Health, 12(2), 90–97. https://doi.org/10.1177/101053950001200207 | Australia  New Zealand  USA | The authors use case studies and previous research to argue that language translation is essential for communicating health risks to multilingual and multicultural communities in the Pacific, including Indigenous communities. Challenges translating scientific concepts are described. |
| Findlay, L. C., & Kohen, D. E. (2013). Linking Culture and Language to Aboriginal Children’s Outcomes: Lessons from Canadian Data. FEL XVII: Endangered Languages beyond Boundaries: Community Connections, Collaborative Approaches and Cross-Disciplinary Research : Proceedings of the 17th FEL Conference, Carleton University, Ottawa, Ontario, Canada, 1-4 October 2013, 132–137. | Canada | This paper summarizes previous literature including national surveys that have identified links between Aboriginal child development and exposure to Aboriginal languages including cognitive, behavioral, and mental health benefits. |
| Fines, B. (1979). The native child in hospital. Dimensions in Health Service, 56(5), 62–64. | Canada | The author provides a commentary on the importance of language translation for Indigenous children in hospitals. |
| First Peoples’ Cultural Council. (2010). Report on the Status of BC First Nation’s Languages. https://fpcc.ca/wp-content/uploads/2020/07/2010-report-on-the-status-of-bc-first-nations-languages.pdf | Canada | This report identifies that health is directly impacted when First Nations languages are lost. It recommends promoting awareness of the direct correlation between language, culture, and wellness and to seek recognition of this link. |
| First Peoples’ Cultural Council. (2018). First Nations Languages and Health: Fact Sheet 10. https://fpcc.ca/wp-content/uploads/2020/08/Fact_Sheet_10_Language___Health.pdf | Canada | This fact sheet summarizing previous literature identifies First Nations languages as playing a role in lower youth suicide, lower diabetes, and resiliency. It indicates that language supports academic success which contributes to better health through increased income, and that Indigenous languages contain knowledge about health, wellbeing, and cultural values. They call for the development of community language programs for all ages. |
| FitzGerald, C. A., Fullerton, L., Green, D., Hall, M., & Peñaloza, L. J. (2017). The association between positive relationships with adults and suicide-attempt resilience in American Indian youth in New Mexico. American Indian and Alaska Native Mental Health Research, 24(2), 40–53. https://doi.org/10.5820/aian.2402.2017.40 | Canada  USA | Based on a survey, frequency of speaking a language other than English at home was not associated with the prevalence of suicide attempts among American Indian and Alaska Native youth in grades 9-12. The highest suicide attempt prevalence was in youth who spoke a language other than English at home all the time, but this difference was not statistically significant, and there were no differences in prevalence between the groups who spoke a language other than English at home “never”, “less than half the time“, “about half the time”, and “more than half the time but not all of the time”. |
| Fitzgerald, C. M. (2017). Understanding Language Vitality and Reclamation as Resilience: A Framework for Language Endangerment and ‘Loss’ (Commentary on Mufwene). Language, 93(4), e280–e297. <https://doi.org/10.1353/lan.2017.0072> | USA | The author of this commentary argues for a more holistic understanding of language and language revitalization to enable a better understanding of language revitalization's role as a protective factor. They draw on existing research showing links between language loss and worse physical and mental health, as well as improved health with revitalization and language learning. |
| Fuller-Thomson, E., Sellors, A. E., Cameron, R. E., Baiden, P., & Agbeyaka, S. (2019). Factors associated with recovery in Aboriginal people in Canada who had previously been suicidal. Archives of Suicide Research. https://doi.org/10.1080/13811118.2019.1612801 | Canada | In this analysis of data from the 2012 Aboriginal Peoples Survey, speaking an Aboriginal language is one of several factors associated with recovery/remission from suicidal ideation among Aboriginal peoples living off-reserve. |
| Galloway, T., Johnson-Down, L., & Egeland, G. M. (2015). Socioeconomic and cultural correlates of diet quality in the Canadian Arctic: Results from the 2007-2008 Inuit Health Survey. Canadian Journal of Dietetic Practice and Research, 76(3), 117–125. https://doi.org/10.3148/cjdpr-2015-006 | Canada | In this analysis of data from the 2007-2008 Inuit Health Survey involving a 24-hour dietary recall, associations are identified between an Inuit primary language in the home and several indicators of dietary quality set by the U.S. Institute of Medicine. Language is linked to the positive dietary quality indicators of traditional food consumption and lower sodium intake, but also to the negative dietary quality indicator of lower fiber intake. There is no link with cholesterol intake. In particular, women who spoke their language at home consumed a higher percentage of their total calories from protein and had much higher traditional food consumption than women who did not. |
| Garrett, M. T., Parrish, M., Williams, C., Portman, T. A. A., & Grayshield, L. (2014). Where power moves: Understanding and fostering resilience among Native American youth. In M. T. Garrett (Ed.), Youth and adversity: Psychology and influences of child and adolescent resilience and coping. (pp. 67–100). Nova Biomedical Books. | USA | This chapter draws on literature that has identified links between Indigenous languages and youth health, highlighting that language promotes positive youth development and community resilience, is a source of strength and resilience, supports sobriety programs, and connects people to spirituality. |
| Garvey, G., Cunningham, J., He, V. Y., Janda, M., Baade, P., Sabesan, S., Martin, J. H., Fay, M., Adams, J., Kondalsamy-Chennakesavan, S., & Valery, P. C. (2016). Health-related quality of life among Indigenous Australians diagnosed with cancer. Quality of Life Research, 25(8), 1999–2008. https://doi.org/10.1007/s11136-016-1233-6 | Australia | In a questionnaire assessing factors associated with health-related quality of life (HRQoL) for Indigenous Australian adult cancer patients 6 months post-diagnosis, excellent HRQoL was more likely among participants whose main language spoken at home was not English. |
| Gerlach, A. (2018). Exploring socially-responsive approaches to children's rehabilitation with Indigenous communities, families and children. National Collaborating Centre for Aboriginal Health. https://www.deslibris.ca/ID/10096166#details=1 | Canada | Bringing together previous literature and interviews, this report identifies that Indigenous languages are important for child development. The inclusion of Indigenous languages is identified as foundational to transforming the design and delivery of children’s rehabilitation programs and services so that they are respectful, meaningful, and experienced as culturally safe by Indigenous communities and families. The use of Western speech and language assessment tools can result in language differences being misdiagnosed as language disorders among Indigenous children, and the under- or over-identification of communication disorders. |
| Giuliano A, Papenfuss M, de Zapien JD, Tilousi S, & Nuvayestewa L. (1998). Breast cancer screening among Southwest American Indian women living on-reservation. Preventive Medicine, 27(1), 135–143. | USA | This interview study with women over 40 living on the Arizona Hopi reservation reports that use of the Hopi language some of the time was associated with a nearly twofold increase in the likelihood of having had a mammogram in the past 2 years when compared to no use of the language at any time. |
| Godard, L., & Czaykowska-Higgins, E. (2017). Language as a Link to Wellness. 5th International Conference on Language Documentation & Conservation. http://hdl.handle.net/10125/42017 | Canada | The authors cite previous literature linking Indigenous languages with overall health (specifically reduced suicide rates) and spirituality. They also draw on consultations that highlight how language supports social relationships and connection to community, ancestors, and identity. Language revitalization is identified as a contributor to strengthening these links to wellness and to healing. |
| Gonzales, A. A., Garroutte, E., Ton, T. G. N., Goldberg, J., & Buchwald, D. (2012). Effect of tribal language use on colorectal cancer screening among American Indians. Journal of Immigrant and Minority Health, 14(6), 975–982. https://doi.org/10.1007/s10903-012-9598-2 | USA | In this study involving the administration of a survey to a sample of adults residing on the Hopi reservation, no difference is reported in colorectal cancer screening rates or knowledge based on primary language or language spoken at home. |
| Goyal, S., Temple, V., Sawanas, C., & Brown, D. (2018). Cognitive profile of adults with intellectual disabilities from indigenous communities in ontario, canada. Journal of Intellectual and Developmental Disability. https://doi.org/10.3109/13668250.2018.1470160 | Canada | This study administered a cognitive assessment to adults with intellectual disabilities from Indigenous communities in Ontario and reports no significant relationship between first language and cognitive profile. |
| Grayshield, L., Rutherford, J. J., Salazar, S. B., Mihecoby, A. L., & Luna, L. L. (2015). Understanding and healing historical trauma: The perspectives of Native American elders. Journal of Mental Health Counseling, 37(4), 295–307. https://doi.org/10.17744/mehc.37.4.02 | USA | This qualitative research study reports that Elders identified language loss as a component of historical trauma and identified language learning as a way to heal from historical trauma. |
| Groot, G., Waldron, T., Barreno, L., Cochran, D., & Carr, T. (2020). Trust and world view in shared decision making with indigenous patients: A realist synthesis. Journal of Evaluation in Clinical Practice, 26(2), 503–514. https://doi.org/10.1111/jep.13307 | Australia  Canada  New Zealand  USA | The results of this literature review highlight the importance of incorporating Indigenous languages into shared decision making between health care providers and patients to facilitate effective communication. Difficulty translating medical terms and spiritual concepts between Indigenous languages and English is identified as a barrier to shared decision making. |
| Guenzel, N., & Struwe, L. (2020). Historical Trauma, Ethnic Experience, and Mental Health in a Sample of Urban American Indians[Formula: See text]. Journal of the American Psychiatric Nurses Association, 26(2), 145–156. https://doi.org/10.1177/1078390319888266 | USA | This mixed methods study reports that language loss is a major part of historical trauma that contributes to higher rates of mental health problems. |
| Guevremont, A., Arim, R., & Kohen, D. (2016). The Relationships between School Experiences and Mental Health Outcomes among Off-Reserve First Nations Youth. Aboriginal Policy Studies, 5(2). https://doi.org/10.5663/aps.v5i2.24703 | Canada | This secondary analysis of the 2012 Aboriginal Peoples Survey examines the relationships between school experiences and mental health outcomes for First Nations youth living off reserve. Youth who reported that their school supported First Nations language and culture were less likely to have considered suicide. However, this association was not significant after controlling for socio-demographic and other school-related factors. No links were reported between having ever been taught an Aboriginal language in classes at school and any measure of mental health. |
| Gullberg, S. R. W. (2008). Speaking of Health... Official Languages as Part of Quality Health Care in the Northwest Territories (p. 40) [Special Report]. Languages Commissioner of the Northwest Territories. https://olc-nt.ca/wp-content/uploads/2017/01/Speaking-of-Health.pdf | Canada | This report details the lack of equitable treatment in health care services for Indigenous languages speakers in the Northwest Territories based on surveys, complaints received by the Languages Commissioner of the Northwest Territories, and a review of legislation. It identifies that interpreting services are limited, and those that exist are not accessed enough or in a timely fashion, are low quality, and lack sensitivity to language needs. |
| Guthridge, S., Li, L., Silburn, S., Li, S. Q., McKenzie, J., & Lynch, J. (2016). Early influences on developmental outcomes among children, at age 5, in Australia’s Northern Territory. Early Childhood Research Quarterly, 35, 124–134. https://doi.org/10.1016/j.ecresq.2015.12.008 | Australia | Drawing on census data, this study reports that five year-old Aboriginal children in the Northern Territory who spoke English as a second language were more likely to be assessed as developmentally vulnerable in one or more Early Development Census domains than children who spoke English as their first language. The domains included physical health and wellbeing, emotional maturity, language and cognitive skills (school-based), communication skills and general knowledge. This finding was independent of other perinatal and socio-demographic factors. |
| Haggarty JM, Cernovsky Z, Bedard M, & Merskey H. (2008). Suicidality in a sample of arctic households. Suicide & Life-Threatening Behavior, 38(6), 699–707. https://doi.org/10.1521/suli.2008.38.6.699 | Canada | In this study, Inuit people living in a small hamlet in Nunavut completed surveys about their mental health in either English or Inuktitut. An association between survey language choice and suicidal ideation was reported. Younger respondents with high anxiety who chose to complete the survey in English were reported to be at significant risk for suicidal ideation or behaviour. The authors conclude that Indigenous language use is somewhat protective against suicidal ideation. Language choice on the survey was not associated with thoughts or actions of self-harm. |
| Hallett, D., Chandler, M. J., & Lalonde, C. E. (2007). Aboriginal language knowledge and youth suicide. Cognitive Development, 22(3), 392–399. https://doi.org/10.1016/j.cogdev.2007.02.001 | Canada | In this quantitative ecological study of 136 First Nations communities in British Columbia, a negative relationship was reported between a community-level measure of conversational language proficiency and youth suicide rate. Only one youth suicide was reported in five years for the 16 communities in which at least half the band members reported having a conversational knowledge of their own First Nations language. |
| Halseth, R. (2018). Overcoming barriers to culturally safe and appropriate dementia care services and supports for Indigenous peoples in Canada. National Collaborating Centre for Aboriginal Health. https://www.nccah-ccnsa.ca/docs/emerging/RPT-Culturally-Safe-Dementia-Care-Halseth-EN.pdf | Canada | This literature review reports that it is important to provide linguistically appropriate care and to incorporate language and culture into dementia care for Indigenous peoples to alleviate barriers and to improve wellness and quality of life. Language is identified as providing cognitive stimulation and being a source of strength and resilience. |
| Hamilton, R. M. (1996). A study of acculturation and depression in urban and rural native Alaskans [Doctoral dissertation, California School of Professional Psychology]. https://www.proquest.com/docview/304309965?pq-origsite=gscholar&fromopenview=true | USA | This study examines correlations between various aspects of acculturation and depression in urban and rural cohorts of Native Alaskans. A significant correlation is reported between level of depression and use of English outside of the family (i.e., as the language spoken with friends, of newspapers and magazines read, of music listened to, of radio stations listened to, of television programs watched, of jokes familiar with). |
| Hampton, J. W., & Castro, L. (2005). End-of-life issues for American Indians: A commentary. Journal of Cancer Education, 20(1, Suppl), 37–40. https://doi.org/10.1207/s15430154jce2001s_09 | USA | The authors of this commentary argue that determining the primary and secondary languages of American Indian palliative care patients is one of the seven key aspects in the “algorithm“ for discussing end-of-life issues and ensuring successful outcomes. |
| Han, P. K., Hagel, J., Welty, T. K., Ross, R., Leonardson, G., & Keckler, A. (1994). Cultural factors associated with health-risk behavior among the Cheyenne River Sioux. American Indian and Alaska Native Mental Health Research, 5(3), 15–29. | USA | This survey-based study conducted with adult members of the Cheyenne River Sioux tribe reports that females who were fluent in their Indigenous language exhibited better health-related behaviors than those who were not fluent. The same relationship was not reported for males. |
| Hanrahan, M. C. (2002). Identifying the needs of Innu and Inuit patients in urban health settings in Newidentifiedland and Labrador. Canadian Journal of Public Health, 93(2), 149–152. | Canada | In this study on the health care needs of Innu and Inuit people living in small, isolated villages in Canada, the shortage of language interpretation services in urban health care settings and hospitals is reported as a major issue impacting health outcomes. |
| Healey, G. K., & Meadows, L. M. (2008). Tradition and Culture: An Important Determinant of Inuit Women’s Health. Journal of Aboriginal Health, 4(1), 25–33. | Canada | In this study, Inuit women in Nunavut report that losing their language and traditional practices may lead to significant problems related to identity, social inclusion, and wellness, as well as people turning to addictive substances to cope. |
| Healthy Parks Healthy People SA Connection to Country for Aboriginal Health and Wellbeing Working Group. (2018). Joint statement of action: Connection to country for Aboriginal health and wellbeing. South Australian Health and Medical Research Institute (UniSA). https://cdn.environment.sa.gov.au/environment/docs/joint_statement_of_action_connection_to_country_for_aboriginal_health_and_wellbeing.pdf | Australia | In this joint statement of action on connection to country for Aboriginal health and wellbeing, one of the nine key principles for health is recognizing “culture and language as determinants of Aboriginal health and wellbeing”. The authors call on the government to increase the status and expand the use of Aboriginal languages within the public sector to create a system that “recognises the intrinsic link between culture, Country and language for Aboriginal people and embeds Aboriginal ways of seeing, doing and being across the sector. This includes funding the revival of Aboriginal languages across South Australia and establishing a mandate to teach Aboriginal history, culture and languages throughout the South Australian education system.” |
| Henderson, J. A. (2009). Factors associated with use of traditional healers in American Indians and Alaska natives. In M. Incayawar, R. Wintrob, L. Bouchard, & G. Bartocci (Eds.), Psychiatrists and traditional healers: Unwitting partners in global mental health. (pp. 93–106). Wiley-Blackwell. https://doi.org/10.1002/9780470741054.ch8 | USA | In this analysis of data from the 1987–1988 National Medical Expenditure Survey II, American Indian and Alaska Native people who used a traditional healer were 5.8 times more likely to speak English as a second language than those who did not use a traditional healer. The authors interpret this in the context of previous research to indicate that language is a measure of traditionalism, which itself is predictive of working with a traditional healer. |
| Hersh, D., Armstrong, E., McAllister, M., Ciccone, N., Katzenellenbogen, J., Coffin, J., Thompson, S., Hayward, C., Flicker, L., & Woods, D. (2019). General practitioners’ perceptions of their communication with Australian Aboriginal patients with acquired neurogenic communication disorders. Patient Education and Counseling, 102(12), 2310–2317. https://doi.org/10.1016/j.pec.2019.07.029 | Australia | In this qualitative study involving interviews and consultations with doctors from metropolitan Perth and five regional sites in Western Australia, a key theme is how challenges in communication with Aboriginal patients with acquired neurogenic communication disorders can be layered due to language differences, and that this can impede appropriate diagnosis and care. |
| Hilgendorf, A., Guy Reiter, A., Gauthier, J., Krueger, S., Beaumier, K., Corn, R. S., Moore, T. R., Roland, H., Wells, A., Pollard, E., Ansell, S., Oshkeshequoam, J., Adams, A., & Christens, B. D. (2019). Language, Culture, and Collectivism: Uniting Coalition Partners and Promoting Holistic Health in the Menominee Nation. Health Education & Behavior : The Official Publication of the Society for Public Health Education, 46(1_suppl), 81S-87S. https://doi.org/10.1177/1090198119859401 | USA | This article reports on a qualitative case study of the Menominee Wellness Initiative, an Indigenous public health coalition which made language, culture, and collective values the focus of their health promotion work. The positive impacts of anchoring health promotion activities in language included increased participation; nurturing community members’ sense of identity, hope, belonging, and purpose which was seen as supporting both emotional and physical well-being; and recognizing and attending to spiritual health. It also reminded people of their connections to foods which led to establishing a community farm. |
| Hill, D. M. (2013). Traditional Medicine and Restoration of Wellness Strategies. Journal of Aboriginal Health, 5(1). | Canada | Based on an overview of a selection of literature, the author reports that restoring and speaking Indigenous languages is a foundational component of strategies for healing and wellness. They describe the harms of language suppression on self-esteem and how language can be important in practicing traditional healing and medicine. |
| Hirchak, K. A., Leickly, E., Herron, J., Shaw, J., Skalisky, J., Dirks, L. G., Avey, J. P., McPherson, S., Nepom, J., Donovan, D., Buchwald, D., & McDonell, M. G. (2018). Focus groups to increase the cultural acceptability of a contingency management intervention for American Indian and Alaska Native Communities. Journal of Substance Abuse Treatment, 90, 57–63. https://doi.org/10.1016/j.jsat.2018.04.014 | USA | In focus groups about a treatment approach for alcohol use disorders that uses positive reinforcement, participants emphasized the need for cultural adaptations including incorporating Indigenous languages. The authors describe how they subsequently modified their treatment program to incorporate Indigenous languages. |
| Hodge, F. S., & Nandy, K. (2011). Predictors of wellness and American Indians. Journal of Health Care for the Poor and Underserved, 22(3), 791–803. https://doi.org/10.1353/hpu.2011.0093 | USA | In this survey study with people living on rural reservations in California, respondents who rated their wellness as good were more likely to speak their tribal language than those who rated their wellness as poor. |
| Hodge, F. S., Cantrell, B. G., & Kim, S. (2011). Health status and sociodemographic characteristics of the morbidly obese American Indians. Ethnicity & Disease, 21(1), 52–57. | USA | In this survey study with people living on rural reservations in California, no relationship is reported between morbid obesity and ability to speak one's Indigenous language. |
| Hossain, B., & Lamb, L. (2019). Cultural attachment and wellbeing among Canada’s Indigenous people: A rural urban divide. Journal of Happiness Studies: An Interdisciplinary Forum on Subjective Well-Being. https://doi.org/10.1007/s10902-019-00132-8 | Canada | In this analysis of data from the 2012 Aboriginal Peoples survey, a link is reported between higher cultural attachment (including ability to speak and understand an Indigenous language) and lower psychological distress, but only for people living in small cities and rural areas. The authors suggest that the urban/rural divide may be due to self-selection (i.e., people who believe that language and culture are more important tend to move to non-metropolitan areas) or associations between other factors related to living in these areas and wellbeing. |
| House of Representatives Standing Committee on Aboriginal and Torres Strait Islander Affairs. (2012). Our land our languages: Language learning in Indigenous communities (Australia) [Report]. House of Representatives Standing Committee on Aboriginal and Torres Strait Islander Affairs. https://apo.org.au/node/31133 | Australia | In this report drawing on public hearings and a survey, language is described as integral in affirming and maintaining wellbeing, self-esteem and a strong sense of identity for Aboriginal and Torres Strait Islander communities. The report describes how languages contain complex understandings of a person’s culture and their connection with their land, and keep people connected to culture which strengthens feelings of pride and self worth. The authors reference previous research that has reported positive associations between language and general health and wellbeing. |
| Huot, S H. Ho , A. Ko , S. Lam , P. Tactay , J. MacLachlan & R. K. Raanaas (2019) Identifying barriers to healthcare delivery and access in the Circumpolar North: important insights for health professionals, International Journal of Circumpolar Health, 78:1, 1571385, DOI: 10.1080/22423982.2019.1571385 | Canada  USA | In this scoping review of the barriers to healthcare delivery and access in the Circumpolar North, challenges posed by differences in culture and language between healthcare providers and recipients were addressed by 23 articles included in the review. |
| Ireland, S., & Maypilama, E. L. (2020). ‘We are sacred’: An intercultural and multilingual approach to understanding reproductive health literacy for Yolnu girls and women in remote Northern Australia. Health Promotion Journal of Australia, 9710936. https://doi.org/10.1002/hpja.439 | Australia | This qualitative study involving consultations with Yolŋu community members concludes that there is a need for a reproductive health literacy framework that privileges Yolŋu reproductive knowledge, practices, and language. |
| Ireland, Sarah, Narjic, Concepta Wulili, Suzanne Belton, Sherry Saggers & Ann McGrath (2015) ‘Jumping around’: exploring young women's behaviour and knowledge in relation to sexual health in a remote Aboriginal Australian community, Culture, Health & Sexuality, 17:1, 1-16, DOI: 10.1080/13691058.2014.937747 | Australia | In this ethnographic study in a remote Aboriginal community in the Northern Territory, the authors report that not speaking English as a first language compounds the issue of very limited biomedical understandings of sexually transmitted infections and contraception among the young women. The article also reports that differences between sexual health terminology in the local Indigenous language and in English shape cultural understandings. |
| Isaacson, M. J. (2018). Addressing palliative and end-of-life care needs with Native American elders. International Journal of Palliative Nursing, 24(4), 160–168. https://doi.org/10.12968/ijpn.2018.24.4.160 | USA | In this qualitative study about palliative and hospice care with members of a Northern Plains reservation community, results strongly emphasize the need for health care workers at the facility to learn the local language in order to provide effective and culturally-appropriate care. There was a need for fluent interpreters who are available 24/7. The author also discusses the link between understanding the language and understanding the culture. |
| Iwama, M., Marshall, M., Marshall, A., & Bartlett, C. (2009). Two-Eyed Seeing and the Language of Healing in Community-Based Research. Canadian Journal of Native Education, 32(2), 3–23. | Canada | This article provides an account of working together toward healing through the revitalization of the Mi’kmaq language in a community participation model of research and teaching. The authors describe that “diminished fluency threatens the linguistic matrix that creates and sustains the health of individuals in community,” including the use of the healing and spiritual verb tenses of the language. Loss of traditional language impacts connectedness, which damages health. |
| Jacklin, K., & Walker, J. (2012). Trends in Alzheimer’s disease and related dementias among First Nations and Inuit. Health Canada. https://141419f0-5602-433d-85d2-4d5a8ecfd5ec.filesusr.com/ugd/6e29de_05ec1795a75a41b08a0119a561221420.pdf | Canada | In this literature review, the authors identify that “there is limited but promising evidence that the use of Indigenous language in the care of people with dementia may have benefits to the quality of care and the quality of life experienced by the patient. Use of Indigenous language may also prove to be an important prevention tool.” They identify Indigenous language use as a cultural system that may potentially improve or sustain cognitive function in old age. They also report a widespread desire to have services available close to home so that culture and language barriers may be minimized. |
| Jacklin, K., & Warry, W. (2010). Forgetting and Forgotten: Dementia in Aboriginal Seniors. Forgetting and Forgotten: Dementia in Aboriginal Seniors. AAA Annual Meeting. https://doi.org/10.1037/e682032011-001 | Canada | This study involving interviews and focus groups in six Aboriginal communities in Ontario reports that using Aboriginal languages is an important part of caring for people with Alzheimer's disease and related dementias. Listening to language tapes and speaking the language are reported to keep the mind healthy. |
| Jacob, M. M., Sabzalian, L., Johnson, S. R., Jansen, J., & Morse, G. S. (2019). ‘We Need to Make Action NOW, to Help Keep the Language Alive’: Navigating Tensions of Engaging Indigenous Educational Values in University Education. American Journal of Community Psychology, 64(1/2), 126–136. https://doi.org/10.1002/ajcp.12374 | USA | In this study in which researchers analyzed the daily journals of students enrolled in an intensive two-week Indigenous language education summer institute, the authors report that Indigenous language education is crucial for advancing K-12 education practices that honor Indigenous community psychological well-being, and assert that Indigenous community language educators do important survivance work. |
| James, H. (2017). Aboriginal language, identity formation and health [Undergraduate research practicuum deliverable]. First Nations and Indigenous Studies, University of British Columbia and the First Peoples’ Cultural Council. https://fpcc.ca/wp-content/uploads/2020/08/Aboriginal_Language_Identity_Formation_and_Health.pdf | Australia  Canada  USA | The author draws on previous literature to demonstrate that historical trauma stemming from colonization (including ancestral language loss) impacts health, and that ancestral language reintegration and/or revitalization can be effective treatment and prevention strategies. |
| Janssen, I., Lévesque, L., & Xu, F. (2014). Correlates of physical activity among First Nations children residing in First Nations communities in Canada. Canadian Journal of Public Health, 105(6), e412-7. https://doi.org/10.17269/cjph.105.4526 | Canada | This analysis of data from the First Nations Regional Health Survey identified that children who understood and used their First Nations language most often in daily life were were more likely to participate in traditional First Nations physical activities (e.g., berry picking, hunting/trapping, fishing, canoeing/kayaking) than those who used their language less. |
| Jenni, B., Anisman, A., McIvor, O., & Jacobs, P. (2017). An Exploration of the Effects of Mentor-Apprentice Programs on Mentors’ and Apprentices’ Wellbeing. International Journal of Indigenous Health, 12(2), 25–42. https://doi.org/10.18357/ijih122201717783 | Canada | In this study, researchers conducted interviews with participants in a Mentor-Apprentice type program. They report positive impacts on cultural and spiritual health and healing, as well as on health outcomes including recovery from drug and alcohol addictions and breaking alcohol dependency patterns in the family. The authors describe how language loss negatively impacts wellbeing, and that the Elders experienced healing through becoming language mentors. Potentially detrimental effects on apprentices’ well being which resulted from their increasingly busy schedules were fatigue and feelings of exhaustion. |
| Kahn-John, M., Badger, T., McEwen, M. M., Koithan, M., Arnault, D. S., & Chico-Jarillo, T. M. (2020). The Dine (Navajo) Hozho Lifeway: A Focused Ethnography on Intergenerational Understanding of American Indian Cultural Wisdom. Journal of Transcultural Nursing, 32(3), 256–265. https://doi.org/10.1177/1043659620920679 | USA | This ethnographic study with Navajo people reports that listening to and speaking their traditional language is one way that the Diné understanding of cultural wisdom is transferred. The people interviewed believe that cultural wisdom is a health-sustaining protective factor, and as such the past restrictions against speaking Indigenous languages and the current language barriers between generations have had detrimental impacts on health. |
| Kaufert, J. M., Putsch, R. W., & Lavallée, M. (1999). End-of-life decision making among Aboriginal Canadians: Interpretation, mediation, and discord in the communication of ‘bad news’. Journal of Palliative Care, 15(1), 31–38. | Canada | In this study in Winnipeg hospitals, a sample of cases were studied which involved communication in end-of life decisions and intervention by professional interpreters. Results highlight how Aboriginal language interpreters are critical to facilitating communication about end-of-life decision making with patients, including understanding and incorporating culture. It also describes the problems that arise when there is a lack of access to language translation in this process. |
| Kaufert, Joseph M., and Evelyn Shapiro. ‘Cultural, Linguistic and Contextual Factors in Validating the Mental Status Questionnaire: The Experience of Aboriginal Elders in Manitoba’. Transcultural Psychiatric Research Review, vol. 33, no. 3, 1996, pp. 277–96, https://doi.org/10.1177/136346159603300302. | Canada | This study on the experiences of Aboriginal Elders in Manitoba addresses how linguistic variables impact the validity of mental status instruments, including instruments that are used to screen for dementia. |
| Kelly, L., & Minty, A. (2007). End-of-life issues for aboriginal patients: A literature review. Canadian Family Physician Medecin de Famille Canadien, 53(9), 1459–1465. | Canada  USA | This article reviews the literature that describes how language barriers between Indigenous patients and healthcare providers can impact end-of-life care, and that there is a need to use professional interpreters instead of family members. |
| King, C., Atwood, S., Lozada, M., Nelson, A. K., Brown, C., Sabo, S., Curley, C., Muskett, O., Orav, E. J., & Shin, S. (2018). Identifying risk factors for 30-day readmission events among American Indian patients with diabetes in the Four Corners region of the southwest from 2009 to 2016. PloS One, 13(8), e0195476. https://doi.org/10.1371/journal.pone.0195476 | USA | This qualitative study looking at health records over a five year study period reported that the American Indian patients with diabetes who were admitted to a medical center and indicated preference for an American Indian Language (Zuni or Navajo) were less likely to be readmitted within 30 days than those who indicated a preference for English. |
| Kirmayer, L. J., Brass, G. M., Holton, T., Paul, K., Simpson, C., & Tait, C. (2007). Suicide Among Aboriginal People in Canada. The Aboriginal Healing foundation. | Canada | In this report that reviews previous literature to understand the origins of suicide and identify effective interventions for Aboriginal communities, the role of language is discussed in four main contexts: the transgenerational impacts of the loss of language stemming from residential schools; the self-estrangement and loss of self-esteem that can come from the denigration and marginalization of cultural heritage, including language; the alienation from loss of language; and, the promise of cultural initiatives including language learning for suicide prevention. A lack of availability of suicide prevention/intervention services in Indigenous language is also reported. |
| Kwan, P. P., Soniega-Sherwood, J., Esmundo, S., Watts, J., Pike, J., Sabado-Liwag, M., & Palmer, P. H. (2020). Access and utilization of mental health services among Pacific Islanders. Asian American Journal of Psychology, 11(2), 69–78. https://doi.org/10.1037/aap0000172 | USA | This interview study in Southern California reports that culture and language barriers were a reason that the Pacific Islander adults interviewed did not often access and use necessary mental health care. They discussed how language barriers can prevent individuals from communicating well with care providers and from understanding the health information with which they have been provided. People who only spoke Samoan or Tongan reported having a difficult time seeking help because of the lack of providers that speak their language and the absence of health education materials in their native language. |
| Lang, C., Macdonald, M. E., Carnevale, F., Josée Lévesque, M., & Decoursay, A. (2010). Kadiminekak kiwabigonem: Barriers and facilitators to fostering community involvement in a prenatal program in an Algonquin community. Pimatisiwin: A Journal of Aboriginal & Indigenous Community Health, 8(1), 55–81. | Canada | This study reports that language and literacy was one of the five barriers to fostering community involvement in the development and implementation of a pre and postnatal program in the Rapid Lake Algonquin community, because the program is normally offered in English even though Algonquin is the first language of the community. The article describes how the language was subsequently successfully incorporated into the program. |
| Larson, S., Stoeckl, N., Jarvis, D., Addison, J., Grainger, D., Watkin Lui, F., Walalakoo Aboriginal Corporation, Bunuba Dawangarri Aboriginal Corporation Rntbc, Ewamian Aboriginal Corporation Rntbc, & Yanunijarra Aboriginal Corporation Rntbc. (2020). Indigenous Land and Sea Management Programs (ILSMPs) Enhance the Wellbeing of Indigenous Australians. International Journal of Environmental Research and Public Health, 17(1). https://doi.org/10.3390/ijerph17010125 | Australia | In this study involving interviews with members of four Indigenous communities involved in Indigenous land and sea management programs in north Queensland and Western Australia, participants identified “Making sure language is not ‘lost’ (spoken regularly and/or written down) “as one of the four most important factors for the wellbeing of their communities. |
| Latycheva, O., Chera, R., Hampson, C., Masuda, J. R., Stewart, M., Elliott, S. J., & Fenton, N. E. (2013). Engaging First Nation and Inuit communities in asthma management and control: Assessing cultural appropriateness of educational resources. Rural & Remote Health, 13(2), 1–11. | Canada | In this study involving surveys and discussions with participants from First Nations and Inuit communities across Canada, it is reported that tailoring educational materials about asthma management and control to include their languages would increase their uptake. |
| Lévesque, Lucie, et al. ‘Correlates of Physical Activity in First Nations Youth Residing in First Nations and Northern Communities in Canada.’ Canadian Journal of Public Health, vol. 106, no. 2, Feb. 2015, pp. e29-35, https://doi.org/10.17269/cjph.106.4567. | Canada | This secondary analysis of the First Nations Regional Health Survey reports that youth who understood and used their First Nations language most often in daily life were more likely to participate in traditional First Nations physical activities (e.g., berry picking, hunting/trapping, fishing, canoeing/kayaking). |
| Liebenberg, Linda, et al. ‘“It’s Just Part of My Culture“: Understanding Language and Land in the Resilience Processes of Aboriginal Youth’. Youth Resilience and Culture: Commonalities and Complexities., edited by Linda C. Theron et al., vol. 11, Springer Science + Business Media, 2015, pp. 105–16, https://doi.org/10.1007/978-94-017-9415-2_8. | Canada | This chapter explores the potential for policy and governance aimed at the revival of traditional culture to improve youth well-being in remote Aboriginal communities, with particular impacts on resilience. From the perspectives of youth, providing youth with language and land-based activities is key for developing resilience. |
| Lowell, A. (2013). ‘From your own thinking you can’t help us’: Intercultural collaboration to address inequities in services for indigenous Australians in response to the World Report on Disability. International Journal of Speech-Language Pathology, 15(1), 101–105. https://doi.org/10.3109/17549507.2012.725770 | Australia | This commentary reports that a lack of linguistically appropriate speech-language pathology health services for Indigenous people in the Northern Territory creates inequities including misdiagnosis of communication disabilities and inappropriate care. |
| Lowell, Anne, et al. “Finding a Pathway and Making It Strong: Learning from Yolŋu about Meaningful Health Education in a Remote Indigenous Australian Context.” Health Promotion Journal of Australia : Official Journal of Australian Association of Health Promotion Professionals, no. 9710936, 2020, https://doi.org/10.1002/hpja.405. | Australia | This study involving participant observation and semi-structured interviews with educators and community members reports that integrating Yolŋu and biomedical knowledges to build and share meaningful, in-depth (not simplified or directive) oral explanations in local languages with Yolŋu patients is key to improving effective communication about chronic conditions. The article also describes the successful implementation of these findings. |
| Lowitja Institute. We Nurture Our Culture for Our Future, and Our Culture Nurtures Us. Report, Australian Human Rights Commission, 19 Mar. 2020. Australia, apo.org.au, https://apo.org.au/node/302867. | Australia | This report addresses the question of how to shift institutions and thinking beyond medical models of health towards a model that locates culture as the foundation for good health and wellbeing. Language is discussed throughout the report as part of this relationship to health, particularly as a foundation of Indigenous beliefs and knowledge, cultural expression, cultural identity, and cultural continuity. |
| Maina, Geoffrey, et al. “A Scoping Review of School-Based Indigenous Substance Use Prevention in Preteens (7-13 Years).” Substance Abuse Treatment, Prevention & Policy, vol. 15, no. 1, 2020, p. N.PAG-N.PAG. ccm, EBSCOhost, https://doi.org/10.1186/s13011-020-00314-1. | Australia  Canada  USA | This scoping review study reports that centering Indigenous languages in school-based substance use prevention curricula for Indigenous preteens is one factor that enhanced the effectiveness, appropriateness, and sustainability of the programs. |
| Maranzan, A. (2009). The health status and needs of aboriginal people assessed for home care in Ontario [Doctoral dissertation, Lakehead University]. https://knowledgecommons.lakeheadu.ca/handle/2453/3877 | Canada | In this study, quantitative analyses were carried out on data about all clients in Ontario assessed for home care in a one year period, and then interviews were conducted with home care coordinators and providers for Aboriginal people in Northwestern Ontario. Language was identified as a significant challenge for home care service coordination and provision for Aboriginal peoples who primarily spoke their traditional language. In particular it was difficult to coordinate having a translator visit the client's home and then to translate certain English concepts and information into their languages, such as about assessments and procedures. |
| Marmion, Doug, et al. Community, Identity, Wellbeing: The Report of the Second National Indigenous Languages Survey. 2014, p. 79. | Australia | In this analysis of the Second National Indigenous Languages Survey, it is identified that traditional languages are a strong part of Aboriginal and Torres Strait Islander peoples' identities, and that connection with language is critical for their wellbeing. The authors also identify that the goals of language activities are not just to increase speaker numbers and revitalize or maintain languages, but that they are also about helping people connect with language and culture and improving wellbeing. |
| McBeath, Brittany. Conceptualization of Community Wellness in Three First Nations Communities. 12 May 2020. Queen's University. https://qspace.library.queensu.ca/handle/1974/27813. | Canada | In this study involving talking circles with three First Nations communities in Manitoba and Ontario, all communities identified the revitalization of language as a component of community wellness. |
| McCabe, M., Morgan, F., Smith, M., Yazzie, E., Spencer, A., Curley, H., Begay, R., & Gohdes, D. (2003). Lessons learned: Challenges in interpreting diabetes concepts in the Navajo language. Diabetes Care, 26(6), 1913–1914. ccm. https://doi.org/10.2337/diacare.26.6.1913 | USA | This commentary describes how translating medical diabetes concepts from English to Navajo is challenging but important for improving communication with patients. |
| McGrath P. (2006). Exploring Aboriginal peoples’ experience of relocation for treatment during end-of-life care. International Journal of Palliative Nursing, 12(3), 102–108. https://doi.org/10.12968/ijpn.2006.12.3.20692 | Australia | This interview study with Aboriginal patients, carers, and health professionals in the regional, rural and remote areas of the Northern Territory reports that Aboriginal language barriers are one factor that contribute to the frightening experience of relocation away from one's community for medical treatment during end-of-life care. |
| McIntyre I, Broughen C, Trepman E, & Embil JM. (2007). Foot and ankle problems of Aboriginal and non-Aboriginal diabetic patients with end-stage renal disease. Foot & Ankle International, 28(6), 674–686. | Canada | In this study involving retrospective reviews of medical records, interviews, and physical examinations on diabetic patients with end-stage kidney disease in Winnipeg, Manitoba, one factor driving a higher prevalence of foot and ankle problems in Aboriginal than non-Aboriginal patients was language barriers. While 4 out of 11 Aboriginal patients cited language barriers as a reason for inadequate foot care and footwear, none of the 22 non-Aboriginal patients cited experiencing this challenge. |
| McIvor, O., Napoleon, A., & Dickie, K. M. (2009). Language and Culture as Protective Factors for At-Risk Communities. Journal of Aboriginal Health, 5(1), 6–25. https://doi.org/10.18357/ijih51200912327  McIvor, O. (2013). Protective effects of language learning, use and culture on the health and well- being of Indigenous people in Canada. Proceedings of the 17th foundation of endangered Languages (FEL) Conference. FEL XVII: Endangered Languages Beyond Boundaries: Community Connections, Collaborative Approaches and Cross-Disciplinary Research, Carleton University. <http://www.ogmios.org/conferences/proceed2013.htm> | Canada | This literature review study reports that most of the existing literature on the topic of traditional language use and health focuses on the effect of Indigenous-only language use in the home which in turn lowers rates of access to health care. The authors also cite three studies demonstrating protective effects of language on mental health and suicide. |
| McKenzie, James. “Approaching From Many Angles: Seeing the Connections for Our Languages to Live.” Modern Language Journal, vol. 104, no. 2, June 2020, pp. 501–06. | Canada  USA | Drawing on the author's experiences as a “Navajo language revitalizer,” this paper discusses how speaking and revitalizing Indigenous languages is healing. One reason cited is Tlingit scholar X̲ʼunei Lance Twitchell's concept of Indigenous counterhegemonic transformation, which “seeks to expel cultural guilt & shame, external value systems, racist hierarchies & structures, and lateral oppression & violence by embracing respect, healthy communication, kindness, and unfragmented existence.” The author speaks to the link between the decline of speaking Indigenous languages and negative psychosocial effects. |
| McWhirter, J. Jeffries, and Cynthia A. Ryan. “Counseling the Navajo: Cultural Understanding.” Journal of Multicultural Counseling and Development, vol. 19, no. 2, Apr. 1991, pp. 74–82. psyh, 1991-25532-001, EBSCOhost, https://doi.org/10.1002/j.2161-1912.1991.tb00624.x. | USA | In this article, the authors discuss the challenges of providing English-language counseling to people who speak Navajo as a first language, as it can impair communication and there are certain concepts that can't be translated between the languages. This can add further complications to addressing the mental health challenges that patients present with. |
| Meisel, S., and K. Kiely. “Graphic Prescription Labels.” American Journal of Hospital Pharmacy, vol. 38, no. 8, 1981, p. 1116. | USA | This short article addresses the challenges of communicating important medication information to patients who only speak the Inuit language, and the authors present medication prescription labels that they developed which use images and symbols instead of words to overcome this barrier in conjunction with the use of translators. |
| Mercille, Genevieve, et al. “Household Food Insecurity and Canadian Aboriginal Women’s Self-Efficacy in Food Preparation.” Canadian Journal of Dietetic Practice and Research : A Publication of Dietitians of Canada = Revue Canadienne de La Pratique et de La Recherche En Dietetique : Une Publication Des Dietetistes Du Canada, vol. 73, no. 3, 2012, pp. 134–40. | Canada | In this survey study in a semi-isolated reserve community in Quebec, Atikamekw women who rated their understanding of their native language as “very good“ were reported to have better self-efficacy in healthy food preparation when compared with women who rated their understanding as “good.” |
| Mihychuk, Hon. MaryAnn. Breaking Point : The Suicide Crisis in Indigenous Communities. Report of the Standing Committee OnIndigenous and Northern Affairs. June 2017, https://www.deslibris.ca/ID/10092516#details=1. | Canada | Drawing on community testimonies, this report identifies Indigenous language use, retention, and revitalization as critical for suicide prevention in Indigenous communities, and the erosion of the languages as a key challenge for communities. |
| Mitchell AG, Lowell A, Ralph AP. Report on the Patient Educator service at Royal Darwin Hospital, 2001-2009: insights into inter-cultural communication in healthcare. Darwin: Aboriginal Resource and Development Services, 2016. https://www.researchgate.net/publication/304121921_Report_on_the_Patient_Educator_service_at_Royal_Darwin_Hospital_2001-2009_insights_into_inter-cultural_communication_in_healthcare | Australia | The case studies presented in this study from Yolŋu Matha-speaking Patient Educators in Australia's Northern Territory highlight that a high extent of miscommunication between care providers and patients due to linguistic and cultural barriers was causing adverse outcomes. These included patients experiencing prolonged hospitalization due to confusion about management plans, consenting for procedures they did not understand, taking medications incorrectly, not adhering to isolation requirements, and being suspicious/distrustful of healthcare providers. Effective use of Patient Educators dramatically changed patients’ understanding of their conditions, often resulting in major changes in health-related decisions. Patients expressed being very grateful to receive “the full story”. |
| Mitchell, Alice G., Suzanne Belton, Vanessa Johnston, Wopurruwuy Gondarra & Anna P. Ralph (2019) “That Heart Sickness“: Young Aboriginal People’s Understanding ofRheumatic Fever, Medical Anthropology, 38:1, 1-14, DOI: 10.1080/01459740.2018.1482549 | Australia | This ethnographic study in the northern coastal and hinterland regions of the Northern Territory reports that the young Aboriginal participants’ understandings of medical information provided about acute rheumatic fever and rheumatic heart disease was constrained by the clinicians’ use of language rooted in biomedicine and delivered in English, which was a second language for all participants. |
| Mithen, Vincent, et al. “Aboriginal Patient and Interpreter Perspectives on the Delivery of Culturally Safe Hospital-Based Care.” Health Promotion Journal of Australia : Official Journal of Australian Association of Health Promotion Professionals, no. 9710936, 2020, https://doi.org/10.1002/hpja.415. | Australia | This study in the Northern Territory drawing on Aboriginal patient and interpreter perspectives on the delivery of culturally safe hospital-based care reports that Aboriginal patient needs can be better supported by improving systems approaches. These included better documentation of patient language and greater efficiency in use of interpreter services. |
| MJD foundation. (2019). How to improve the National Disability Insurance Scheme for Aboriginal people in remote Australia [Submission on the National Disability Insurance Scheme]. https://apo.org.au/node/270581 | Australia | This report on improving the national disability insurance scheme for Aboriginal people in remote Australia identifies funding interpreter support as a priority area for immediate attention by an incoming government. They describe that as English is not the first language for many Aboriginal people with disabilities in the Northern Territory of Australia, patients without access to interpreters cannot exercise their choice, achieve their goals, or effectively use the funding in their plans. |
| Møller, Helle (2013) “Double culturedness“: the “capital“ of Inuit nurses,International Journal of Circumpolar Health, 72:1, 21266, DOI: 10.3402/ijch.v72i0.21266  Møller, Helle. ‘You Need to Be Double Cultured to Function Here’: Toward an Anthropology of Inuit Nursing in Greenland and Nunavut. University of Alberta, 2011. | Canada | This qualitative, ethnographic study reports that the “double culturedness” of Inuit nurses in Nunavut, including their ability to speak the Inuit language, improves their ability to provide health care as they can better understand, negotiate, and interact with patients. |
| Møller, Helle. “Culturally Safe Communication and the Power of Language in Arctic Nursing.” Communication Culturellement Sécuritaire et Pouvoir de La Langue Dans Les Soins Infirmiers de l’Arctique., vol. 40, no. 1, Jan. 2016, pp. 85–104. fph. | Canada | This study involving interviews with Inuit nurses and nursing students reports that being able to speak the Inuit language is a part of culturally safe nursing care that needs to be implemented in conjunction with knowledge about Inuit history and colonialism and cognizant of the power relations inherent in the provider interaction. The author asserts that Inuit nurses are invaluable for creating culturally safe care environments in the Arctic. |
| Moore, Sylvia (2019) Language and identity in an Indigenous teacher education program, International Journal of Circumpolar Health, 78:2, 1506213, DOI:10.1080/22423982.2018.1506213 | Canada | This article provides a case study of students training to become Inuktitut language teachers to examine the role of Inuktitut in post-secondary education. The author identifies that language learning may contribute to an increased awareness of, and connection to, one's Indigenous group, and that this in turn can enhance wellbeing. The author also asserts that language and the ability to connect to one’s culture are important determinants of health, and the loss of language and resulting cultural erosion has contributed to poorer health outcomes for many Indigenous peoples in Canada. The author posits that language rejuvenation programs such as the Inuit Bachelor of Education program can be understood as health promotion strategies. |
| Morcom, L. A. (2017). Self-esteem and cultural identity in Aboriginal language immersion kindergarteners. Journal of Language, Identity, and Education, 16(6), 365–380. https://doi.org/10.1080/15348458.2017.1366271 | Canada | In this quantitative study, the personal and collective self-esteem of a group of 10 kindergarteners enrolled in an Anishinaabemowin (Ojibwe) immersion education program in Manitoulin Island, Ontario, was measured and compared to a previous study with Inuit children. It is reported that the Anishinaabemowin immersion program had a positive impact on both aspects of self-esteem. |
| Mounga, Va, and Erin Maughan. ‘Breast Cancer in Pacific Islander Women: Overcoming Barriers to Screening and Treatment.’ Nursing for Women’s Health, vol. 16, no. 1, 2012, pp. 26–35, https://doi.org/10.1111/j.1751-486X.2012.01697.x. | USA | This literature review reports that providing language-appropriate health education materials to native Hawaiian, Samoan and Tongan women living in the continental United States reduces breast cancer mortality rates. |
| Myers, T., Bullock, S. L., Calzavara, L. M., Cockerill, R., Marshall, V. W., & George-Mandoka, C. (1999). Culture and sexual practices in response to HIV among Aboriginal people living on-reserve in Ontario. Culture, Health & Sexuality, 1(1), 19–37. https://doi.org/10.1080/136910599301148 | Canada | In this quantitative study involving surveys of Aboriginal people living on eleven reserves in Ontario, no relationship was reported between ability to speak an Indigenous language and engagement in high-risk sexual behaviors among Aboriginal people living on-reserve in Ontario. |
| Nasreen, Sharifa, et al. “Are Indigenous Determinants of Health Associated with Self-Reported Health Professional-Diagnosed Anxiety Disorders among Canadian First Nations Adults?: Findings from the 2012 Aboriginal Peoples Survey.” Community Mental Health Journal, vol. 54, no. 4, May 2018, pp. 460–68. psyh, 2018-18331-001, EBSCOhost, https://doi.org/10.1007/s10597-017-0165-0. | Canada | This analysis of data from the 2012 Aboriginal Peoples Survey did not report any association between ability to speak a First Nations language and being diagnosed with an anxiety disorder among adults. |
| National Collaborating Centre for Aboriginal Health. (2012). The State of Knowledge of Aboriginal Health: A Review of Aboriginal Public Health in Canada. National Collaborating Centre for Aboriginal Health. https://www.ccnsa-nccah.ca/docs/context/RPT-StateKnowledgeReview-EN.pdf | Canada | This report draws on previous literature and summarizes existing programs and services to identify that language is a social determinant of Indigenous peoples' health in Canada and is essential to improving health outcomes. Maintenance of language and cultural traditions contributes to individual and community identity, and is central to perceptions and experiences of health and illness. Language suppression affects self-identity, well-being, self-esteem and empowerment, all of which are identified as key factors in individual and community healing. Speaking only an Indigenous language is a unique barrier to health care for Aboriginal people living in rural and remote locations, and a lack of translation services can add additional stress for these patients. |
| National Collaborating Centre for Aboriginal Health. (2016). Culture and language as social determinants of First Nations, Inuit, and Métis health (Social Determinants of Health). | Canada | This fact sheet draws on examples of current language initiatives, existing literature, and national survey data to argue that language is a social determinant of First Nations, Inuit and Métis health. Language and culture are sources of healing and resilience (both protective and remedial), influence perceptions and experiences of health and illness and impact the care they may receive in health care settings, can ease intergenerational traumas, promote holistic healing, and rebuild self-esteem. |
| Newell, Sarah Lynn, et al. ‘Cultural Continuity and Inuit Health in Arctic Canada.’ Journal of Epidemiology and Community Health, vol. 74, no. 1, 2020, pp. 64–70, https://doi.org/10.1136/jech-2018-211856. | Canada | This analysis of data from the Arctic Supplement of the Aboriginal Peoples' Survey reports that among people who spoke even a few words of Inuktitut or Inuinnaqtun, there was an association between having good self-rated health and having access to at least one government service in their language. |
| Nicholls, Margaret (2003) What Motivates Intergenerational Practices in Aotearoa/New Zealand?, Journal of Intergenerational Relationships, 1:1, 179-181, DOI: 10.1300/ J194v01n01_17 | New Zealand | This article describes how *te kohanga reo*, the Māori language nest, is based on the principles of support, respect, love and acceptance of everyone involved in the children’s well being, and that it is vital to the well being, health and benefit of not only the family, but also to the whole of Māori society. *taha tina*, meaning health of body and mind, is a fundamental value underlying *te kohanga reo*. |
| Noreen, Willows, Louise Johnson-Down, Moubarac Jean-Claude, Michel Lucas, Elizabeth Robinson & Malek Batal (2018) Factors associated with the intake of traditional foods in the Eeyou Istchee (Cree) of northern Quebec include age, speaking the Cree language and food sovereignty indicators, International Journal of Circumpolar Health, 77:1, 1536251, DOI: 10.1080/22423982.2018.1536251 | Canada | In this quantitative survey study with Eeyou Istchee (Cree) people in northern Quebec, an association was reported between only speaking Cree at home and eating more traditional foods for males, but not for females. |
| Nova Scotia Aboriginal Home Care Steering Committee. (2010). Aboriginal Long Term Care in Nova Scotia. https://novascotia.ca/dhw/ccs/documents/Aboriginal-Long-Term-Care-in-Nova-Scotia.pdf | Canada | In this study about the provision of long term care for First Nations people living on-reserve in Nova Scotia, several communities noted the importance of language and that long term care residents, especially seniors suffering from dementia, would not be able to communicate with facility staff or other residents without interpreter support. A culturally relevant long term care model includes having staff that can understand and speak the language; this was seen as a minimum requirement for improving current long term care facilities. This study reports that communicating and information sharing with the elderly is optimal if conducted in their first language. |
| Oldfield, J., & Jackson, T. (2019). Childhood abuse or trauma: A racial perspective. Children Australia, 44(1), 42–48. https://doi.org/10.1017/cha.2018.48 | Australia  New Zealand  USA | This literature review examines the trauma faced by Aboriginal children in relation to colonization and racism, and particularly institutionalized racism. It reports that the destruction and diminishment of Indigenous languages is a source of racial trauma, and that such trauma can be mitigated through education, by adopting specialized Indigenous language and cultural curricula. |
| Oldfield, Janine Gai. ‘Anangu Muru Wunka - Talking Black Fella : A Critical Policy Analysis of the Northern Territory Compulsory Teaching in English for the First Four Hours of Each School Day’, 2016. Australian Education Index, 2255540205; 220459. | Australia | This study investigated the effects of a 2008 Northern Territory of Australia education language policy that mandated teaching in English for the first four hours of each school day, with focus on two remote Indigenous communities. It is reported that a community that suspended its bilingual program in response to this policy complained of more negative academic, well-being, behavioral, and cognitive issues among the children and a deterioration in resilience when compared to the community that was able to continue its bilingual program. |
| Ontario Centres for Learning, Research and Innovation in Long-Term Care and Sue Cragg Consulting. Supporting Indigenous Culture in Ontario’s Long-Term Care Homes: Needs Assessment and Ideas for 2017-18. (2017). https://clri-ltc.ca/resource/supporting-Indigenous-culture-in-ontarios-long-term-care-homes-needs-assessment/ | Canada | In this report drawing on literature and consultations with guidance by an expert Advisory Group, access to long-term care in an Indigenous patient's language is reported as a key part of culturally safe care that improves quality of life for long-term care patients in Ontario. Literature is cited highlighting how communication in the resident’s own language that is culturally specific is related to physical and mental health benefits such as reduced social isolation, lower rates of depression, fewer falls and hospitalizations, an improved likelihood of following medication guidelines and understanding medical decision making, and can minimize misdiagnosis. Patients may be vulnerable to inappropriate care because of language barriers. This can be especially important when, in the later stages of dementia, a resident uses only their first learned language. |
| Ontario Native Education Counselling Association. Indigenous Well-Being in Schools: Understanding, Promoting and Supporting Indigenous Learners. 2018, https://oneca.com/documents/Wellbeing_Final%20Report_WEB.pdf. | Canada | Language is mentioned throughout this report that explores how to promote the well-being of Indigenous students in Ontario schools. A positive sense of identity and the collective well-being of a community are identified as important and as being deeply rooted in the connection to land, language, culture, and places that connect families and communities. A system that supports all learners to experience Indigenous language was seen as supporting the well-being of Indigenous students. |
| Oster, Richard T., et al. ‘Cultural Continuity, Traditional Indigenous Language, and Diabetes in Alberta First Nations: A Mixed Methods Study’. International Journal for Equity in Health, vol. 13, no. 1, Oct. 2014, p. 92. BioMed Central, https://doi.org/10.1186/s12939-014-0092-4. | Canada | This mixed-mode study combined interviews with Cree and Blackfoot political leaders with an analysis of provincial administrative data for Alberta First Nations communities to examine the relationships between language (as a proxy measure of cultural continuity) and diabetes prevalence. The authors report a strong association between high community language knowledge and lower diabetes prevalence in the communities that was independent of median household income, unemployment rate, and high school completion rate. |
| Pauktuutit Inuit Women of Canada. Tukisiviit: Do You Understand? (2012). [Tukisiviit Summary Report 2012-2014]. Pauktuutit Inuit Women of Canada. https://www.deslibris.ca/ID/242943#details=1 | Canada | This report on a national forum documents how providing care in Inuktitut using newly developed, accurate Inuktitut HIV/AIDS terminology improved communication with healthcare providers as well as compliance, and improved the standard of care overall. Additionally, accurate terminology is reported to contribute to language preservation and cultural identity. |
| Pawu-Kurlpurlurnu WJ, Holmes M and Box L. 2008. Ngurra-kurlu: A way of working with Warlpiri people, DKCRC Report 41. Desert Knowledge CRC, Alice Springs. https://www.nintione.com.au/resource/DKCRC-Report-41-Ngurra-kurlu.pdf | Australia | The authors of this report present the concept of *Ngurra-kurlu* which contains five key interrelated elements of Warlpiri culture: language, land, law, ceremony, and skin. When *Ngurra-kurlu* is supported, it creates a system that is conducive to the healthy functioning of Warlpiri people and country. |
| Pearce, M. E., et al. ‘The Cedar Project: Exploring Determinants of Psychological Distress among Young Indigenous People Who Use Drugs in Three Canadian Cities’. Global Mental Health, vol. 5, Oct. 2018. https://doi.org/10.1017/gmh.2018.26. | Canada | In this study, two questionnaires were administered to a cohort of young Indigenous peoples who use illicit drugs in three cities in British Columbia. Results from quantitative analyses suggest a marginal association between being able to speak an Indigenous language and lower psychological distress among males. The same association was not reported among females. |
| Pearce, Margo E., et al. ‘The Cedar Project: Resilience in the Face of HIV Vulnerability within a Cohort Study Involving Young Indigenous People Who Use Drugs in Three Canadian Cities.’ BMC Public Health, vol. 15, no. 100968562, 2015, p. 1095, [https://doi.org/10.1186/s12889-015-2417-7](https://doi.org/10.1186/s12889-015-2417-7.)  Pearce, Margo. The Cedar Project : Understanding the Association between Childhood Maltreatment and Psychological Distress, Resilience, and HIV and HCV Vulnerability among Young Indigenous People Who Use Drugs in Three Canadian Cities. University of British Columbia, 2014, <http://hdl.handle.net/2429/51767.>  Pearce, Margo E., et al. ‘The Cedar Project: Risk and Protective Factors for Resilience among Young, Urban Indigenous People Who Use Drugs and Have Experienced Early Childhood Trauma’. XXth ISPCAN International Congress on Child Abuse and Neglect, Nagoya, Japan, September 14-17, 2014 [Abstracts], International Society for Prevention of Child Abuse and Neglect, 2014, p. 97. <https://doi.org/10.1037/e500792015-094.> | Canada | In this study, three questionnaires were administered to a cohort of young Indigenous peoples who use illicit drugs in three cities in British Columbia. Results from quantitative analyses indicate significant associations between resilience and having grown up in a home where their traditional language was often or always spoken, as well as currently knowing how to speak a traditional language. Resilience was defined as positive adaptation despite adversity and measured by the Connor-Davidson Resilience Scale. |
| Penney, C. A. ‘Interpretation for Inuit Patients Essential Element of Health Care in Eastern Arctic.’ CMAJ : Canadian Medical Association Journal = Journal de l’Association Medicale Canadienne, vol. 150, no. 11, 1994, pp. 1860–61. | Canada | The author of this article argues that providing interpretation services for Inuit patients is an essential element of health care in the Eastern Arctic, and highlights critical issues including the lack of training for interpreters, challenges translating across cultural difference, and the need to develop medical terminology in Indigenous languages. The author asserts that with appropriate training, language interpreters can bridge cultural gaps, provide patient support, generate community acceptance of medical management and aid in the development and delivery of culturally appropriate health education, thereby improving the quality of health care overall. |
| Perdue, David G., et al. ‘Culture and Colorectal Cancer Screening on Three American Indian Reservations.’ Ethnicity & Disease, vol. 21, no. 3, 2011, pp. 342–48. | USA | This study measuring the level of uptake of colorectal cancer screening among adults over age 50 living across three reservations in South Dakota and Arizona reports that individuals who spoke their traditional language at home were significantly less likely to have had an endoscopy in their lifetime than those who did not speak their language at home. Those who spoke an American Indian language were 40% less likely to report colonoscopy or flexible sigmoidoscopy as compared to English-only speakers. |
| Perry, Alta. Indigenous Knowledge and Health: Exploring and Comparing Mainstream Academic and Indigenous Community Perspectives. Simon Fraser University, 2009. Open WorldCat, <https://summit.sfu.ca/item/9786.> | Canada | In this ethnographic pilot study in a Secwepemc community, respondents shared the view that all Indigenous knowledge (including languages) is important to health, health of the individual and health of the community. When asked to prioritize aspects of Indigenous knowledge as important to health, the respondents ranked language first. |
| Phillips, Morgan. Understanding Resilience Through Revitalizing Traditional Ways of Healing in a Kanienkehaka Community. Concordia University, Aug. 2010. spectrum.library.concordia.ca, https://spectrum.library.concordia.ca/id/eprint/7071/. | Canada | Through examining individual narratives, this thesis explores how the Kanien’kehá:ka at Kahnawake approach healing and wellness through incorporating language and culture into public health and social services organizations. Resilience, community, self determination and the integration of traditional ways of healing are supported by learning and living the Kanien’kehá:ka language. |
| Pier, P. T. (1998). An exploratory study of community trauma and culturally responsive counseling with Chamorro clients [Doctoral dissertation, University of Massachusetts Amherst]. ProQuest Dissertations Publishing. https://www.proquest.com/docview/304461615?pq-origsite=gscholar&fromopenview=true | USA | This study involving interviews with counselors, psychologists, psychiatrists, and social workers who routinely treat Chamorro clients on Guam identifies Indigenous language loss as a cause of trauma stemming from American colonization and as an important issue for mental health practitioners to be aware of and incorporate into responsive counseling practices for this population. |
| Pitama, S., Ahuriri-Driscoll, A., Huria, T., Lacey, C., & Robertson, P. (2011). The value of te reo in primary care. Journal of Primary Health Care, 3(2), 123–127. | New Zealand | This qualitative study explores *te reo Māori* (Māori language) as a determinant of health from 30 Māori patients' perspectives. The ability of primary health care staff to speak *te reo* was reported as an important factor in cultural competency, an indicator of quality of care, and a supportive factor of Māori health gains as it contributes to relationships where patients feel valued. |
| Plank, G. A. (2001). Application of the Cross Battery Approach in the assessment of American Indian children: A viable alternative. American Indian and Alaska Native Mental Health Research, 10(1), 21–33. https://doi.org/10.5820/aian.1001.2001.21 | USA | This commentary discusses the importance of incorporating both language and culture into psychometric assessments to properly identify intellectual disabilities among American Indian children. |
| Plaud JJ, Schweigman K, & Welty TK. (1997). Health and depression among American Indians: Psychosocial data from the Strong Heart Study phase II. International Journal of Rehabilitation & Health, 3(1), 51–59. | USA | This quantitative study with upper midwest American Indian adults reports a relationship between lower scores on a depression scale and ability to speak one's native language. |
| Puna ET, Tiatia-Seath J (2017) Defining positive mental wellbeing for New Zealand-born Cook Islands youth. Journal of Indigenous Wellbeing: Te Mauri – Pimatisiwin 2, 97–107. | New Zealand | In this qualitative study involving interviews with New Zealand-born Cook Islands Indigenous youth, maintaining social and cultural connections, including language retention and revitalisation, are reported as being central to enhancing positive mental wellbeing and preventing suicide. |
| Quigley R, Mann J, Robertson J, Bonython-Ericson S. Are we there yet? Exploring the journey to quality stroke care for Aboriginal and Torres Strait Islander peoples in rural and remote Queensland. Rural and Remote Health 2019; 19: 4850. https://doi.org/10.22605 | Australia | In this survey study with stroke survivors, carers, and stakeholders in rural and remote Queensland, it is reported that stroke services can be made more culturally appropriate through including resources specific to the language, literacy, and cultural needs of Aboriginal and Torres Strait Islander populations as well as advocating for increased availability and use of language interpreters. |
| Redwood, Diana G., et al. ‘Traditional Foods and Physical Activity Patterns and Associations with Cultural Factors in a Diverse Alaska Native Population.’ International Journal of Circumpolar Health, vol. 67, no. 4, 2008, pp. 335–48. | USA | In this survey study with a population of Alaska Native people, speaking a Native language at home was associated with both eating traditional food and involvement in traditional physical activities. Individuals who reported consuming ten or more traditional foods or participating in three or more traditional physical activities in the past year were more likely to speak their Native language at home than those who consumed fewer traditional foods or engaged in less physical activities. |
| Redwood, Diana, et al. ‘Differences in Cigarette and Smokeless Tobacco Use among American Indian and Alaska Native People Living in Alaska and the Southwest United States’. Nicotine & Tobacco Research, vol. 12, no. 7, July 2010, pp. 791–96, https://doi.org/10.1093/ntr/ntq087. | USA | In this survey study with Navajo and Alaska Native people, speaking an Indigenous language at home was associated with current use of smokeless tobacco products, while current use of cigarettes or cigar products was associated with only speaking English in the home. |
| Redwood, Diana, et al. “Occupational and Environmental Exposures among Alaska Native and American Indian People Living in Alaska and the Southwest United States.” Journal of Environmental Health, vol. 74, no. 9, 2012, pp. 22–28. | USA | This quantitative study reports that Indigenous language use at home is associated with an increased likelihood of occupational and environmental hazard exposures (e.g. to petroleum products, military chemicals, asbestos, pesticides, and welding/silversmithing) for a sample of Alaska Native and American Indian peoples living in Alaska and the southwest United States. |
| Reilly, Rachel, et al. ‘Aboriginal Experiences of Cancer and Care Coordination: Lessons from the Cancer Data and Aboriginal Disparities (CanDAD) Narratives.’ Health Expectations, vol. 21, no. 5, Oct. 2018, pp. 927–36, https://doi.org/10.1111/hex.12687. | Australia | In this interview study with Aboriginal cancer patients, survivors, carers, and service providers at a major metropolitan hospital in South Australia, a lack of language interpreting services is identified as a critical issue leading to problems with information sharing and communication between patients and providers. A lack of cultural safety within hospitals is attributed in part to a lack of staff who speak Aboriginal languages. |
| Restoule, J. P. “Education as Healing : How Urban Aboriginal Men Described Post-Secondary Schooling as Decolonising.” Australian Journal of Indigenous Education, vol. 34, 2005, pp. 123–31, https://doi.org/10.1017/S132601110000404X. | Canada | In this study involving learning circles with urban Aboriginal men, learning Aboriginal languages in post-secondary settings is reported as a factor which contributes to healing. |
| Reyhner, Jon. ‘Indigenous Language Immersion Schools for Strong Indigenous Identities’. Heritage Language Journal, 2010 | USA | This article makes the case that Indigenous language immersion programs are vital to healing from the negative effects of colonialism and assimilationist schooling. |
| Richmond, Chantelle A. M. “The Social Determinants of Inuit Health: A Focus on Social Support in the Canadian Arctic.” International Journal of Circumpolar Health, vol. 68, no. 5, 2009, pp. 471–87. | Canada | This secondary analysis of the Arctic Supplement of the 2001 Aboriginal Peoples Survey reports that Inuit people who speak or understood an Aboriginal language are significantly more likely to report high levels of all types of social support (positive social interaction, emotional support, tangible support and affection and intimacy) than those who do not speak or understand an Aboriginal language. The authors connect this back to health through referencing social epidemiological literature which provides strong evidence that people with less access to social support tend to experience poorer health outcomes. |
| Robinson, Frances, et al. ‘Breast Cancer Education for Native American Women: Creating Culturally Relevant Communications.’ Clinical Journal of Oncology Nursing, vol. 9, no. 6, 2005, pp. 689–92. | USA | This commentary discusses how incorporating language considerations into breast cancer education for Navajo women improves cultural relevance. |
| Robson, Bridget. ‘Editorial: Te Reo Māori Is Imperative for Research and Practice in Aotearoa. He Taonga Te Reo. Kōrerotia ! Tuhia!’ Nursing Praxis in New Zealand, vol. 32, no. 3, Nov. 2016, pp. 5–6, https://doi.org/10.36951/ngpxnz.2016.009. | New Zealand | This editorial asserts the critical role of *te reo* *Māori* in health care in Aotearoa New Zealand, including how using the language helps people to feel more respected and fosters good relationships with patients. |
| Rodriguez-Lonebear, D., Barcelo, N. E., Akee, R., & Carroll, S. R. (2020). American Indian Reservations and COVID-19: Correlates of Early Infection Rates in the Pandemic. Journal of Public Health Management and Practice, 26(4), 371–377. https://doi.org/10.1097/PHH.0000000000001206 | USA | This quantitative study in Oklahoma reports that COVID-19 rates were lower on reservations with higher percentages of English language-only households. |
| Royal Commission of Aboriginal Peoples, Gathering Strength: Report of the Royal Commission on Aboriginal Peoples. Volume 3, 1996, p. 730, https://www.bac-lac.gc.ca/eng/discover/aboriginal-heritage/royal-commission-aboriginal-peoples/Pages/item.aspx?IdNumber=402. | Canada | In this report drawing on testimonies collected by the Royal Commission on Aboriginal Peoples, language is referenced throughout as both a harmful element of colonization (loss of language and the imposition of a colonial language) and as an important part of health and healing. |
| Rumbold, Alice R., et al. ‘Challenges to Providing Fetal Anomaly Testing in a Cross-Cultural Environment: Experiences of Practitioners Caring for Aboriginal Women’. Birth: Issues in Perinatal Care, vol. 42, no. 4, Dec. 2015, pp. 362–68. psyh, 2015-37043-001, EBSCOhost, https://doi.org/10.1111/birt.12182. | Australia | In this interview study examining health care practitioners' perceptions of barriers to providing fetal anomaly testing for Aboriginal women in the Northern Territory, a prevalent theme is communicating across language differences. Explaining the concept of “risk” when delivering screening test results is identified as particularly challenging on account of a perceived lack of an equivalent concept in Aboriginal languages. Aboriginal health practitioners are identified as being able to assist with overcoming language barriers as they are knowledgeable facilitators as well as interpreters, but it is reported that they are underutilized. |
| Running Bear, U., Croy, C. D., Kaufman, C. E., Thayer, Z. M., Manson, S. M., & The AI-SUPERPFP Team. (2018). The relationship of five boarding school experiences and physical health status among Northern Plains Tribes. Quality of Life Research, 27(1), 153–157. https://doi.org/10.1007/s11136-017-1742-y | USA | In this secondary analysis of quantitative data about Northern Plains American Indian adults who attended boarding schools, a relationship is reported between lower physical health scores and the experience of having been punished for using their Indigenous language in school. The effect was strongest among those who were aged 8 or older when they entered boarding school. |
| Russell, L. (2018). Te Oranga Hinengaro: Report on Māori Mental Wellbeing Results from the New Zealand Mental Health Monitor & Health and Lifestyles Survey. Wellington: Health Promotion Agency/Te Hiringa Hauora. | New Zealand | This report of the Māori Mental Wellbeing Results from three nationwide surveys reports a strong connection between proficiency in te reo Māori and a connection to culture, which is seen as key for mental wellbeing. When looked at alone, no link was reported between ability to speak te reo Māori well or very well and the probability of feeling isolated. |
| Ryan, C. J., Cooke, M., & Leatherdale, S. T. (2016). Factors associated with heavy drinking among off-reserve First nations and Métis youth and adults: Evidence from the 2012 Canadian Aboriginal Peoples Survey. Preventive Medicine, 87, 95–102. http://dx.doi. org/10.1016/j.ypmed.2016.02.008. | Canada | In this analysis of data from the 2012 Aboriginal Peoples Survey, neither speaking an Aboriginal language nor exposure to an Aboriginal language were significantly correlated in either direction with heavy drinking among First Nations or Métis youth or adults. |
| Ryan, Christopher J., et al. ‘A Cross-Sectional Examination of the Correlates of Current Smoking among off-Reserve First Nations and Métis Adults: Evidence from the 2012 Aboriginal Peoples Survey.’ Addictive Behaviors, vol. 54, no. 2gw, 7603486, 2016, pp. 75–81, https://doi.org/10.1016/j.addbeh.2015.12.004. | Canada | In this analysis of data about First Nations and Métis adults from the 2012 Aboriginal Peoples Survey, people who spoke an Aboriginal language or were exposed to Aboriginal languages at home and in the community were more likely to be current smokers than those who neither spoke the language nor were exposed to the language at home or in the community. |
| Ryan, Christopher J., et al. ‘The Correlates of Current Smoking among Adult Métis: Evidence from the Aboriginal Peoples Survey and Métis Supplement.’ Canadian Journal of Public Health = Revue Canadienne de Sante Publique, vol. 106, no. 5, 2015, pp. e271-6, https://doi.org/10.17269/cjph.106.5053. | Canada | In this analysis of data about Métis adults from the 2006 Aboriginal Peoples Survey and Métis supplement, people who spoke an Aboriginal language and/or or lived in a house where an Aboriginal language was spoken were more likely to be current smokers than those who neither spoke the language nor lived in a house where an Aboriginal language was spoken. |
| Ryan, Christopher J., et al. ‘The Correlates of Physical Activity among Adult Métis.’ Ethnicity & Health, vol. 23, no. 6, Aug. 2018, pp. 629–48, https://doi.org/10.1080/13557858.2017.1294655. | Canada | In this analysis of data about Métis adults from the 2006 Aboriginal Peoples Survey and Métis supplement, speaking an Aboriginal language was not associated with leisure-time physical activity or active transportation (walking). |
| Ryan, Christopher, et al. “Factors Associated with Current Smoking among Off-Reserve First Nations and Métis Youth: Results from the 2012 Aboriginal Peoples Survey.” The Journal of Primary Prevention, vol. 38, no. 1–2, Apr. 2017, pp. 105–19, https://doi.org/10.1007/s10935-016-0456-1. | Canada | In this analysis of data about First Nations and Métis youth aged 15-17 from the 2012 Aboriginal Peoples Survey, youth with knowledge of an Aboriginal language were more likely to be current smokers. |
| Salmon, Minette, et al. Defining the Indefinable: Descriptors of Aboriginal and Torres Strait Islander Peoples’ Cultures and Their Links to Health and Wellbeing. Lowitja Institute, 1 Sept. 2019, https://apo.org.au/node/264456. | Australia  Canada  New Zealand  USA | In this literature review, language was one of the six cultural domains identified as enabling or being related to producing good health and wellbeing among Indigenous peoples. |
| Schmidt A. The loss of Australia’s Aboriginal language heritage. Canberra: Aboriginal Studies Press; 1990. (The Institute Report Series). | Australia | In Chapter 2 of this report, an argument presented for keeping Aboriginal languages strong is that they are key to self-esteem. A case study is provided which supports this. |
| Schulling, Sharon Kay. Differences and Similarities between Yup’ik Generic Care and Professional Nursing Care: Implications for Nursing Education. University of Nebraska, 2003. | USA | In this interview study with Yup'ik healthcare providers, when asked “What do you think is the most important thing to know about when caring for someone from your culture?“, one-third commented on issues around the use of language. They described how speaking and understanding the Yup’ik language is a key to fully understanding Yup’ik culture, and that failure to speak the language creates barriers to communication. |
| Schultz, R., Abbott, T., Yamaguchi, J., & Cairney, S. (2018). Indigenous land management as primary health care: Qualitative analysis from the Interplay research project in remote Australia. BMC Health Services Research, 18(1), Article 960. https://doi.org/10.1186/s12913-018-3764-8 | Australia | In this study involving community focus groups and interviews to develop a wellbeing framework by and for Indigenous Australians in remote regions, Indigenous languages are identified in the final model as a factor that enhances wellbeing. |
| Schultz, R., Quinn, S. J., Abbott, T., Cairney, S., & Yamaguchi, J. (2019). Quantification of interplaying relationships between wellbeing priorities of Aboriginal people in remote Australia. The International Indigenous Policy Journal, 10(3). doi: https://doi.org/10.18584/iipj.2019.10.3.8165 | Australia | This article describes statistical modeling of community wellbeing priorities based on surveys administered to Aboriginal and Torres Strait Islanders living in four remote settlements. Cultural practice was associated with Aboriginal language literacy and empowerment, and both were associated with wellbeing. Language literacy had the strongest relationship to wellbeing of all factors in the study, and for women in particular. |
| Schumacher, Mary Catherine, et al. ‘Prevalence and Predictors of Cancer Screening among American Indian and Alaska Native People: The EARTH Study.’ Cancer Causes & Control : CCC, vol. 19, no. 7, 2008, pp. 725–37, https://doi.org/10.1007/s10552-008-9135-8. | USA | This survey study of Alaska Native people and people living on Navajo reservations in Arizona and New Mexico identifies that individuals who spoke only English at home, compared to those who spoke their Native language, were more likely to have received a Pap test in the past three years, and also more likely to have received a colonoscopy or sigmoidoscopy. The finding was most apparent in Alaska. The same relationship was not identified for mammography screening uptake. |
| Shahid S, et al. ‘Barriers to Participation of Aboriginal People in Cancer Care: Communication in the Hospital Setting.’ Medical Journal of Australia, vol. 190, no. 10, May 2009, pp. 574–79, https://doi.org/10.5694/j.1326-5377.2009.tb02569.x. | Australia | This qualitative study about Aboriginal patients’ views on effective communication between Aboriginal people and health service providers in Western Australian hospital settings reports that language barriers are one of many factors which impede communication and erode trust with care providers. The authors report that there were no Aboriginal language interpreter services available in tertiary hospitals in Perth at the time of the study. |
| Sharma, Richa, et al. “The Cedar Project: Historical, Structural and Interpersonal Determinants of Involvement in Survival Sex Work over Time among Indigenous Women Who Have Used Drugs in Two Canadian Cities.” The International Journal on Drug Policy, vol. 87, no. 9014759, 2020, p. 103012, https://doi.org/10.1016/j.drugpo.2020.103012. | Canada | In this study, questionnaires were administered to a cohort of young Indigenous peoples who use illicit drugs in three cities in British Columbia. Results from quantitative analyses did not identify a link between any measure of Indigenous language speaking or knowledge and current involvement in survival sex work. |
| Shea, Haley, et al. “Cultural Revitalization as a Restorative Process to Combat Racial and Cultural Trauma and Promote Living Well.” Cultural Diversity and Ethnic Minority Psychology, vol. 25, no. 4, Oct. 2019, pp. 553–65, https://doi.org/10.1037/cdp0000250. | USA | In this study involving ethnographic observation, interview, and survey methods, Myaamia college students and community members expressed that reclaiming one’s culture and language had an impact on restoring wellness among the Miami Tribe. |
| Shield, J. M., Kearns, T. M., Garngulkpuy, J., Walpulay, L., Gundjirryirr, R., Bundhala, L., Djarpanbuluwuy, V., Andrews, R. M., & Judd, J. (2018). Cross-Cultural, Aboriginal Language, Discovery Education for Health Literacy and Informed Consent in a Remote Aboriginal Community in the Northern Territory, Australia. Tropical Medicine and Infectious Disease, 3(1). https://doi.org/10.3390/tropicalmed3010015 | Australia | This article details the development of an oral health literacy program for a remote Australian Aboriginal community where community feedback supported incorporating the local language and culture as important and effective components of the program. |
| Shrivastava, Richa, et al. ‘Relational Continuity of Oral Health Care in Indigenous Communities: A Qualitative Study.’ BMC Oral Health, vol. 19, no. 1, 2019, p. 287, https://doi.org/10.1186/s12903-019-0986-z. | Canada | This qualitative study involved interviews with oral health patients, providers and administrators in Cree communities in Northern Québec to explore the relational continuity of oral health care, defined as an ongoing relationship between patients and providers that connects care over time and bridges discontinuous events. Language barriers are reported as a major barrier to relational continuity, especially in case of children and elderly patients. One enabler was promoting effective communication, which included developing specific cultural competency training, healthcare professionals learning the Cree language, and engaging with communities to develop a Cree dental glossary. |
| Sibthorpe B, et al. ‘Self-Assessed Health among Indigenous Australians: How Valid Is a Global Question?’ American Journal of Public Health, vol. 91, no. 10, Oct. 2001, pp. 1660–63, https://doi.org/10.2105/AJPH.91.10.1660. | Australia | In this analysis of data from the 1994 National Aboriginal and Torres Strait Islander Survey (NATSIS), a link was reported between not speaking English as a primary language at home and self-reporting fair or poor health, even after accounting for other factors. |
| Sivak, Leda, et al. ‘“Language Breathes Life“-Barngarla Community Perspectives on the Wellbeing Impacts of Reclaiming a Dormant Australian Aboriginal Language’. International Journal of Environmental Research and Public Health, vol. 16, no. 20, MDPI, Oct. 2019, https://doi.org/10.3390/ijerph16203918.  Sivak, Leda, et al. “Can the Revival of Indigenous Languages Improve the Mental Health and Social and Emotional Wellbeing of Aboriginal and Torres Strait Islander People?“ TheMHS E-Book of Proceedings, 2019, https://portal.sahmriresearch.org/en/publications/can-the-revival-of-Indigenous-languages-improve-the-mental-health. | Australia | The results reported in this qualitative interview study reflect the vital importance of Barngarla language to Barngarla identity, spirituality, connectedness, and cultural, social, and emotional wellbeing. |
| Smylie JA; Aboriginal Health Issues Committee. A guide for health professionals working with Aboriginal peoples: cross cultural understanding. SOGC Policy Statement. J Obstet Gynaecol Can 2001;23(2):157-67 | Canada | In this guide for health professionals working with Aboriginal peoples, Aboriginal languages are mentioned frequently. One recommendation is that Aboriginal peoples should receive treatment in their own languages, whenever possible. Reasons behind this are so that Aboriginal peoples can be full participants in decision making, overcome language and communication barriers, and receive culturally appropriate health care. |
| Steinhauer, Diana, and James Lamouche. ‘Miyo-Pimâtisiwin, “A Good Path“: Indigenous Knowledges, Languages, and Traditions in Education and Health’. Determinants of Indigenous Peoples’ Health, edited by Margo Greenwood et al., Second, Canadian Scholars’ Press, 2018. | Canada | The authors of this chapter describe how from an Indigenous perspective, health is linked to language as well as to other factors including culture, politics, land, and social relationships. |
| Stewart, T., et al. ‘A Navajo Health Consumer Survey.’ Medical Care, 1980, http://hslic-nhd.health.unm.edu/Articles/Details/9701099. | USA | In this study involving the distribution of a health consumer survey to Navajo families in three areas of a Navajo reservation, issues arising from the translation of English to Navajo was identified as a major concern in their encounters with medical providers. |
| Stout, Madeleine Dion. Atikowisi Miýw-Ayawin, Ascribed Health and Wellness, to Kaskitamasowin Miýw-Ayawin, Achieved Health and Wellness: Shifting the Paradigm. Determinants of Indigenous Peoples' Health. Edited by Margo Greenwood et al., Second, Canadian Scholars’ Press, 2018. | Canada | In this chapter, the author argues for expanding the determinants of health to include Indigenous languages and other factors that are “central to the social and cultural thought leadership of Indigenous peoples.” |
| Sur, Roger, et al. “A Pacific Islander Organization’s Approach Towards Increasing Community Colorectal Cancer Knowledge and Beliefs.” Californian Journal of Health Promotion, vol. 11, no. 2, June 2013, pp. 12–20, https://doi.org/10.32398/cjhp.v11i2.1527. | USA | This study involving workshops with Pacific Islander adults in California examined an educational intervention about colorectal cancer to determine if changes in knowledge and beliefs were associated with positive intentions to pursue screening. A key finding is that it is important to tailor health screening education programs to the linguistic needs of communities. |
| Taff, A., Chee, M., Hall, J., Hall, M. Y. D., Martin, K. N., & Johnston, A. (2018). Indigenous language use impacts wellness. In K. L. Rehg & L. Campbell (Eds.), The Oxford handbook of endangered languages (pp. 861–884). Oxford University Press. https://doi.org/10.1093/oxfordhb/9780190610029.013.41 | Australia  Canada  USA | In this chapter, eye witness accounts indicate that loss of language leads to shame, lack of direction, stress, tears, and alcohol and drug abuse, while persistent language use leads to pleasure, joyful emotion, wellness, balance, a healthy spirit, identity, and direction. Literature is cited reporting that language use reduces suicide rates, diabetes symptoms, and risk factors for youth, and that learning any language benefits cognitive health. |
| Tanjasiri, Sora Park, et al. ‘Exploring Access to Cancer Control Services for Asian-American and Pacific Islander Communities in Southern California.’ Ethnicity & Disease, vol. 14, no. 3 Suppl 1, 2004, pp. S14-9. | USA | This article reports preliminary findings of a mapping study which identified that Chamorro and Samoan communities in two California counties face significant geographic barriers to linguistically appropriate health and social services. This was despite the fact that the two counties studied comprise some of the largest populations of their nationalities in the state. Improvements in access to medical interpretation are recommended. |
| Tapia, K. A., Garvey, G., McEntee, M. F., Rickard, M., Lydiard, L., & Brennan, P. C. (2019). Breast screening attendance of Aboriginal and Torres Strait Islander women in the Northern Territory of Australia. Australian and New Zealand Journal of Public Health, 43(4), 334–339. https://doi.org/10.1111/1753-6405.12917 | Australia | In this study involving quantitative analyses of data from a national population‐based breast screening program, it is reported that Aboriginal and Torres Strait Islander women in the Northern Territory who spoke a main language other than English at home were less likely to have had a mammogram than those who spoke English as their main language. |
| Taylor, K. A. (2010). Intercultural communication in Central Australian Indigenous health care: A critical ethnography. [Doctoral thesis, The Flinders University of South Australia]. https://trove.nla.gov.au/work/234595362?keyword=indigenous%20languages%20and%20health&l-advlanguage=English&l-format=Thesis | Australia | This study uses critical ethnography to examine health care communication from the perspective of Indigenous first language speaking clients and English first language healthcare professionals. Both parties describe multiple barriers and frustrations with communications, a lack of cultural safety, and ongoing colonizing practices, attitudes, beliefs and power structures. While there was a tacit acceptance of these barriers, both groups desired more effective dialogue and better relationships. |
| Taylor, Kerry A., et al. ‘Intercultural Communications in Remote Aboriginal Australian Communities: What Works in Dementia Education and Management?’ Health Sociology Review, vol. 21, no. 2, June 2012, pp. 208–19, https://doi.org/10.5172/hesr.2012.21.2.208. | Australia | This study evaluates a targeted dementia awareness resource that was piloted in English and three Aboriginal languages in remote Aboriginal communities. It is reported that presenting a dementia awareness resource in local Aboriginal languages helps to validate culture, enhance engagement, and provide an opportunity to develop health vocabularies where there are no existing translatable terms, all of which are key components of effective intercultural communication. |
| Teevale, Tasileta, et al. ‘Binge Drinking and Alcohol-Related Behaviours amongst Pacific Youth: A National Survey of Secondary School Students.’ The New Zealand Medical Journal, vol. 125, no. 1352, 2012, pp. 60–70. | New Zealand | In this analysis of data from a nationally representative survey, youth whose parents spoke a Pacific language were reported to be less likely to binge drink than other youth populations. No association was reported between the youth's own use of a Pacific language and binge drinking. |
| Temple, J. B., & Russell, J. (2018). Food Insecurity among Older Aboriginal and Torres Strait Islanders. International Journal of Environmental Research and Public Health, 15(8). https://doi.org/10.3390/ijerph15081766 | Australia | In this analysis of data from the 2012–2013 National Aboriginal and Torres Strait Islander Nutrition and Physical Activity Survey, measures of Indigenous language use were associated with exposure to food insecurity among older Aboriginal and Torres Strait Islander people. The effect was more pronounced for people living in remote areas. |
| Teng, Andrea, et al. ‘What Protects against Pre-Diabetes Progressing to Diabetes? Observational Study of Integrated Health and Social Data.’ Diabetes Research and Clinical Practice, vol. 148, no. ebi, 8508335, 2019, pp. 119–29, https://doi.org/10.1016/j.diabres.2018.12.003. | New Zealand | In this study involving quantitative analyses of data on adults with pre-diabetes in the upper North Island of New Zealand, speaking *te reo Māori* was reported to be a protective factor against the progression of pre-diabetes to diabetes. |
| Thieberger, Nicholas. ‘Language Maintenance: Why Bother?’ Multilingua, vol. 9, no. 4, 1990, pp. 333–58. | Australia | This study draws on existing literature to assert that individual well-being is one of the seven principal arguments for maintaining Australian Aboriginal languages, with a primary focus on the cognitive benefits of bilingualism. |
| Thompson, E. J. C. (2012). ‘Hedekeyeh Hots’ih Kahidi’—’Our Ancestors Are in Us’: Strengthening our voices through language revitalization from a Tahltan worldview [Doctoral dissertation, University of Victoria]. https://dspace.library.uvic.ca:8443/handle/1828/4213?show=full | Canada | In this study involving conversations with co-researchers, it is reported that language revitalization for Tahltan people can be the start of a process in which they begin to heal from the impacts of past losses as it allows their voices to become stronger and healthier. The author describes the connection between language revitalization and healing as forms of empowerment. |
| Thunderbird Partnership foundation, & Health Canada. (2015). First Nations mental wellness continuum framework. Health Canada. http://epe.lac-bac.gc.ca/100/201/301/weekly_checklist/2015/internet/w15-12-F-E.html/collections/collection_2015/sc-hc/H34-278-1-2014-eng.pdf | Canada | This report describes the development of The First Nations Mental Wellness Continuum Framework through regional discussions, a national gathering, and a national validation and implementation session. The framework includes culture (including language) as the foundational and underlying theme for mental wellness, holding all other components together. Identity, a key element of spiritual wellness, is derived from factors such as language, land, and ancestry. The loss of language has contributed to trauma, the result of which is linked in the report to higher rates of suicide, mental health and addictions issues as well as other significant disparities in health, while the maintenance of language and culture are reported to have ensured strength and resilience, enhancing the skills and knowledge of individuals, families and communities, thereby improving mental wellness. It is also reported that effective health promotion and prevention strategies draw on local languages, while language barriers such as lack of translation services can impede access to services and lead to the deterioration of health. |
| Townsend, C. K. M. (2014). Impacts of hawaiian language loss and promotion via the linguistic landscape [Doctoral dissertation, University of Hawaiʻi at Mānoa]. ProQuest Dissertations Publishing. https://www.proquest.com/docview/1652530324?pq-origsite=gscholar&fromopenview=true | USA | In this focus group study, Native Hawaiian people expressed a belief that learning the language strengthens their cultural identity, self-esteem, and the Hawaiian community, and that a bilingual landscape would improve the status of the Hawaiian language and Native Hawaiian health. Support among Hawaiian citizens for a bilingual linguistic landscape was associated with a belief in the positive impact of bilingual signage on the Hawaiian language and Native Hawaiian health. |
| Tsethlikai, Monica. ‘An Exploratory Analysis of American Indian Children’s Cultural Engagement, Fluid Cognitive Skills, and Standardized Verbal IQ Scores’. Developmental Psychology, vol. 47, no. 1, Jan. 2011, pp. 192–202, https://doi.org/10.1037/a0020803. | USA | This cross-sectional quantitative study reports that cognitive skill scores vary as a function of Tohono O’odham language knowledge and age: children who understand and/or speak Tohono O’odham started out with lower average cognitive skill scores than did children with no language knowledge, but mean scores generally increased in the older age groups such that they were equal to or higher than those of non-speakers by age 9. |
| van Beek, S. (2016). Intersections: Indigenous Language, Health and Wellness [Undergraduate Thesis, University of British Columbia]. https://fpcc.ca/wp-content/uploads/2020/08/Intersections_Indigenous_Language_Health_and_Wellness_WebVersion.pdf | Australia  Canada  New Zealand  USA | This literature review reports links between Indigenous language use and reduced diabetes, suicide, and HIV rates. The author reports on several themes in the literature: indicators of health and wellbeing, academic success, Indigenous wellness concepts, identity, and resilience. Culture and language are reported to be frequently tied together in the literature. |
| Van Bewer, V., & Woodgate, R. L. (2017). Examining the correlates of current smoking among off-reserve First Nations, Métis and Inuit youth: Evidence from the 2012 Aboriginal Peoples Survey. Addictive Behaviors, 69, 93–97. https://doi.org/10.1016/j.addbeh.2017.02.017 | Canada | This analysis of data from the 2012 Aboriginal Peoples Survey reports a correlation between speaking an Aboriginal language and current smoking among First Nations, Métis and Inuit youth. |
| Vass, Alyssa, et al. ‘Health Literacy and Australian Indigenous Peoples: An Analysis of the Role of Language and Worldview’. Health Promotion Journal of Australia, vol. 22, no. 1, 2011, pp. 33–37, https://doi.org/10.1071/HE11033. | Australia | Drawing on the professional experiences of the authors and their Yolŋu and non-Indigenous colleagues at community development organization in the Northern Territory, the authors report that incorporating language is an integral part of health education when working with Indigenous peoples whose first language is not English. |
| Verney, Steven P., et al. ‘Cultural Considerations in the Neuropsychological Assessment of American Indians/Alaska Natives’. Minority and Cross-Cultural Aspects of Neuropsychological Assessment: Enduring and Emerging Trends., 2nd Ed., edited by F. Richard Ferraro, Taylor & Francis, 2016, pp. 115–58. | USA | This literature review identifies language as an important cultural consideration that impacts the validity of neuropsychological assessments of American Indian and Alaska Native populations. |
| Verney, Steven P., et al. ‘The Associations among Sociocultural Factors and Neuropsychological Functioning in Older American Indians: The Strong Heart Study’. Neuropsychology, vol. 33, no. 8, Nov. 2019, pp. 1078–88, https://doi.org/10.1037/neu0000574. | USA | This study looks at a group of American Indian people aged 60 and older who were fluent in English and had varying proficiencies in their traditional languages. Higher scores on three neuropsychological and motor tests were associated with less ability to speak a Native American language. There was no association between language ability and two tests of verbal memory. |
| Vining, Christine, et al. ‘Speech-Language Assessment Considerations for American Indian and Alaska Native Children Who Are Dual Language Learners.’ Perspectives of the ASHA Special Interest Groups, vol. 2, no. 14, Oct. 2017, pp. 29–40, https://doi.org/10.1044/persp2.SIG14.29. | USA | The authors of this article argue that speech-language pathologists can play a critical role in reducing the overrepresentation of American Indian and Alaska Native children in special education by using a holistic assessment process that is responsive to the communication patterns used at home and in community contexts. The authors also provide a framework for distinguishing actual language disorders from differences associated with cultural and linguistic diversity. |
| Waldram, J. B. (1990a). Access to traditional medicine in a western Canadian city. Medical Anthropology, 12(3), 325–348. | Canada | In this secondary analysis of interviews with urban Indigenous peoples in Saskatoon, participants who spoke an Indigenous mother tongue or who spoke their Indigenous language most of the time were more likely to desire access to a traditional healer, more likely to access a traditional healer if one was available, and more likely to believe in the efficacy or the existence of traditional Indigenous medicine. |
| Waldram, James B. ‘The Persistence of Traditional Medicine in Urban Areas: The Case of Canada’s Indians.’ American Indian and Alaska Native Mental Health Research, vol. 4, no. 1, Jan. 1990b, pp. 9–29. | Canada | This examination of previous literature reports associations between speaking an Indigenous language and seeking the services of a traditional Indigenous healer as well as believing in the superiority of traditional medicines for some medical problems. |
| Watson, R., Castleden, H., Tui’kn Partnership, Masuda, J., King, M., & Stewart, M. (2012). Identifying gaps in asthma education, health promotion, and social support for Mi’kmaq families in Unama’ki (Cape Breton), Nova Scotia, Canada. [Erratum appears in Prev Chronic Dis 2013;10. Http://www.cdc.gov/pcd/issues/2013/12_0039e.htm.]. Preventing Chronic Disease, 9(101205018). https://doi.org/10.5888/pcd9.120039 | Canada | In this study, several of the caregivers of Mi’kmaq families in Unama’ki (Cape Breton) who were interviewed indicate that language barriers contribute to asthma support gaps for children and youth, including a lack of health promotion materials in their language and challenges understanding English. It is suggested that future community-based health interventions and health-promoting resources should be provided in the Mi’kmaq language. |
| Whalen, D. H., Moss, M., & Baldwin, D. (2016). Healing through language: Positive physical health effects of Indigenous language use [version 1; peer review: 2 approved with reservations]. F1000Research, 5(852). <https://f1000research.com/articles/5-852> | Canada  USA | The authors of this article review literature and existing language programs. They report positive physical health benefits of language maintenance and revitalization including decreased youth suicide and diabetes rates, less smoking, having generally good health and wellness, practicing traditional health promoting behaviours, and reduced risk factors including alcohol consumption, illicit drug use, and violence victimization. |
| Whitbeck, L. B., Adams, G. W., Hoyt, D. R., & Chen, X. (2004). Conceptualizing and Measuring Historical Trauma Among American Indian People. American Journal of Community Psychology, 33(3–4), 119–130. https://doi.org/10.1023/B:AJCP.0000027000.77357.31 | Canada  USA | The authors of this survey study report that loss of language is a key element of historical trauma for the adult American Indian population studied in the upper Midwest of the United States and Ontario. One third of respondents thought daily or several times per day about loss of traditional language, and these thoughts were associated with negative feelings including anxiety, depression, anger and avoidance. |
| Wilgosh, L., Mulcahy, R., & Watters, B. (1986). Assessing intellectual performance of culturally different, Inuit children with the WISC--R. Canadian Journal of Behavioural Science / Revue Canadienne Des Sciences Du Comportement, 18(3), 270–277. https://doi.org/10.1037/h0079994 | Canada | This study of 366 Inuit children for whom English is a second language reports that more than three-quarters were diagnostically misclassified by a commonly applied cognitive assessment scale, largely due to reasons of somewhat limited English language verbal comprehension. |
| Williams, Shayne T. (2011). The importance of teaching and learning Aboriginal languages and cultures: The triangularity between language and culture, educational engagement, and community cultural health and wellbeing: A literature based research study for the New South Wales context. Sydney: New South Wales Department of Education and Communities: Aboriginal Affairs. [https://www.Aboriginalaffairs.nsw.gov.au/media/website_pages/research-and-publications/completed-research-and-evaluation/Final-report-final-version.pdf](https://www.aboriginalaffairs.nsw.gov.au/media/website_pages/research-and-publications/completed-research-and-evaluation/Final-report-final-version.pdf) | Australia | The author of this study draws on previous literature to argue for the importance of maintaining or revitalizing mother tongues for cultural health and wellbeing, which in turn support spiritual health. |
| Wilson, R. D. (1997). Bringing them home: Report of the National Inquiry into the Separation of Aboriginal and Torres Strait Islander Children from Their Families. Human Rights and Equal Opportunity Commission. https://humanrights.gov.au/our-work/bringing-them-home-report-1997 | Australia | In this human rights commission report describing the experiences of Aboriginal and Torres Strait people affected by forced removal from family homes, not allowing the children to speak their languages is identified throughout as a source of pain, grief, loss of identity, and separation from family, community and culture. Resulting language differences inhibit family reunions and the rebuilding of relationships. |
| Wolsko, C., Mohatt, G. V., Lardon, C., & Burket, R. (2009). Smoking, chewing, and cultural identity: Prevalence and correlates of tobacco use among the Yup’ik—The Center for Alaska Native Health Research (CANHR) study. Cultural Diversity and Ethnic Minority Psychology, 15(2), 165–172. https://doi.org/10.1037/a0015323 | USA | In this survey study of Yup'ik people residing in the Yukon-Kuskokwim Delta region of Alaska, smokeless tobacco users spoke their traditional language more frequently than those who smoked cigarettes only and those who did not use any tobacco. |
| Wright, S. C., & Taylor, D. M. (1995). Identity and the language of the classroom: Investigating the impact of heritage versus second language instruction on personal and collective self-esteem. Journal of Educational Psychology, 87(2), 241–252. https://doi.org/10.1037/0022-0663.87.2.241 | Canada | This study measured the personal and collective self-esteem of Inuit, White, and mixed-heritage (Inuit-White) children living in a subarctic community before and after their 1st year in either a heritage language or English/French program. Early heritage language education was reported to have a positive effect on the personal and collective self-esteem of Inuit students—a benefit that was not provided by English or French language instruction. |
| Yashadhana, A., Fields, T., Blitner, G., Stanley, R., & Zwi, A. B. (2020). Trust, culture and communication: Determinants of eye health and care among Indigenous people with diabetes in Australia. BMJ Global Health, 5(1), e001999. https://doi.org/10.1136/bmjgh-2019-001999 | Australia | In this study involving interviews and focus groups with Indigenous people with diabetes and primary care clinicians in four remote Indigenous communities in Australia, the authors report that patients had relatively low knowledge of how diabetes affects eye health, and that limited access to interpreters and language barriers were determining factors in eye health and care. Patients assessed as having a limited knowledge of diabetic eye care were mainly located in the two communities where Indigenous languages are predominantly spoken. Language differences were reported to be a key barrier to effective communication between patients and clinicians and to understanding and accessing meaningful health information. |
| Young, T. K. (1996). Sociocultural and behavioural determinants of obesity among Inuit in the central Canadian Arctic. Social Science & Medicine, 43(11), 1665–1671. | Canada | This quantitative study, including questionnaires and body size measurements, identifies links between some measures of obesity and fluency in the Inuit language among Inuit people in the central Canadian Arctic. |
| Zhang, Y., Lee, E. T., Cowan, L. D., North, K. E., Wild, R. A., & Howard, B. V. (2005). Hysterectomy prevalence and cardiovascular disease risk factors in American Indian women. Maturitas, 52(3–4), 328–336. | USA | In this quantitative study of American Indian women, an association between speaking their native language less frequently and higher hysterectomy prevalence was initially identified that disappeared after factoring in the geographic location of the women. |
| Zienczuk, N., & Egeland, G. M. (2012). Association between socioeconomic status and overweight and obesity among Inuit adults: International Polar Year Inuit Health Survey, 2007-2008. International Journal of Circumpolar Health, 71(1), 1–7. https://doi.org/10.3402/ijch.v71i0.18419 | Canada | This analysis of data from the 2007-2008 International Polar Year Inuit Health Survey reports that the prevalence of overweight and obese people was higher among those who spoke only English at home or both Inuit and English at home compared to those who only spoke an Inuit language at home. A gradient is reported where odds of an at-risk BMI increases if both an Inuit language and English are spoken at home and even further increases if only English is spoken at home. |
| Zuckermann, Ghil’ad, and Michael Walsh. ‘“Our Ancestors Are Happy!“ Revivalistics in the Service of Indigenous Wellbeing’. Indigenous Languages: Their Value to the Community: FEL XVIII Okinawa, 2013, https://www.adelaide.edu.au/directory/ghilad.zuckermann?dsn=directory.file;field=data;id=34174;m=view.  Zuckermann, Ghil‘ad. ‘Our Ancestors Are Happy: Language Revival and Mental Health’. Revitalistics, Oxford University Press, 2020, DOI: 10.1093/oso/9780199812776.001.0001. | Australia | With a focus on the Australian context, this work draws on previous literature in support of the conceptualization of language as core to Indigenous peoples' wellbeing. By drawing on a case study of a current reclamation project with the Barngarla Aboriginal people, the authors hypothesize that language reclamation often results in mental health empowerment. |
